# EpidemicKabu a new method to identify epidemic waves and their peaks and valleys

**DOI:** 10.1101/2024.03.11.24304124

**Authors:** Lina Marcela Ruiz Galvis, Anderson Alexis Ruales Barbosa, Oscar Ignacio Mendoza Cardozo, Noël Christopher Barengo, Jose L. Peñalvo, Paula Andrea Diaz Valencia

## Abstract

**INTRODUCTION:** The dynamical behavior of epidemic curves is an oscillation between a very low and very high number of incident cases throughout the time. These oscillations are commonly called waves of the epidemic curve. The concept of epidemic waves lacks a consensual definition and a simple methodology that can be used for many diseases.

**OBJECTIVE:** We describe in this study the EpidemicKabu method to identify the start and the end of past epidemic waves but also their peaks and valleys.

**METHOD:** The methodology is divided into processing of the curve, waves detection, and peaks and valleys delimitation. For processing the curve, a Gaussian kernel was used to diminish the noise and to smooth the curve. The first and second derivatives of the curve were used for the detection of waves, delimitation of peaks and valleys. The methodology was derived into the open access library. The method was tested using COVID-19 daily cases reported between 2020 and 2022 for different countries. After detection of waves, we made some measures related to the size of the waves for those countries.

**RESULTS:** The results of the method were the dates of start and end of waves, peaks, and valleys. The dates are displayed on graphs and added as a new column in a dataset. We found that Belgium was the country recording the highest ratio of incident cases per 100 people by day in a wave.

**CONCLUSION:** The EpidemicKabu method is simple, easy to use, and very useful in estimating epidemic waves. The methodology requires expert judgment in order to set a parameter that could only have three possible values.

## Introduction

The dynamical behavior of certain epidemic curves often exhibits oscillations, leading to fluctuations between a very low and very high number of incident cases over the time [1,2]. These fluctuations, commonly referred to as waves in the epidemic curve [1,2], signify varying severities of the epidemic and subsequently impact the allocation of resources. The identification of epidemic waves lacks a consensual approach [3,4]. However, since the onset of the COVID-19 pandemic, there have been intensified attempts to systematically define, identify, and predict epidemic waves [5–13].

Previous studies have developed methods to identify epidemic waves based on operational definitions. For instance, Levene [8] defined that within a time series an epidemic wave spans over a period from one valley (minimum) to the next, with a peak (maximum), occurring between them. Zhang et al. [13] proposed that a wave is characterized by some upward and/or downward periods. The increase in an upward period or the decrease in a downward period have to be substantial by sustaining itself over a period of time to distinguish them from an uptick, a downtick, reporting errors, or volatility in new cases. These characteristics are also part of the definition by Srivastava et al. [7]. There are also studies that specifically point out that a rise in the mean number of cases between two consecutive weeks higher than 20% is indicative of an upcoming epidemic wave [14]

Other studies identified epidemic waves by designing a method with an implicit definition [6], or identifying a threshold over and under which a wave starts and ends, respectively [12]. In spite of the different approaches to identify waves, all the previous studies agreed that a complete wave includes a rise in the number of cases, a defined peak, and a decline [4]. Additionally, despite definitional problems, using these descriptive terms has important values for planning and public health [11].

The study of epidemic waves is useful to predict the behavior of future waves (i.e., beginning, duration, and final) and therefore to predict the incidence of new cases [10,12]. It is also useful for retrospective analysis of epidemiological data, such as the estimation of the hospital length of stay through the waves of COVID-19 [15].

One of the main challenges of the epidemic wave identification is distinguishing an upcoming wave from a temporary spike/drop or a group of spikes/drops in infection counts [4,7]. This temporary spike/drop is the noise in the epidemic curve and is due to the demographic and environmental variability [16]. But also due to the quality and timeliness of the surveillance data available [17].

Thus, we describe in this study a new method to identify the start and end of epidemic waves and to demilite their peaks and valleys. We showcased its use using surveillance data from 15 countries worldwide as part of the activities of the project unCoVer, and a Gaussian filter to reduce the noise and avoid the temporary spikes/drops. Python library intended for widespread community utilization.

## Methodology

EpidemicKabu is an original and novel methodology to identify waves, peaks and valleys from an epidemic curve detailed in this paper and available as a Python library at https://pypi.org/project/EpidemicKabu/. The EpidemicKabu name is equivalent to “epidemic curve”, being Kabú the sounds of the word “curve” in Japanese (i.e., カーブ (kābu)). This method allows for the identification of epidemic waves from an recorded time serie of incident cases. The outcome of this development is freely available and accessible for everyone, with an easy way to use, useful to standardize epidemic waves retrospectively. It is based on the overall definition concept of what constitutes an epidemic wave i.e., a rise in the number of cases, a defined peak, and a subsequent decline, as well as the authors’ expert judgment in fitting some model parameters. The methodology follows a series of steps, namely: 1) Smoothening of the epidemic curve (i.e. reduction of noise); 2) Processing of the epidemic curve; 3) Identification of waves; and 4) Delimitation of peaks and valleys. Throughout these steps, the model uses epidemic surveillance data as main input. These could include the daily number of incident cases, prevalent cases, or a daily indicator (e.g., daily incidence rate or attack rate). A demonstration of the EpidemicKabu using real databases and the methodological details are presented in the following chapters.

## 1. Smoothening of the epidemic curve

In order to reduce the noise in the epidemic curve, we initially assumed that the observed curve can be adjusted by interpolation to a theoretical function such as linear, polynomial, or spline [18]. However, these functions do not describe in a correct way the epidemic curve behavior of peaks and valleys. For this reason, we opted to use of a Gaussian kernel to smooth the epidemic curve. This Gaussian kernel is defined by the following function (1):

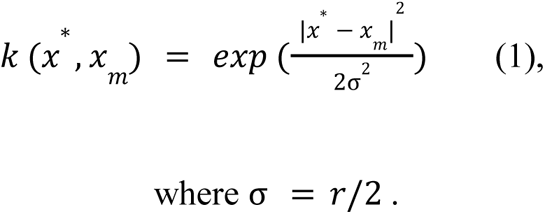

Where σ is the standard deviation, r is the Gaussian kernel radius value, *x*\* are all the temporal serie values or cases, and *x_m_* is the value defined each time to apply the Gaussian kernel. To smooth the curve we followed the following steps to each value in the time series. First, we applied the Gaussian kernel function to the value and obtained the kernel values. Second, we standardized the kernel values to maintain the scale. We propose to standardized them by a normalization with the sum of all the kernel values. Third, we sum all the kernels values normalized and the smoothed value. This method is described in [18] though we adapt it to the discrete data of epidemic curves.

## 2. Processing the curve

A step-by-step process for epidemic curve processing is illustrated in Figure 1. Step one consists of selecting a database with the cases and their date of report. In step two, the case values are normalized by their maximum. The Gaussian smoothing is applied in step three. For this step, the user should set the Gaussian kernel radius value (i.e., *kernel)*. This value depends on the particular curve being processed in such a way that a new curve could require a different *kernel*. We use the next approach to estimate the kernel value (i.e., r):

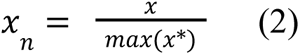

**Fig 1.**
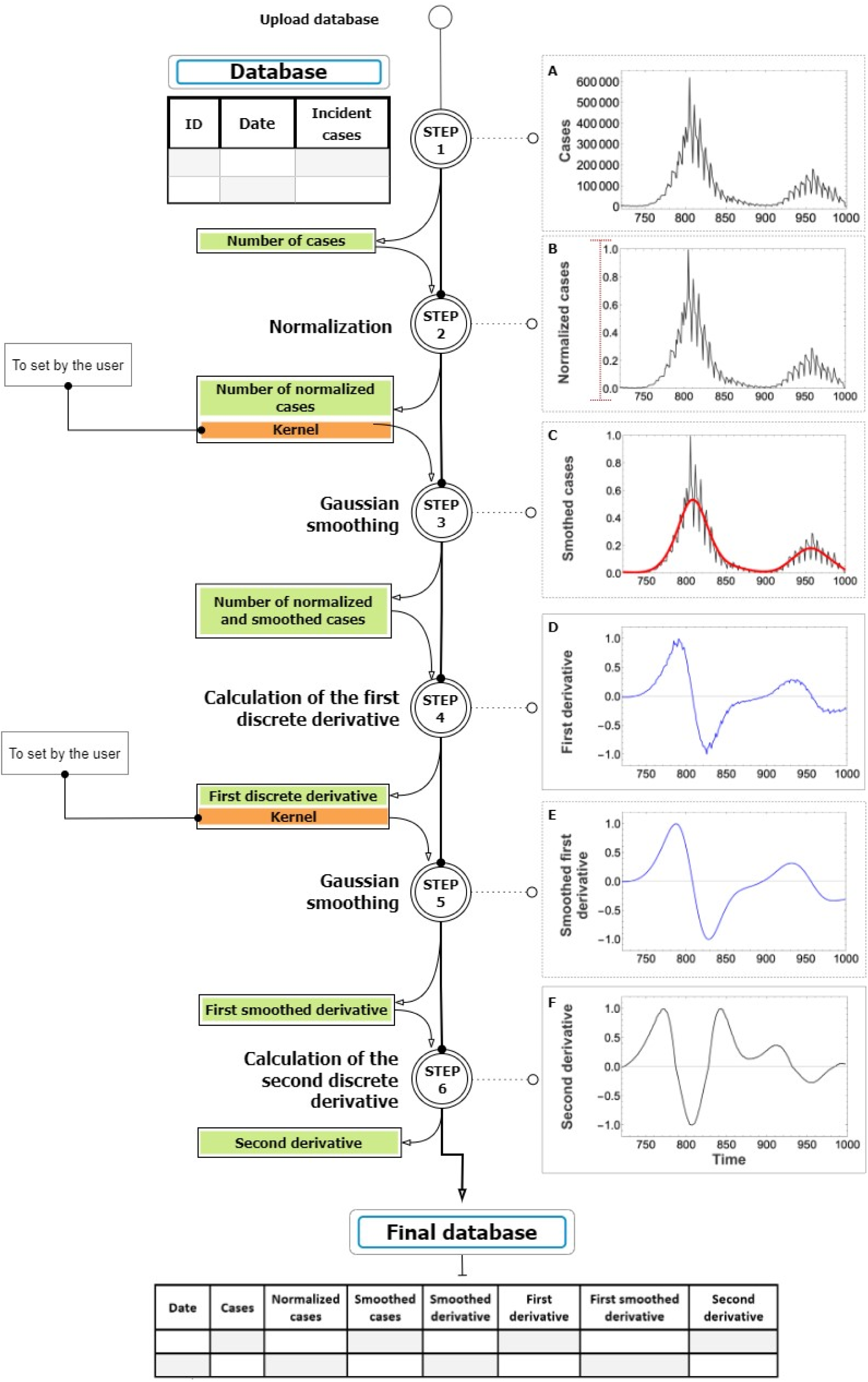
Scheme of the steps to process the epidemic curve. Each step has its input and outputs in green, the parameters to set are in orange, and a graph illustrating the process of each step. **(A)**. The raw epidemic curve in gray, **(B)**. The raw epidemic curve normalized, **(C)**. The smoothed epidemic curve in red, **(D)**. The first derivative in blue, **(E)**. The smoothed first derivative in blue, **(F)**. The second derivative in black. These plots are made with dummy data just to explain the methodology step by step.

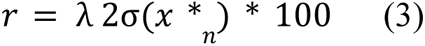

Where *x_n_* is a normalized value of cases, *x* is a value of cases, *x*\* are all the temporal serie values or cases, r is the Gaussian kernel radius value, σ(*x_n_*) is the standard deviation of the normalized cases, and λ is a parameter that could have three possible values. We recommend λ equal to 1, but it also could represent a strong constraint with λ equal to 2 or a weak constraint with λ equal to 0.5. After smoothing the curve, we estimated the first derivative in step four and then smoothen it in step five with the Gaussian kernel. The first derivate allows to identify local minimums when it cuts the x axis going from negative to positive values.

The local minimums delimit waves. Finally, we estimated the second derivative. The second derivate allows to identify the concavity changes or inflexion points which delimit the start and end of a peak or valley. The final database has columns for dates, cases by date, normalized cases, smoothed cases, first derivative values, first derivative smoothed, and second derivative.

## 3. Identification of waves

We used the smoothened first derivative in the final database to detect the waves of the curve (Figure 2). A wave was delimited by the points in which the smoothed first derivative cuts the x-axis passing from negative to positive values. This represents a point between two waves in which the difference in the number of cases between consecutive days is zero. In this way the first step detects the cut points, which are filtered in the second and third steps using the second derivative and a *threshold* parameter that is adjusted by the user. This threshold is by default set to one but could be optionally changed by the user.

**Fig 2.**
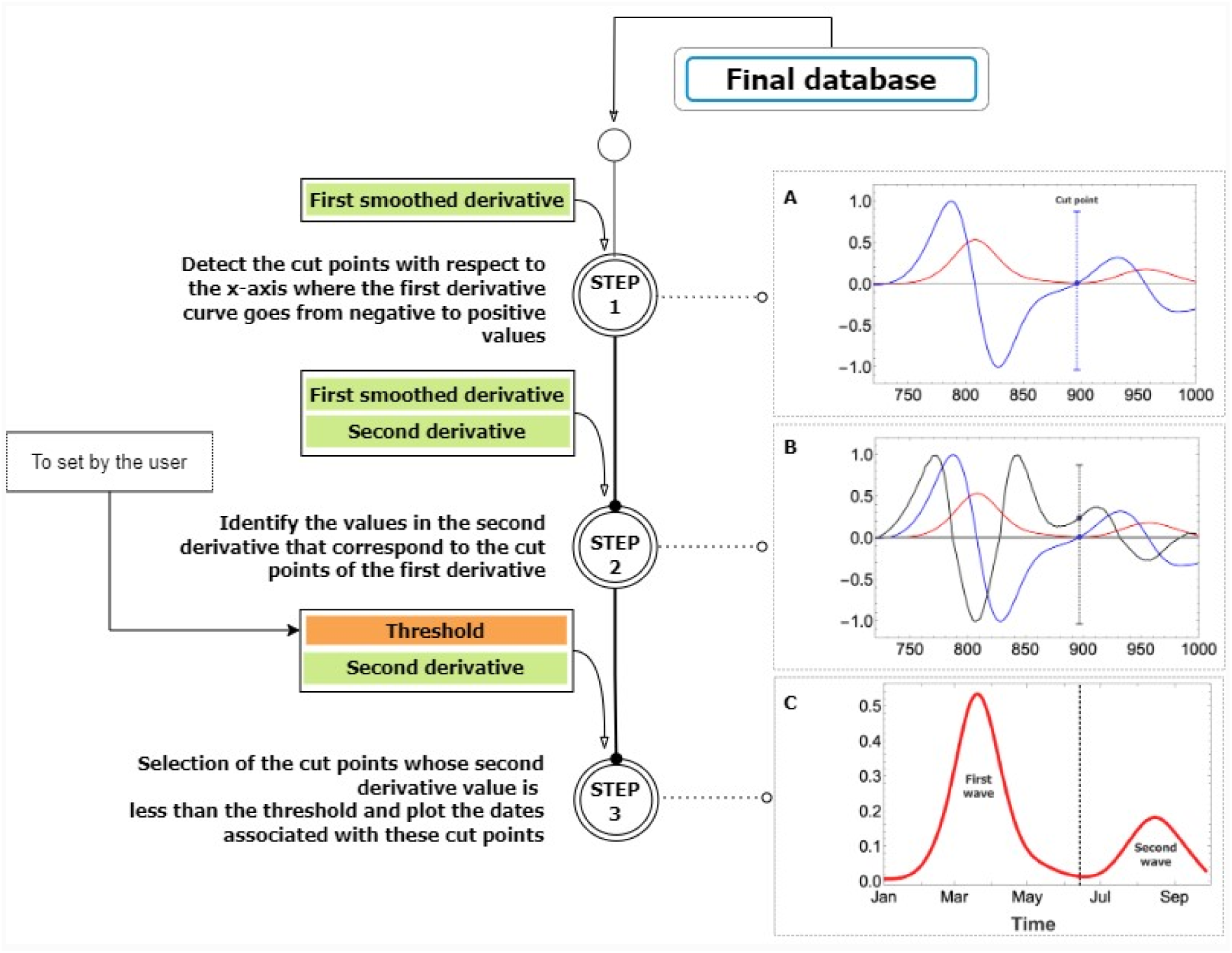
Scheme of the steps to detect the wave. Each step has its input and outputs in green, the parameters to set are in orange, and a graph illustrating the process of each step, **(A)**. The smoothed epidemic curve in red and the first derivative in blue, the cut point is marked on the blue curve, **(B).** The smoothed epidemic curve in red, the first derivative in blue, and the second derivative in black, the point marks on the black curve is the value on the second derivative that corresponds to the position of the cut point in the first derivative **(C)**. The smoothed epidemic curve in red with a dashed line marking the end and start of waves. These plots are made with dummy data just to explain the methodology step by step.

## 4. Detection of peaks and valleys

We used the second derivative in the final database to detect the peaks and valleys of the curve (Figure 3). A peak is delimited by the points in which the second derivative cuts the x-axis passing from positive to negative values and passing from negative to positive values. For each wave, both cut points represent the maximum change in the increase and decrease of cases, respectively. In other words, they represent the maximum difference between consecutive days when the epidemic curve is increasing and when it is decreasing for each wave. The cut points are identified in the first step. In the second and third steps, these cut points are compared with the *maximum point within each wave* (the wave is delimited by the *cut points of the wave* previously identified) from the smoothed and normalized epidemic curve. If there are more than one cut point between the *cut points of the waves* and the *maximum point within the wave*, it is selected the cut point closest to the *maximum point within the wave*.

**Fig 3.**
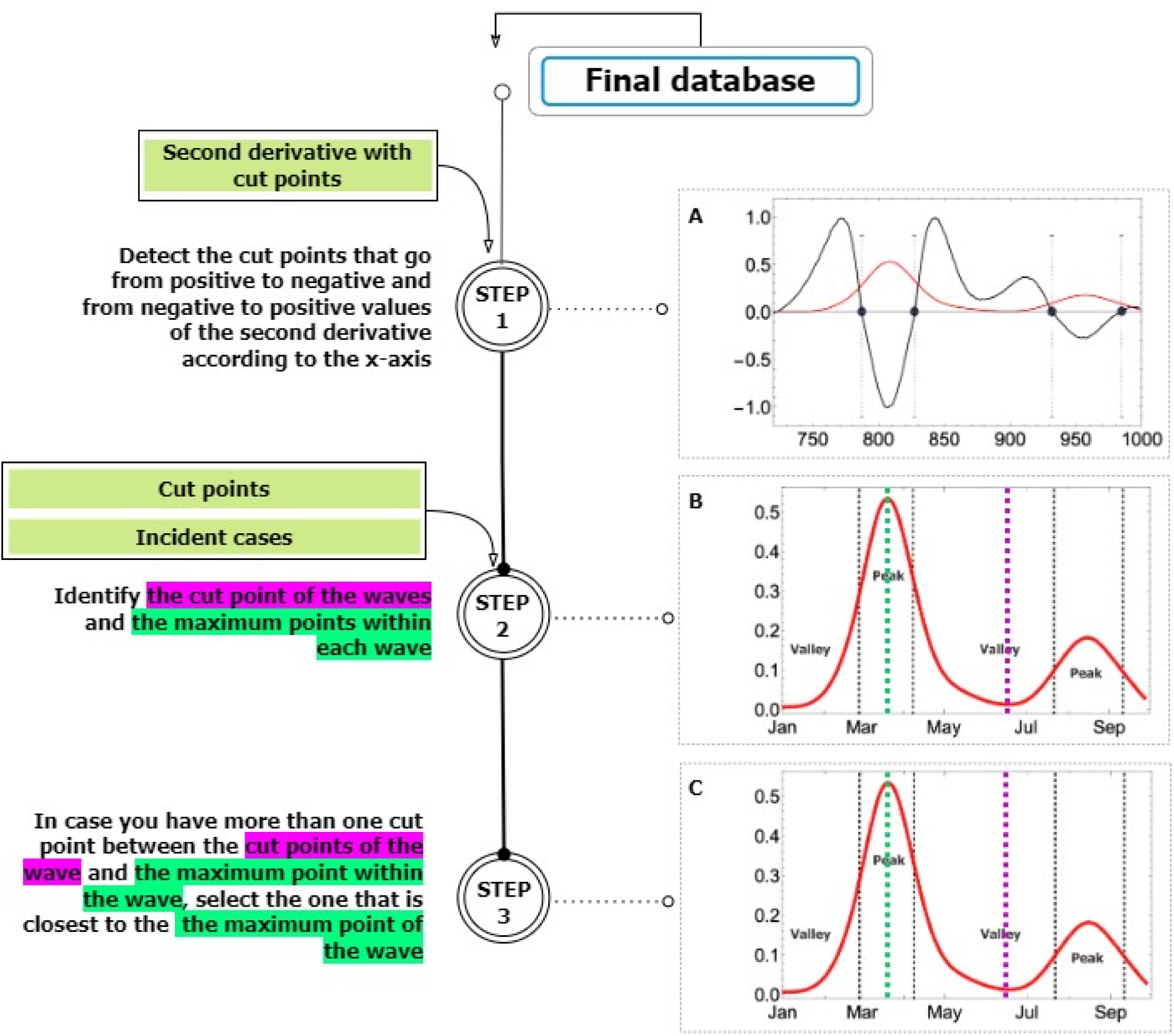
Scheme of the steps to detect peaks and valleys. Each step has its input and outputs in green, the parameters to set are in orange, and a graph illustrating the process of each step, **(A)**. The smoothed epidemic curve in red and the second derivative in black, the cuts point are marked on the black curve, **(B)**. The smoothed epidemic curve in red, the *cut points* of peaks and valleys in black dashed line, the *cut point of the waves* in magenta dashed line,, the *maximum point within a wave* in green dashed line, **(C)**. The smoothed epidemic curve in red, the *cut points* of peaks and valleys in black dashed line, the *cut point of the waves* in magenta dashed line,, the *maximum point within a wave* in green dashed line. These plots are made with dummy data just to explain the methodology step by step.

### Application of EpidemicKabu on multi-country COVID-19 surveillance data

We tested the useability of EpidemicKabu within the activities of the project Unravelling Data for Rapid Evidence-Based Response to COVID-19 network (unCoVer)[19]. unCoVer gathers COVID-19 related data from 15 countries in an attempt to standardize and streamline the use of data for evidence-based insights into the COVID-19 pandemic. The countries represented in unCoVer, and used for the testing phase are: Belgium, Bosnia and Herzegovina, Brazil, Colombia, Croatia, Ireland, Italy, Luxembourg, Norway, Republic of Korea, Romania, Spain, The United Kingdom, United States, and Turkiye. For this, we estimated the waves as well as peaks and valleys of each country’s database with cases reported daily between 2020 and 2022 [20].

### Using EpidemicKabu to estimate the size of the epidemic waves

We also used the already mentioned data to estimate the size of the epidemic waves. For this, we used EpidemicKabu to estimate the waves of each country’s database with daily dates and an indicator estimated by date between 2020 and 2022 to make comparisons between countries. The indicator was the ratio between the daily number of cases and the total size of the population that could get the disease each year multiplied by 100, which is similar to the attack rate indicator [21]. We assume that the total population of each country represents the total population susceptible to the SARS-CoV-2 [22,23]. After identifying the waves with EpidemicKabu for each country, we use the smoothened epidemic curve to calculate four measures related to the wave size: the maximum value of cases by date of each wave (maximum), the sum of the indicator between the start and the end of each wave (total), the number of days between the start and the end of each wave (duration in days), and the ratio between the the total and the duration in days for each wave of each country.

## Results

### EpidemicKabu Library

We built an open access Python library https://pypi.org/project/EpidemicKabu/, https://github.com/LinaMRuizG/EpidemicKabuLibrary that could be used by researchers.

### Testing EpidemicKabu on COVID-19 data

Fig 4A and SI Fig 1-14 show the waves and Fig 4B and SI Fig 15-28 show the peaks and valleys of the COVID-19 epidemic curves between 2020 and 2022 for the 15 countries of interest. For each country, we set the *kernels* for the epidemic curve (i.e., *kernel1*) and its first derivative (i.e., *kernel2*) as stated in SI Table 1. The number of waves varied between 3 (for Norway) and 8 (for Belgium, Luxembourg, and Spain). The mean number of waves by country is 6.06 and a median is 6 (IQR 5.25 –7.0). All the codes of this section and additional versions of these plots are in https://github.com/LinaMRuizG/EpidemicKabuLibrary/tree/main/examples.

**Fig 4.**
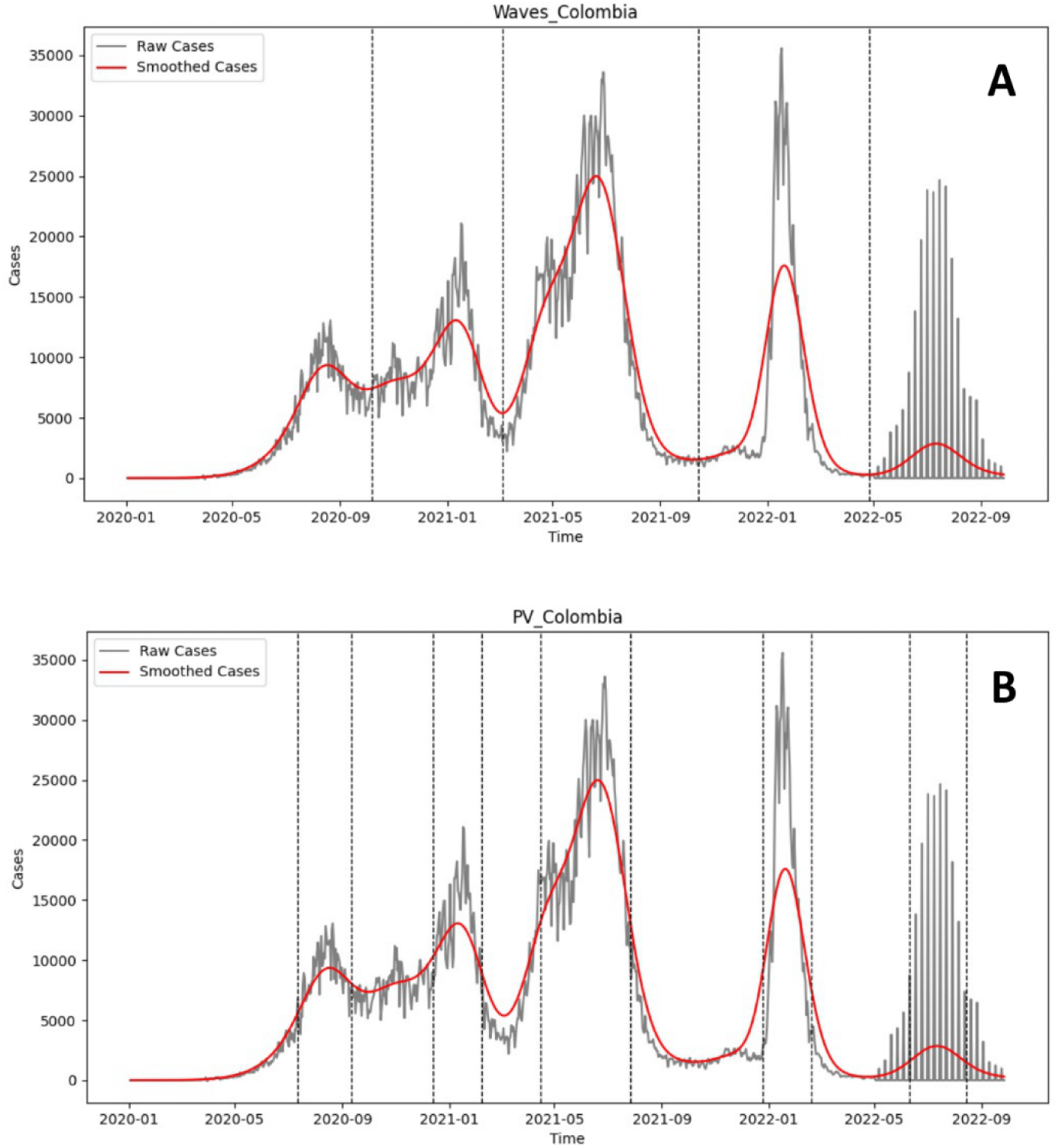
Waves and Peaks and Valleys of Colombia COVID-19 epidemic curve using EpidemicKabu. (**A**) Waves, **(B)** Peaks and valleys. The raw epidemic curve in gray shows the daily incident cases of COVID-19 between 2020 and 2022, the smoothed epidemic curve is in red, and the dashed vertical lines are the waves delimitations. *PV: Peaks and Valleys.

### Using EpidemicKabu to estimate the size of the epidemic waves

Fig 5 shows the waves estimated for Colombia with the curves of the indicator. As expected, these estimated waves are equal to the estimated waves for the epidemic curve with the number of cases by date, and this also happens with the other 14 countries (SI Fig 29-42). For each country, we set the kernels as previously mentioned (SI Table 1). All the codes of this section and additional versions of these plots are in https://github.com/LinaMRuizG/EpidemicKabuLibrary/tree/main/examples.

**Fig 5.**
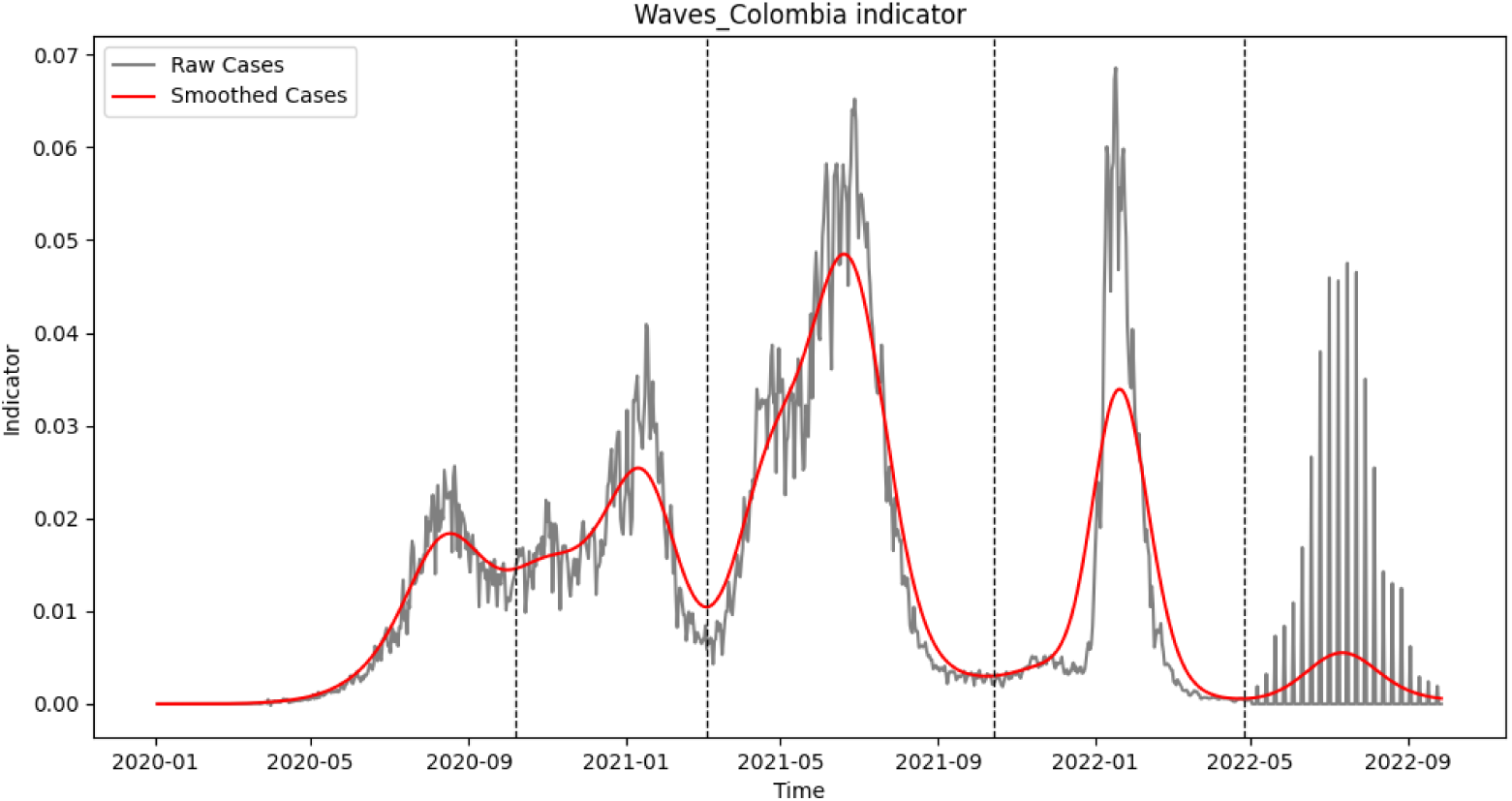
Waves of Colombia COVID-19 epidemic curve with an indicator using EpidemicKabu. The raw epidemic curve in gray shows the daily indicator, the smoothed epidemic curve is in red, and the dashed vertical lines are the waves delimitations. Additional plots in (plots with the indicator) https://github.com/LinaMRuizG/EpidemicKabuLibrary/tree/main/examples/plots.

Fig 6 shows the distribution of the measures related to the wave size for each country. Specifically, Fig 6A shows a boxplot for the maximum value of incident cases reached in each wave of all the waves of each country. From this is possible to identify that, Republic of South Korea is the country with both the lowest and the highest maximum value of incident cases reached in a wave (i.e., 0.0003 and 0.45 cases by 100 people, respectively). This country also has the highest mean and standard deviation (i.e., 0.16 and 0.21, respectively) from all the maximum values of each wave. Colombia is the country with the lowest mean (i.e., 0.024 cases by 100 people), and Brazil has the lowest standard deviation of 0.01.

**Fig 6.**
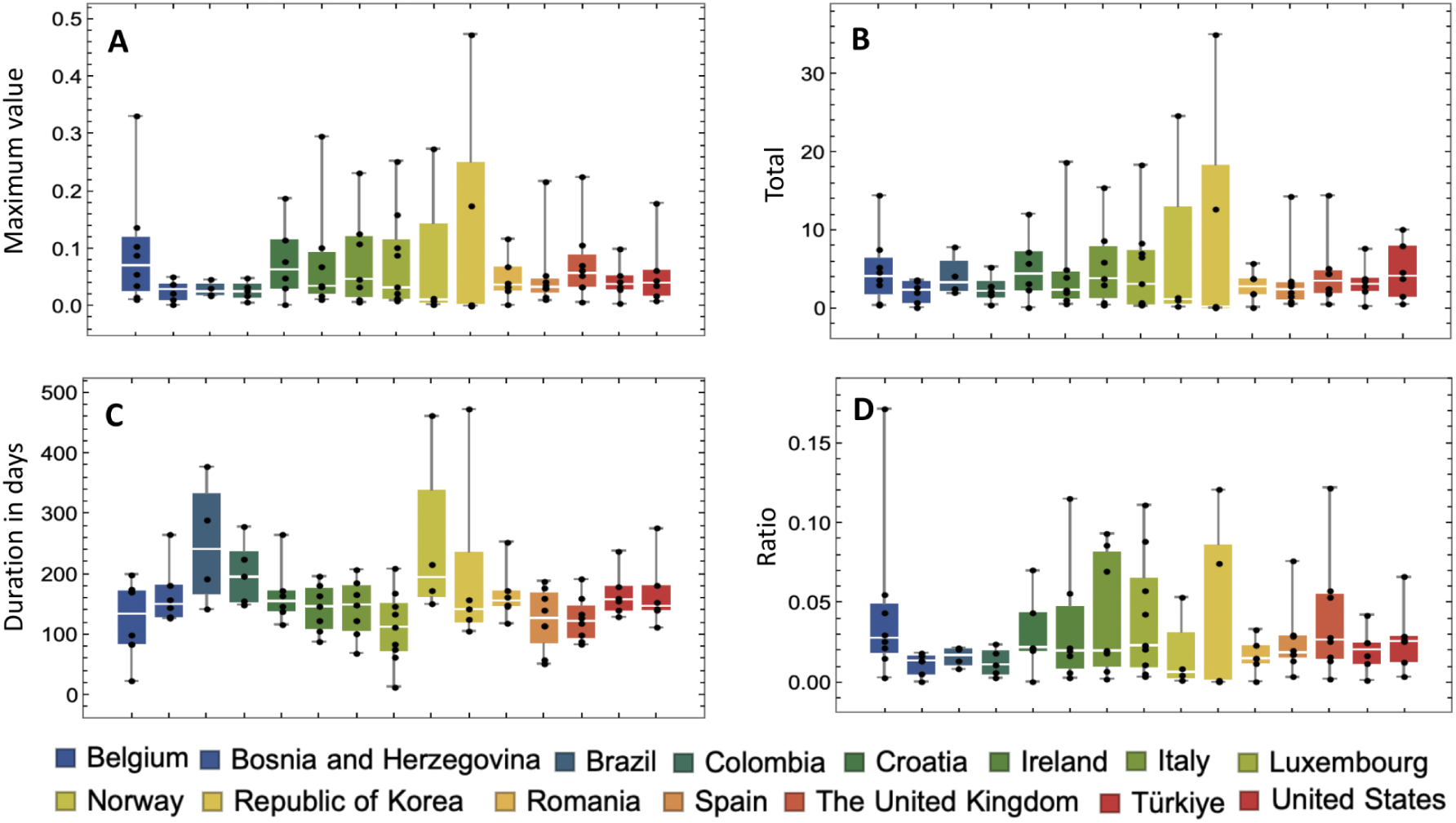
Measures for the COVID-19 waves of all countries using the indicator of daily incident cases. (**A**). Boxplots of the maximum value of the indicator for each wave of each country, (**B).** Boxplots of the sum of the indicator between the start and the end of the wave for each wave of each country, (**C).** Boxplots of the number of days between the start and the end of the wave for each wave of each country, (**D).** Boxplots of the ratio between the Total and the Duration in days for each wave of each country. *The bottom shows the countries names from left to right and top to bottom order as they appear in the plots.

Fig 6B shows a boxplot of the sum of the indicator between the start and the end of a wave for each wave of each country. The Republic of South Korea is the country with both the lowest and the highest total number of incident cases by 100 people by wave (i.e., 0.02 and 35.06, respectively), it also have the highest mean 11.9 and standard deviation 16.5 for the total number of incident cases by 100 people by wave of its set of waves. Bosnia and Herzegovina is the country with the lowest mean (i.e., 2.03 cases by 100 people in total by wave) and the lowest standard deviation of 1.44.

Fig 6C shows boxplots of the number of days between the start and the end of the wave for each wave of each country. Luxembourg has the wave with lowest duration (i.e., 11 days) and also one of the lowest means (i.e., 123 days). Republic of Korea had the wave with the highest duration (i.e., 472 days) and the highest standard deviation of the set of waves by country (i.e., 166.2 days by wave). Norway have the highest mean of duration of the set of waves by country (i.e., 333.7 days by wave). Finally, the United Kingdom has the lowest standard deviation of 36.88.

Fig 6D shows boxplots of the ratio between the total number of incident cases by wave and the duration in days for each wave of each country. The countries with the lowest and highest ratio are Republic of South Korea (i.e., 0.00015) and Belgium (i.e., 0.17), respectively. The countries with the lowest and highest mean of the ratios by set of waves are Bosnia and Herzegovina (i.e., 0.01) and Belgium (i.e., 0.05), respectively. Finally, the countries with the lowest and highest standard deviations of the set of ratios for each wave are Brazil (i.e., 0.006) and Republic of Korea (i.e., 0.06), respectively.

SI Fig 43 shows the same measures already described for the set of waves of each country but in comparison with the set of waves of all countries at once. For the maximum value by wave, the lowest value is 0.0003, the highest value is 0.45, the mean is 0.06, and the standard deviation is 0.08. For the total value by wave, the lowest value is 0.02, the highest value is 35.06, the mean is 4.71, and the standard deviation is 6.24. For the duration in days, the lowest value is 11, the highest value is 472, the mean is 164.67, and the standard deviation is 78.7. For the ratio, the lowest value is 0.0001, the highest value is 0.17, the mean is 0.027, and the standard deviation is 0.03.

## Discussion

One of the main challenges of the identification of epidemic waves is distinguishing an upcoming wave from a temporary spike/drop in infection counts [4,7], as this noise affects the estimation of the waves. We used a discrete version of a Gaussian filter to avoid it, which allows for a more accurate identification of the waves and their peaks and valleys. This is a more nuanced approach than rolling the average mean, which is the standard practice in previous studies [5,11]. Additionally, unlike previous studies, we included the smoothening the first derivative reducing additional noise and improving the performance of the wave detection.

The epidemic curve has a significant amount of noise because of the inherent stochasticity of the transmission, recovery, births, and deaths (i.e., demographic variability), and the environment impacts on the epidemic outcomes (i.e., environmental variability) [16]. Additionally, noise is introduced due to the variable quality and timeliness of the surveillance data available, such as under-reporting and reporting delays, respectively[17]. Under-reporting causes some fraction of infections to never be reported, while delays redistribute reports of infections incorrectly across time [17].

Sometimes, as in the case of the United Kingdom curve, the waves detected by this methodology appear not to be in accordance with the statement: a wave requires substantial, significant and sustained periods of increase [11], and it ends when the virus is brought under control and the cases fall substantially or they remain stable over time [13]. However, this could be improved by increasing the kernel value for smoothing the epidemic curve. Here is when the expert judgment comes to filter the identified waves. Future work will diminish the subjectivity of this parameter.

Our methodology has two parameters to be set but the user and a third one that is optional. Similarly, previous methods have one or more than one parameters to be set by the user [11,13]. We noticed that although epidemicKabu does not set the minimum duration of a wave as in the methodology designed by Harvety et al. (2022) [11], the duration in days of our detected waves are in the range stablished by this method. Excluding the last waves in Belgium and Bosnia-Herzegovina. However these waves are the last one in their time series and they are incomplete. According to Harvety et al. (2022) [11], the minimum duration of a wave can be informed by biological information on the transmission of the disease; for COVID-19 they set 35 days [11].

There are previous studies that systematically define, identify, and predict epidemic waves [5–13] but the methodology in less standardized. Our methodology is easy to apply, and aiming to be a globally used method that allows comparative studies and to have a common language to use. On the other hand, as for previous methodology [24], we strongly recommend EpidemicKabu methodology to be used on sufficient historical data. In such way that the expert judgment could consider the transmission dynamics and select a kernel that deletes the noise under the minimum duration time of the wave.

The estimation of waves is very useful for retrospective analysis of epidemiological data. In the case of the COVID-19 pandemic, there are several studies investigating the characteristics of the waves: identification of pronounced changes of lifestyle habits throughout the waves [25], other reviewed changes in dream [26]; overview of technologies implemented during the first wave of the COVID-19 pandemic [27]; identification of the existence of strong external drivers (i.e., temperature and absolute humidity) of transmission intensity throughout waves [28]; comparison of epidemiologic features such as demographics, transmission chains, case fatality rates, social activity levels and public health responses [29]; sociodemographics, source population at risk factors, transmission places, and case frequency [30]; identification of differences in patients’ profiles [31]. However, some of these studies do not used a clear and systematic methodology to define the studied waves.

As expected, each identified wave has a peak in the middle of two valleys which are shared between consecutive waves. The critical parameters of the algorithm are the kernels used to smooth both the epidemic curve and its first derivative. This parameter should be set by the user and its magnitude is already exemplified in the previous sessions. Though the user could explore different values for both kernel, we advise to set them as equal as in the examples. Future work could test the library with COVID-19 data for other countries or even other epidemics.

To our knowledge, there are no previous studies that measure the size of the epidemic waves. We found that although Republic of South Korea is the country recording the highest maximum value (i.e. the height of the wave or the highest point in the wave) and the highest total number of incident cases by 100 people by wave, it is not the country with the highest ratio of incident cases by 100 people by day because its waves have a long duration. However, Belgium is the country with the highest ratio because its waves have a short duration, and also the country with the highest ratio mean over all its recorded waves. Future research could compare this results with those for other epidemics.

## Conclusion

EpidemicKabu is an open-access library in Python to estimate waves from epidemic curves and their peaks an valleys. It is built with the aim to be a globally used methodology by researchers and in this way to allow comparison between studies that analyze characteristics of epidemic waves. It face the noise in the epidemic curve with a Gaussian smoothing of the curves, for which the user should set the kernel parameter.

## Data Availability

All the code can be accessed at https://github.com/LinaMRuizG/EpidemicKabuLibrary and https://pypi.org/project/EpidemicKabu/. All authors had full access to the study data and accepted the responsibility to submit for publication

## Acknowledgements

We give thanks to the European Union’s Horizon 2020 Research and Innovation Programme (unCoVer, Grant Agreement No 101016216), and the ReFReCA project which is financed by the Ministry of Science, Technology and Innovation of Colombia – Minciencias (code 111584467754).

## Supplementary Information

**SI Fig 1. Waves of Belgium COVID-19 epidemic curve using EpidemicKabu.** The raw epidemic curve in gray shows the daily incident cases of COVID-19 between 2020 and 2022, the smoothed epidemic curve is in red, and the dashed vertical lines are the waves delimitations.

**SI Fig 2. Waves of Bosnia and Herzegovina COVID-19 epidemic curve using EpidemicKabu.** The raw epidemic curve in gray shows the daily incident cases of COVID-19 between 2020 and 2022, the smoothed epidemic curve is in red, and the dashed vertical lines are the waves delimitations.

**SI Fig 3. Waves of Brazil COVID-19 epidemic curve using EpidemicKabu.** The raw epidemic curve in gray shows the daily incident cases of COVID-19 between 2020 and 2022, the smoothed epidemic curve is in red, and the dashed vertical lines are the waves delimitations.

**SI Fig 4. Waves of Croatia COVID-19 epidemic curve using EpidemicKabu.** The raw epidemic curve in gray shows the daily incident cases of COVID-19 between 2020 and 2022, the smoothed epidemic curve is in red, and the dashed vertical lines are the waves delimitations.

**SI Fig 5. Waves of Ireland COVID-19 epidemic curve using EpidemicKabu.** The raw epidemic curve in gray shows the daily incident cases of COVID-19 between 2020 and 2022, the smoothed epidemic curve is in red, and the dashed vertical lines are the waves delimitations.

**SI Fig 6. Waves of Italy COVID-19 epidemic curve using EpidemicKabu.** The raw epidemic curve in gray shows the daily incident cases of COVID-19 between 2020 and 2022, the smoothed epidemic curve is in red, and the dashed vertical lines are the waves delimitations.

**SI Fig 7. Waves of Luxembourg COVID-19 epidemic curve using EpidemicKabu.** The raw epidemic curve in gray shows the daily incident cases of COVID-19 between 2020 and 2022, the smoothed epidemic curve is in red, and the dashed vertical lines are the waves delimitations.

**SI Fig 8. Waves of Norway COVID-19 epidemic curve using EpidemicKabu.** The raw epidemic curve in gray shows the daily incident cases of COVID-19 between 2020 and 2022, the smoothed epidemic curve is in red, and the dashed vertical lines are the waves delimitations.

**SI Fig 9. Waves of Republic of Korea COVID-19 epidemic curve using EpidemicKabu.** The raw epidemic curve in gray shows the daily incident cases of COVID-19 between 2020 and 2022, the smoothed epidemic curve is in red, and the dashed vertical lines are the waves delimitations.

**SI Fig 10. Waves of Romania COVID-19 epidemic curve using EpidemicKabu.** The raw epidemic curve in gray shows the daily incident cases of COVID-19 between 2020 and 2022, the smoothed epidemic curve is in red, and the dashed vertical lines are the waves delimitations.

**SI Fig 11. Waves of Spain COVID-19 epidemic curve using EpidemicKabu.** The raw epidemic curve in gray shows the daily incident cases of COVID-19 between 2020 and 2022, the smoothed epidemic curve is in red, and the dashed vertical lines are the waves delimitations.

**SI Fig 12. Waves of United Kingdom COVID-19 epidemic curve using EpidemicKabu.** The raw epidemic curve in gray shows the daily incident cases of COVID-19 between 2020 and 2022, the smoothed epidemic curve is in red, and the dashed vertical lines are the waves delimitations.

**SI Fig 13. Waves of Türkiye COVID-19 epidemic curve using EpidemicKabu.** The raw epidemic curve in gray shows the daily incident cases of COVID-19 between 2020 and 2022, the smoothed epidemic curve is in red, and the dashed vertical lines are the waves delimitations.

**SI Fig 14. Waves of United States COVID-19 epidemic curve using EpidemicKabu.** The raw epidemic curve in gray shows the daily incident cases of COVID-19 between 2020 and 2022, the smoothed epidemic curve is in red, and the dashed vertical lines are the waves delimitations.

**SI Fig 15. Peaks and Valleys of Belgium COVID-19 epidemic curve using EpidemicKabu.** The raw epidemic curve in gray shows the daily incident cases of COVID-19 between 2020 and 2022, the smoothed epidemic curve is in red, and the dashed vertical lines are the peaks and valleys delimitations. *PV: Peaks and Valleys.

**SI Fig 16. Peaks and Valleys of Bosnia and Herzegovina COVID-19 epidemic curve using EpidemicKabu.** The raw epidemic curve in gray shows the daily incident cases of COVID-19 between 2020 and 2022, the smoothed epidemic curve is in red, and the dashed vertical lines are the peaks and valleys delimitations. *PV: Peaks

**SI Fig 17. Peaks and Valleys of Brazil COVID-19 epidemic curve using EpidemicKabu.** The raw epidemic curve in gray shows the daily incident cases of COVID-19 between 2020 and 2022, the smoothed epidemic curve is in red, and the dashed vertical lines are the peaks and valleys delimitations. *PV: Peaks and Valleys.

**SI Fig 18. Peaks and Valleys of Croatia COVID-19 epidemic curve using EpidemicKabu.** The raw epidemic curve in gray shows the daily incident cases of COVID-19 between 2020 and 2022, the smoothed epidemic curve is in red, and the dashed vertical lines are the peaks and valleys delimitations. *PV: Peaks and Valleys.

**SI Fig 19. Peaks and Valleys of Ireland COVID-19 epidemic curve using EpidemicKabu.** The raw epidemic curve in gray shows the daily incident cases of COVID-19 between 2020 and 2022, the smoothed epidemic curve is in red, and the dashed vertical lines are the peaks and valleys delimitations. *PV: Peaks and Valleys.

**SI Fig 20. Peaks and Valleys of Italy COVID-19 epidemic curve using EpidemicKabu.** The raw epidemic curve in gray shows the daily incident cases of COVID-19 between 2020 and 2022, the smoothed epidemic curve is in red, and the dashed vertical lines are the peaks and valleys delimitations. *PV: Peaks and Valleys.

**SI Fig 21. Peaks and Valleys of Luxembourg COVID-19 epidemic curve using EpidemicKabu.** The raw epidemic curve in gray shows the daily incident cases of COVID-19 between 2020 and 2022, the smoothed epidemic curve is in red, and the dashed vertical lines are the peaks and valleys delimitations. *PV: Peaks and Valleys.

**SI Fig 22. Peaks and Valleys of Norway COVID-19 epidemic curve using EpidemicKabu.** The raw epidemic curve in gray shows the daily incident cases of COVID-19 between 2020 and 2022, the smoothed epidemic curve is in red, and the dashed vertical lines are the peaks and valleys delimitations. *PV: Peaks and Valleys.

**SI Fig 23. Peaks and Valleys of Republic of Korea COVID-19 epidemic curve using EpidemicKabu.** The raw epidemic curve in gray shows the daily incident cases of COVID-19 between 2020 and 2022, the smoothed epidemic curve is in red, and the dashed vertical lines are the peaks and valleys delimitations. *PV: Peaks and Valleys.

**SI Fig 24. Peaks and Valleys of Romania COVID-19 epidemic curve using EpidemicKabu.** The raw epidemic curve in gray shows the daily incident cases of COVID-19 between 2020 and 2022, the smoothed epidemic curve is in red, and the dashed vertical lines are the peaks and valleys delimitations. *PV: Peaks and Valleys.

**SI Fig 25. Peaks and Valleys of Spain COVID-19 epidemic curve using EpidemicKabu.** The raw epidemic curve in gray shows the daily incident cases of COVID-19 between 2020 and 2022, the smoothed epidemic curve is in red, and the dashed vertical lines are the peaks and valleys delimitations. *PV: Peaks and Valleys.

**SI Fig 26. Peaks and Valleys of United Kingdom COVID-19 epidemic curve using EpidemicKabu.** The raw epidemic curve in gray shows the daily incident cases of COVID-19 between 2020 and 2022, the smoothed epidemic curve is in red, and the dashed vertical lines are the peaks and valleys delimitations. *PV: Peaks and Valleys.

**SI Fig 27. Peaks and Valleys of Türkiye COVID-19 epidemic curve using EpidemicKabu.** The raw epidemic curve in gray shows the daily incident cases of COVID-19 between 2020 and 2022, the smoothed epidemic curve is in red, and the dashed vertical lines are the peaks and valleys delimitations. *PV: Peaks and Valleys.

**SI Fig 28. Peaks and Valleys of United States COVID-19 epidemic curve using EpidemicKabu.** The raw epidemic curve in gray shows the daily incident cases of COVID-19 between 2020 and 2022, the smoothed epidemic curve is in red, and the dashed vertical lines are the peaks and valleys delimitations. *PV: Peaks and Valleys.

**S1 Table 1. Configuration table.** It shows for each country the kernel1 to smooth epidemic curve and kernel2 to smooth its first derivative. **λ** refers if the used kernel was the recommended or the strong or weak constraint. A complete file with kernels for other countries is in https://github.com/LinaMRuizG/EpidemicKabuLibrary/blob/main/examples/data/configurationFile.csv

**SI Fig 29. Waves of Belgium COVID-19 epidemic curve with an indicator.** The raw epidemic curve in gray shows the daily indicator, the smoothed epidemic curve is in red, and the dashed vertical lines are the waves delimitations.

**SI Fig 30. Waves of Bosnia and Herzegovina COVID-19 epidemic curve with an indicator.** The raw epidemic curve in gray shows the daily indicator, the smoothed epidemic curve is in red, and the dashed vertical lines are the waves delimitations.

**SI Fig 31. Waves of Brazil COVID-19 epidemic curve with an indicator.** The raw epidemic curve in gray shows the daily indicator, the smoothed epidemic curve is in red, and the dashed vertical lines are the waves delimitations.

**SI Fig 32. Waves of Croatia COVID-19 epidemic curve with an indicator.** The raw epidemic curve in gray shows the daily indicator, the smoothed epidemic curve is in red, and the dashed vertical lines are the waves delimitations.

**SI Fig 33. Waves of Ireland COVID-19 epidemic curve with an indicator.** The raw epidemic curve in gray shows the daily indicator, the smoothed epidemic curve is in red, and the dashed vertical lines are the waves delimitations.

**SI Fig 34. Waves of Italy COVID-19 epidemic curve with an indicator.** The raw epidemic curve in gray shows the daily indicator, the smoothed epidemic curve is in red, and the dashed vertical lines are the waves delimitations.

**SI Fig 35. Waves of Luxembourg COVID-19 epidemic curve with an indicator.** The raw epidemic curve in gray shows the daily indicator, the smoothed epidemic curve is in red, and the dashed vertical lines are the waves delimitations.

**SI Fig 36. Waves of Norway COVID-19 epidemic curve with an indicator.** The raw epidemic curve in gray shows the daily indicator, the smoothed epidemic curve is in red, and the dashed vertical lines are the waves delimitations.

**SI Fig 37. Waves of Republic of Korea COVID-19 epidemic curve with an indicator.** The raw epidemic curve in gray shows the daily indicator, the smoothed epidemic curve is in red, and the dashed vertical lines are the waves delimitations.

**SI Fig 38. Waves of Romania COVID-19 epidemic curve with an indicator.** The raw epidemic curve in gray shows the daily indicator, the smoothed epidemic curve is in red, and the dashed vertical lines are the waves delimitations.

**SI Fig 39. Waves of Spain COVID-19 epidemic curve with an indicator.** The raw epidemic curve in gray shows the daily indicator, the smoothed epidemic curve is in red, and the dashed vertical lines are the waves delimitations.

**SI Fig 40. Waves of United Kingdom COVID-19 epidemic curve with an indicator.** The raw epidemic curve in gray shows the daily indicator, the smoothed epidemic curve is in red, and the dashed vertical lines are the waves delimitations.

**SI Fig 41. Waves of Colombia COVID-19 epidemic curve with an indicator.** The raw epidemic curve in gray shows the daily indicator, the smoothed epidemic curve is in red, and the dashed vertical lines are the waves delimitations.

**SI Fig 42. Waves of United States COVID-19 epidemic curve with an indicator.** The raw epidemic curve in gray shows the daily indicator, the smoothed epidemic curve is in red, and the dashed vertical lines are the waves delimitations.

