## SupplementaryInformation for "EpidemicKabu a new method to identify epidemic waves and their peaks and valleys": SupplementaryInformation.pdf

Supplementary Information: **Kabu a new method to identify epidemic waves and their peaks and valleys**

### Waves

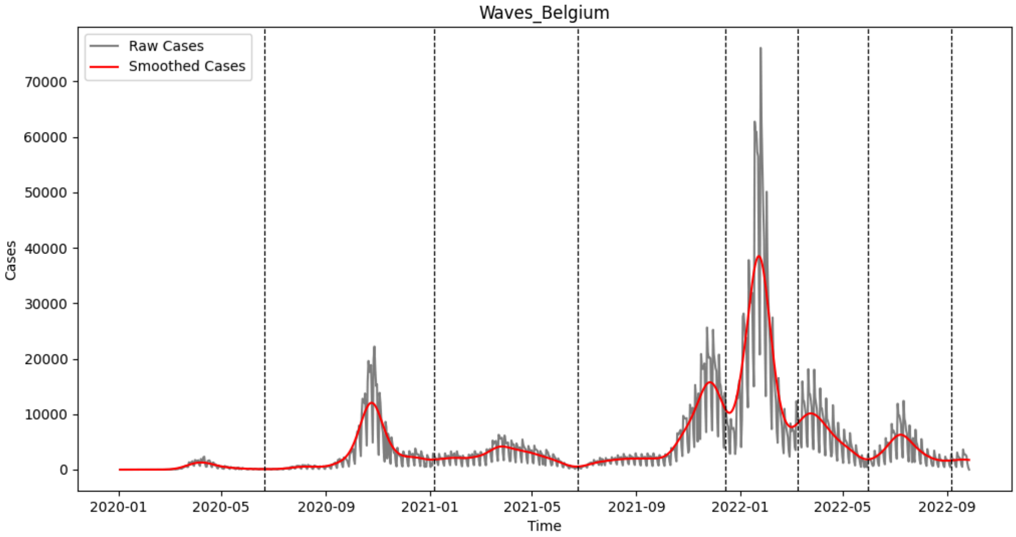

**SI Fig 1. Waves of Belgium COVID-19 epidemic curve using EpidemicKabu.** The raw epidemic curve in gray shows the daily incident cases of COVID-19 between 2020 and 2022, the smoothed epidemic curve is in red, and the dashed vertical lines are the waves delimitations.

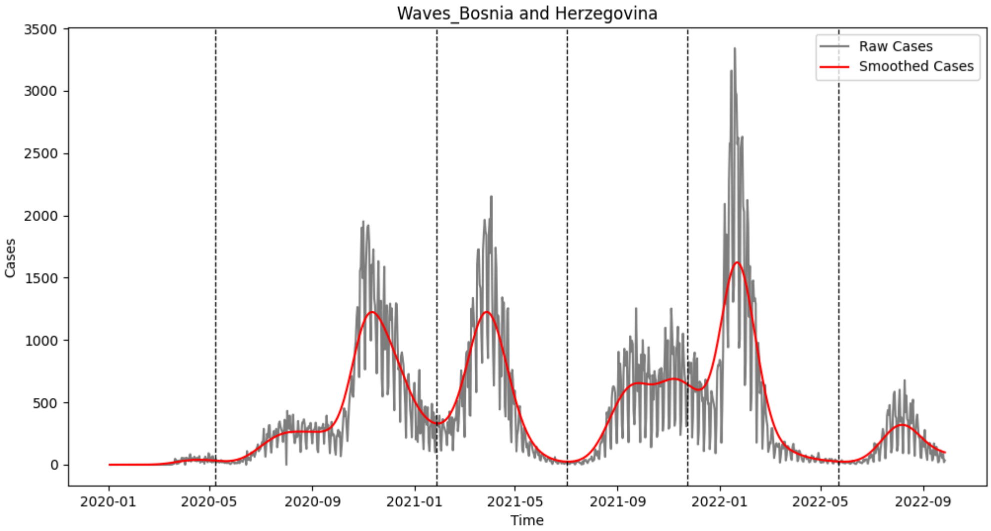

**SI Fig 2. Waves of Bosnia and Herzegovina COVID-19 epidemic curve using EpidemicKabu.** The raw epidemic curve in gray shows the daily incident cases of COVID-19 between 2020 and 2022, the smoothed epidemic curve is in red, and the dashed vertical lines are the waves delimitations.

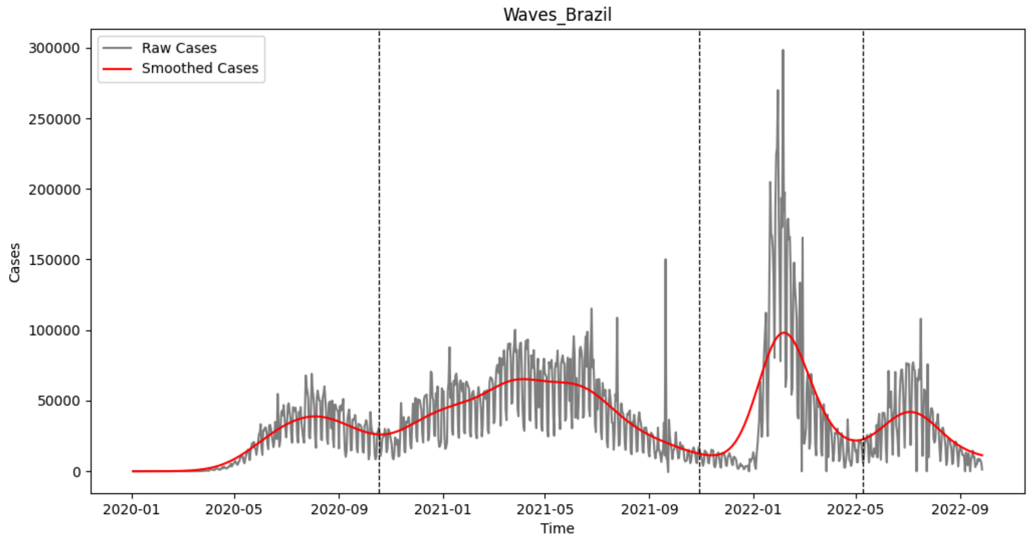

**SI Fig 3. Waves of Brazil COVID-19 epidemic curve using EpidemicKabu.** The raw epidemic curve in gray shows the daily incident cases of COVID-19 between 2020 and 2022, the smoothed epidemic curve is in red, and the dashed vertical lines are the waves delimitations.

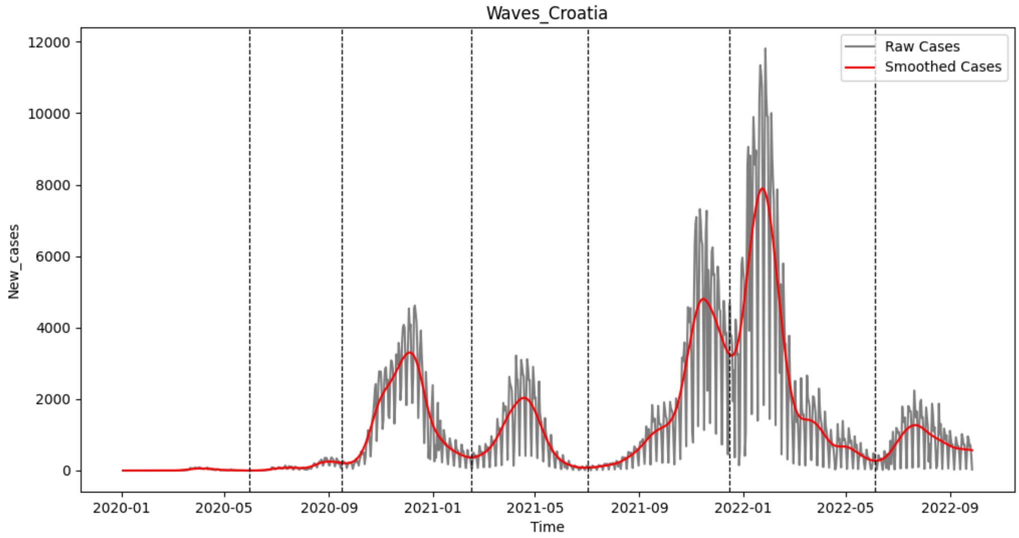

**SI Fig 4. Waves of Croatia COVID-19 epidemic curve using EpidemicKabu.** The raw epidemic curve in gray shows the daily incident cases of COVID-19 between 2020 and 2022, the smoothed epidemic curve is in red, and the dashed vertical lines are the waves delimitations.

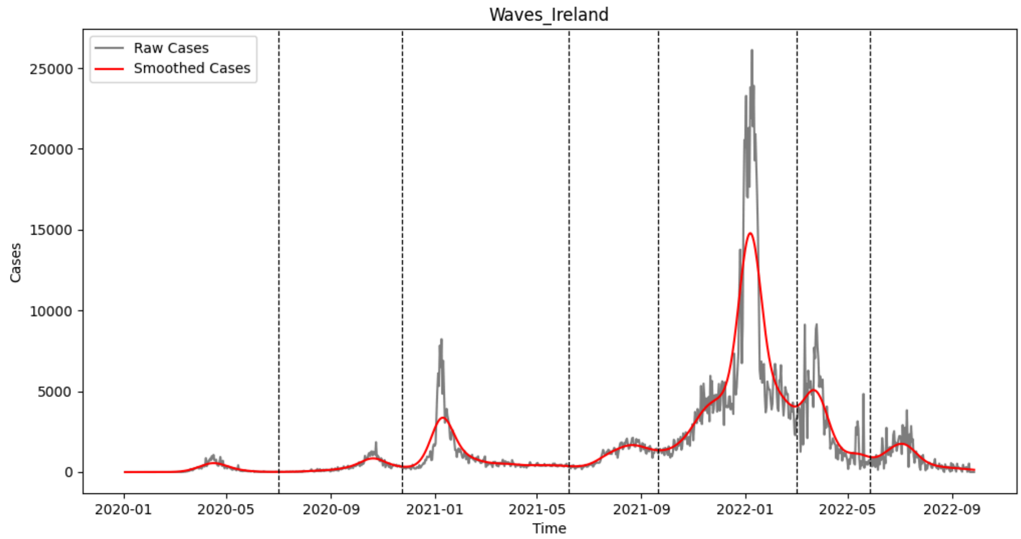

**SI Fig 5. Waves of Ireland COVID-19 epidemic curve using EpidemicKabu.** The raw epidemic curve in gray shows the daily incident cases of COVID-19 between 2020 and 2022, the smoothed epidemic curve is in red, and the dashed vertical lines are the waves delimitations.

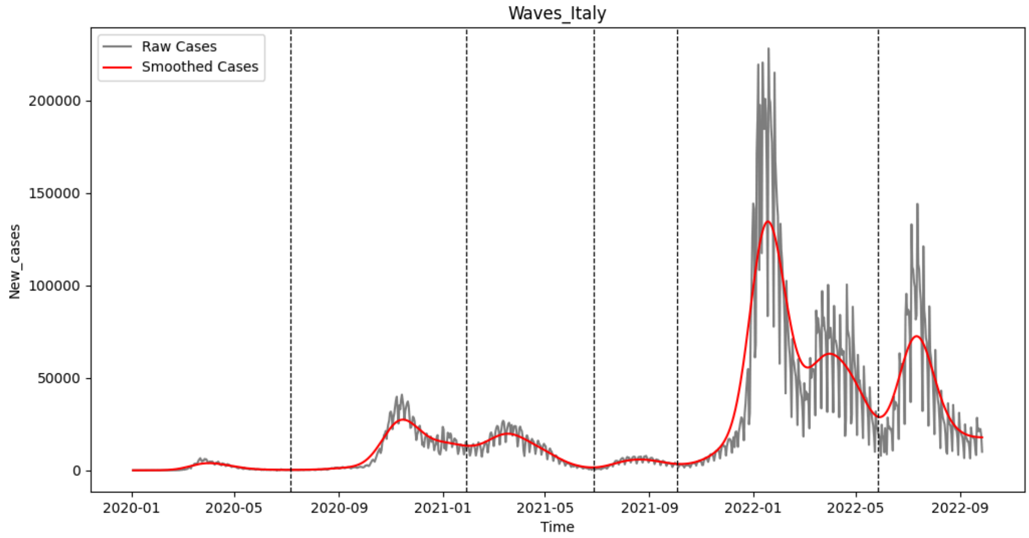

**SI Fig 6. Waves of Italy COVID-19 epidemic curve using EpidemicKabu.** The raw epidemic curve in gray shows the daily incident cases of COVID-19 between 2020 and 2022, the smoothed epidemic curve is in red, and the dashed vertical lines are the waves delimitations.

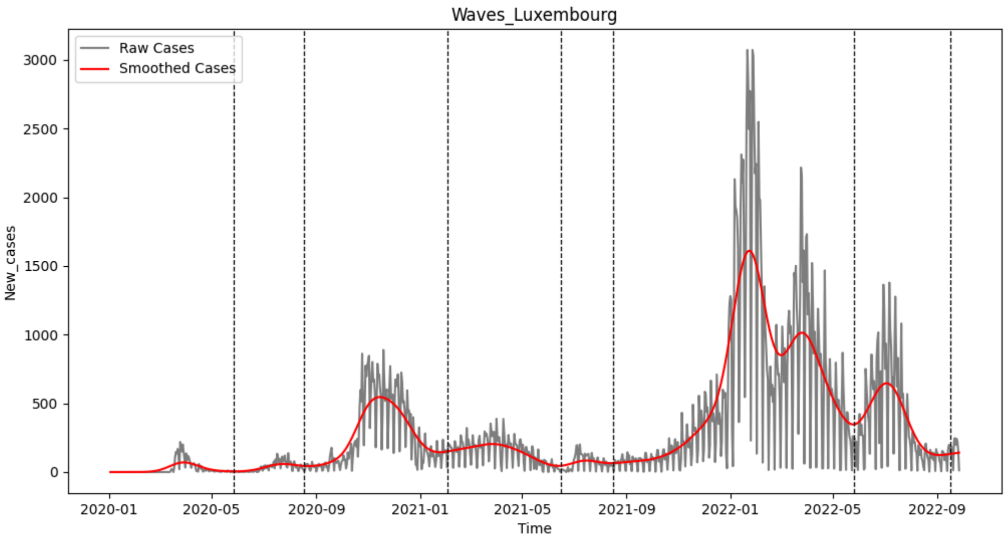

**SI Fig 7. Waves of Luxembourg COVID-19 epidemic curve using EpidemicKabu.** The raw epidemic curve in gray shows the daily incident cases of COVID-19 between 2020 and 2022, the smoothed epidemic curve is in red, and the dashed vertical lines are the waves delimitations.

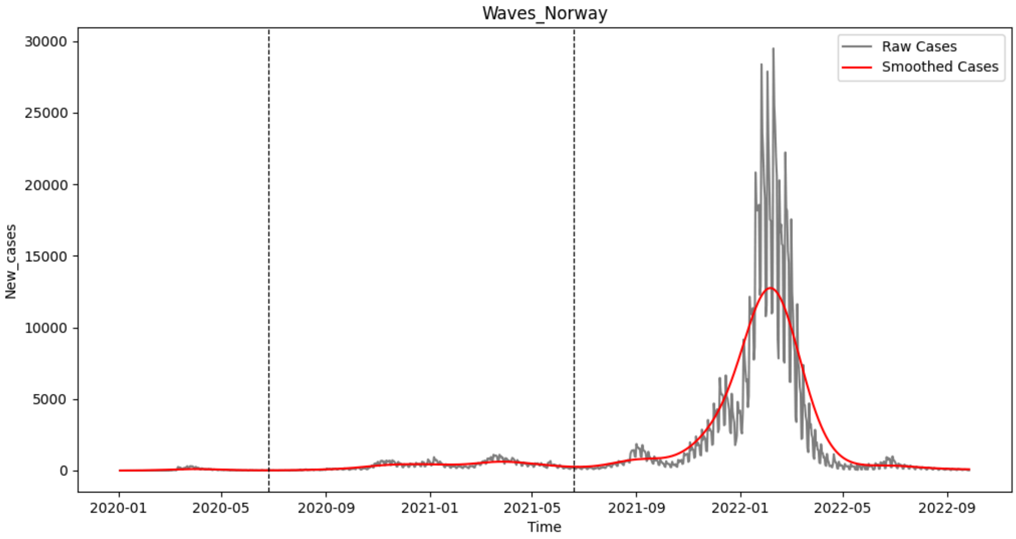

**SI Fig 8. Waves of Norway COVID-19 epidemic curve using EpidemicKabu.** The raw epidemic curve in gray shows the daily incident cases of COVID-19 between 2020 and 2022, the smoothed epidemic curve is in red, and the dashed vertical lines are the waves delimitations.

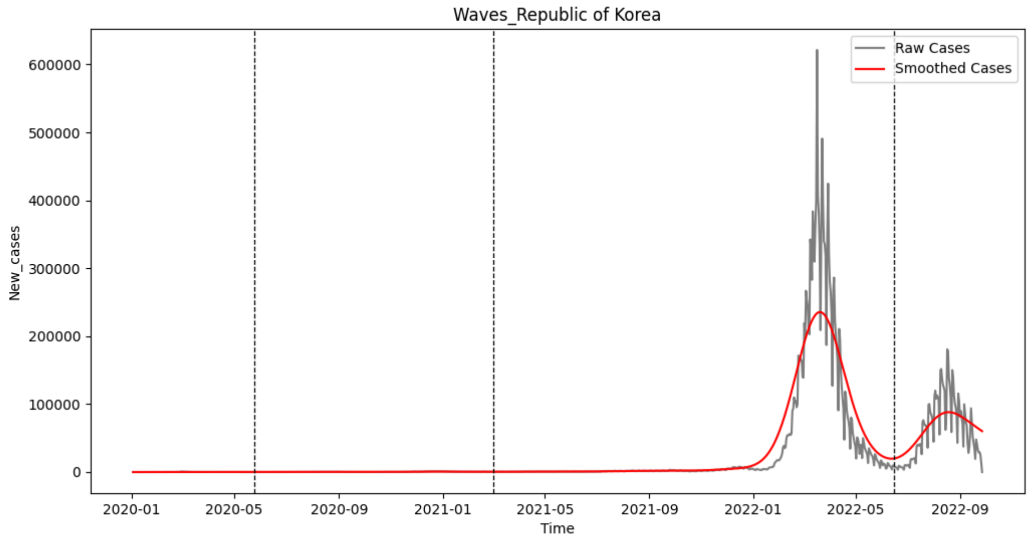

**SI Fig 9. Waves of Republic of Korea COVID-19 epidemic curve using EpidemicKabu.** The raw epidemic curve in gray shows the daily incident cases of COVID-19 between 2020 and 2022, the smoothed epidemic curve is in red, and the dashed vertical lines are the waves delimitations.

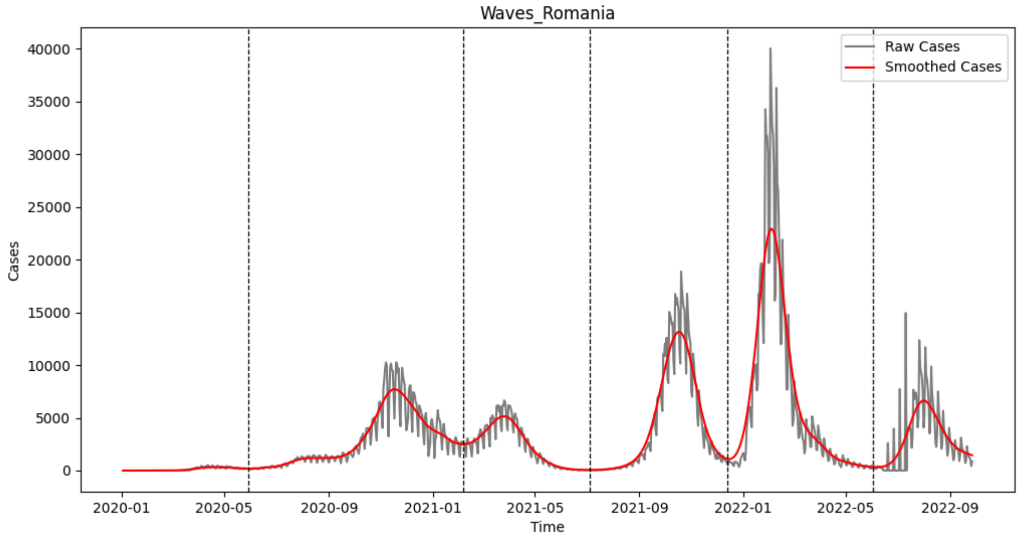

**SI Fig 10. Waves of Romania COVID-19 epidemic curve using EpidemicKabu.** The raw epidemic curve in gray shows the daily incident cases of COVID-19 between 2020 and 2022, the smoothed epidemic curve is in red, and the dashed vertical lines are the waves delimitations.

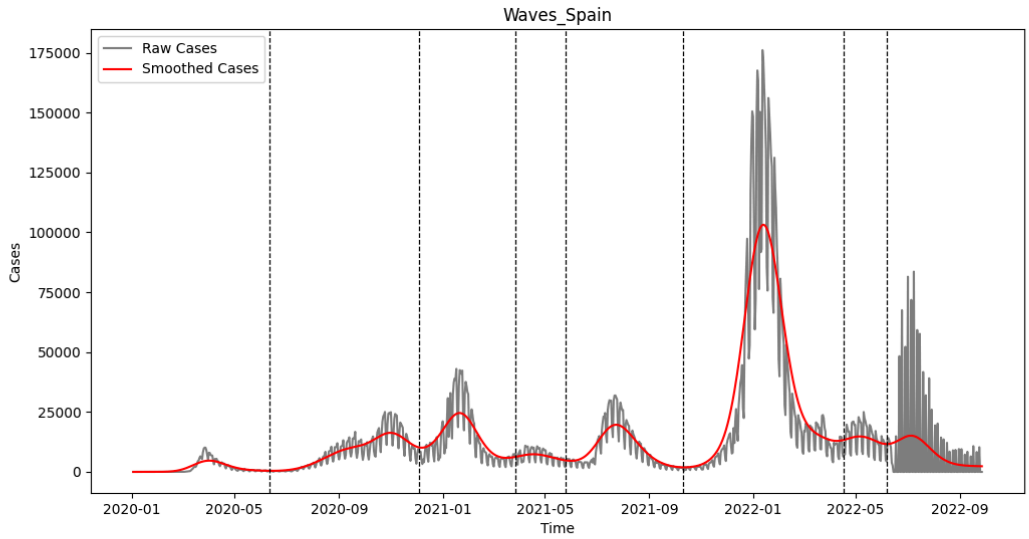

**SI Fig 11. Waves of Spain COVID-19 epidemic curve using EpidemicKabu.** The raw epidemic curve in gray shows the daily incident cases of COVID-19 between 2020 and 2022, the smoothed epidemic curve is in red, and the dashed vertical lines are the waves delimitations.

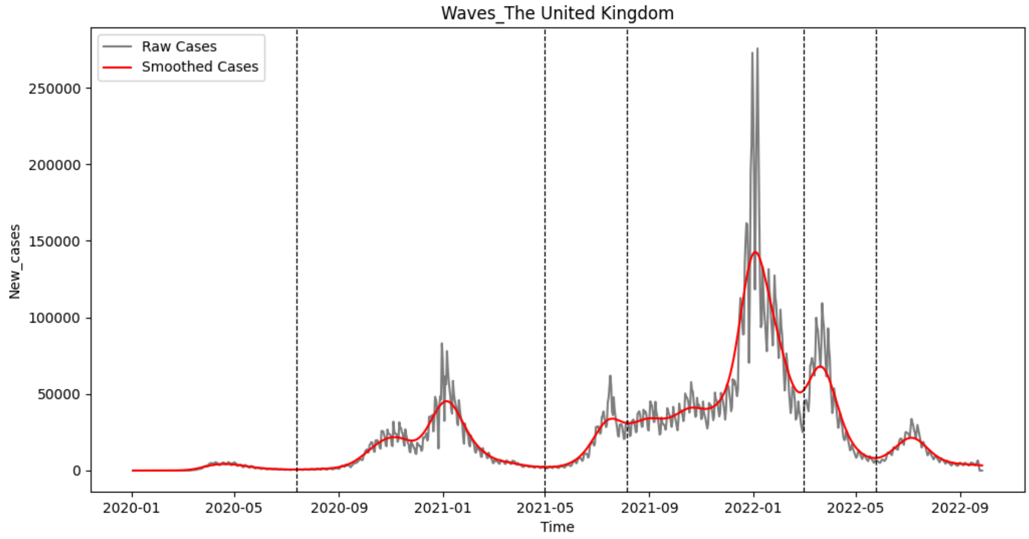

**SI Fig 12. Waves of United Kingdom COVID-19 epidemic curve using EpidemicKabu.** The raw epidemic curve in gray  shows the daily incident cases of COVID-19 between 2020 and 2022, the smoothed epidemic curve is in red, and the dashed vertical lines are the waves delimitations.

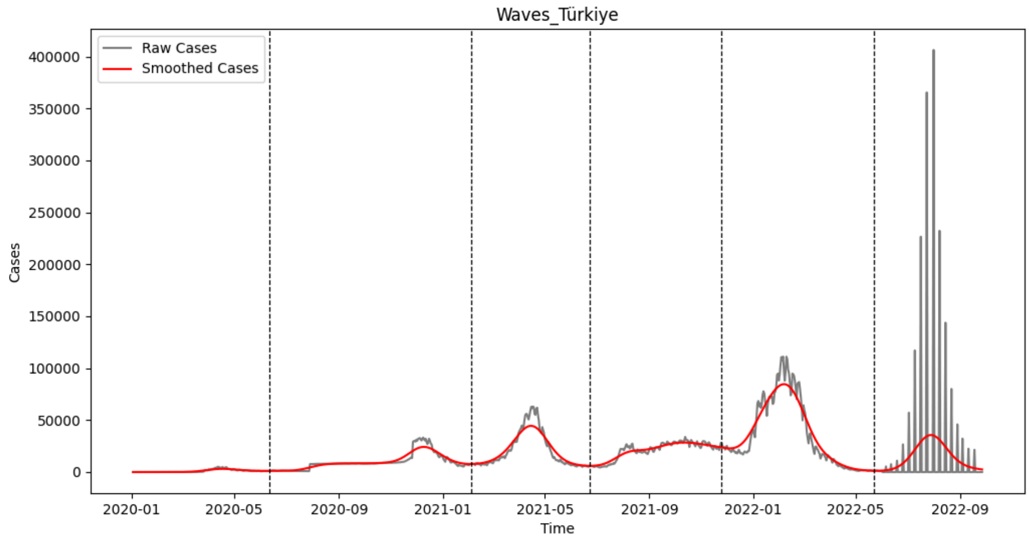

**SI Fig 13. Waves of Türkiye COVID-19 epidemic curve using EpidemicKabu.** The raw epidemic curve in gray  shows the daily incident cases of COVID-19 between 2020 and 2022, the smoothed epidemic curve is in red, and the dashed vertical lines are the waves delimitations.

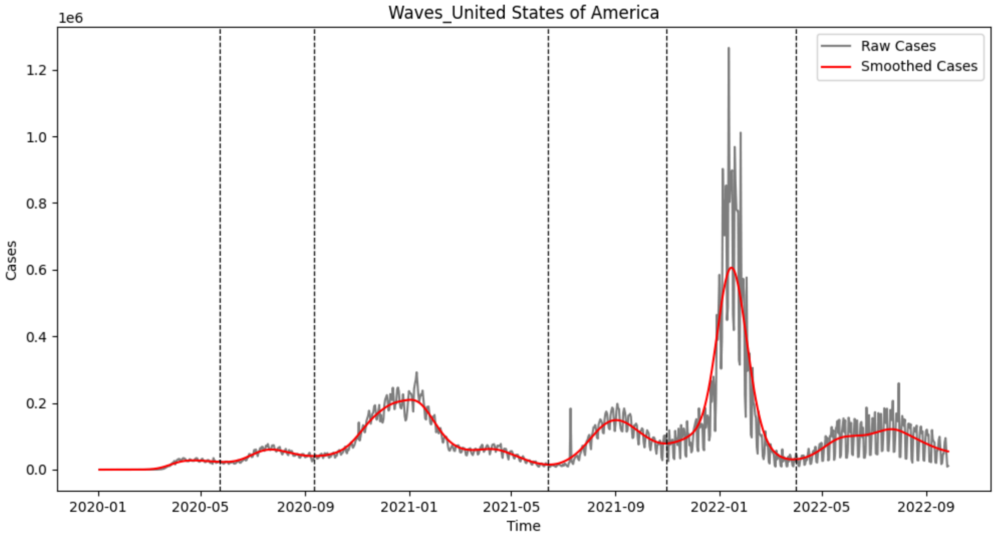

**SI Fig 14. Waves of United States COVID-19 epidemic curve using EpidemicKabu.** The raw epidemic curve in gray  shows the daily incident cases of COVID-19 between 2020 and 2022, the smoothed epidemic curve is in red, and the dashed vertical lines are the waves delimitations.

### Peaks and Valleys

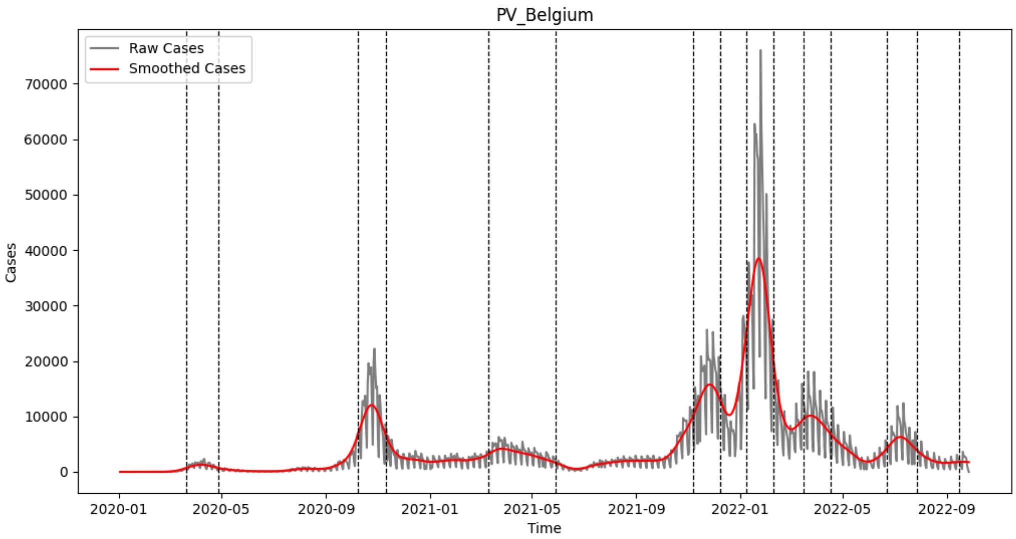

**SI Fig 15. Peaks and Valleys of Belgium COVID-19 epidemic curve using EpidemicKabu.** The raw epidemic curve in gray shows the daily incident cases of COVID-19 between 2020 and 2022, the smoothed epidemic curve is in red, and the dashed vertical lines are the peaks and valleys delimitations. *PV: Peaks and Valleys.

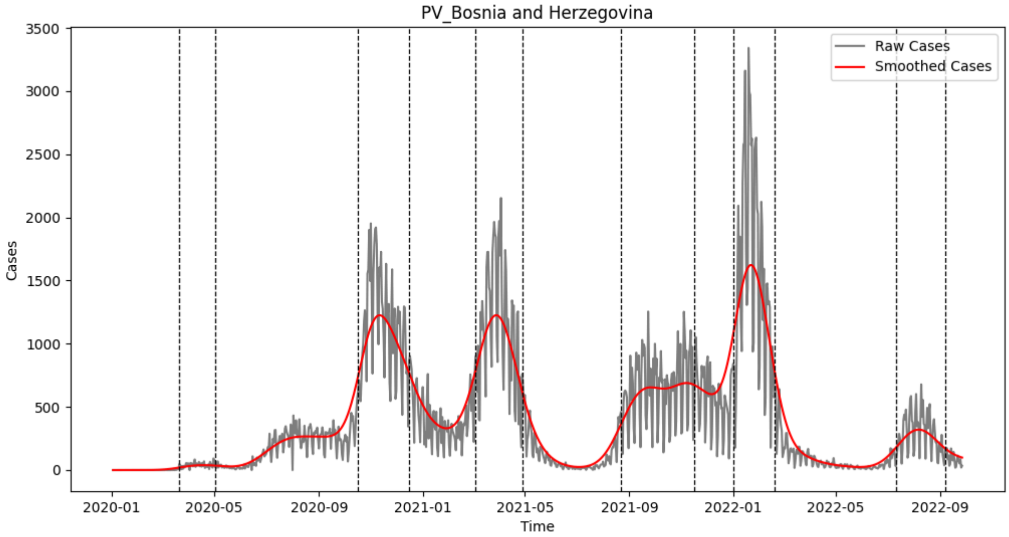

**SI Fig 16. Peaks and Valleys of Bosnia and Herzegovina COVID-19 epidemic curve using EpidemicKabu.** The raw epidemic curve in gray shows the daily incident cases of COVID-19 between 2020 and 2022, the smoothed epidemic curve is in red, and the dashed vertical lines are the peaks and valleys delimitations. *PV: Peaks and Valleys.

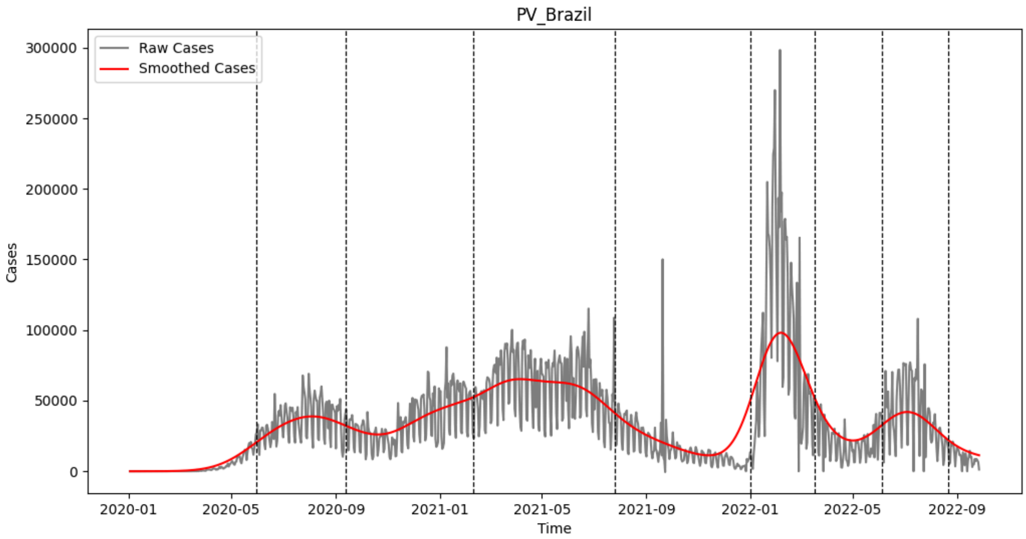

**SI Fig 17. Peaks and Valleys of Brazil COVID-19 epidemic curve using EpidemicKabu.** The raw epidemic curve in gray shows the daily incident cases of COVID-19 between 2020 and 2022, the smoothed epidemic curve is in red, and the dashed vertical lines are the peaks and valleys delimitations. *PV: Peaks and Valleys.

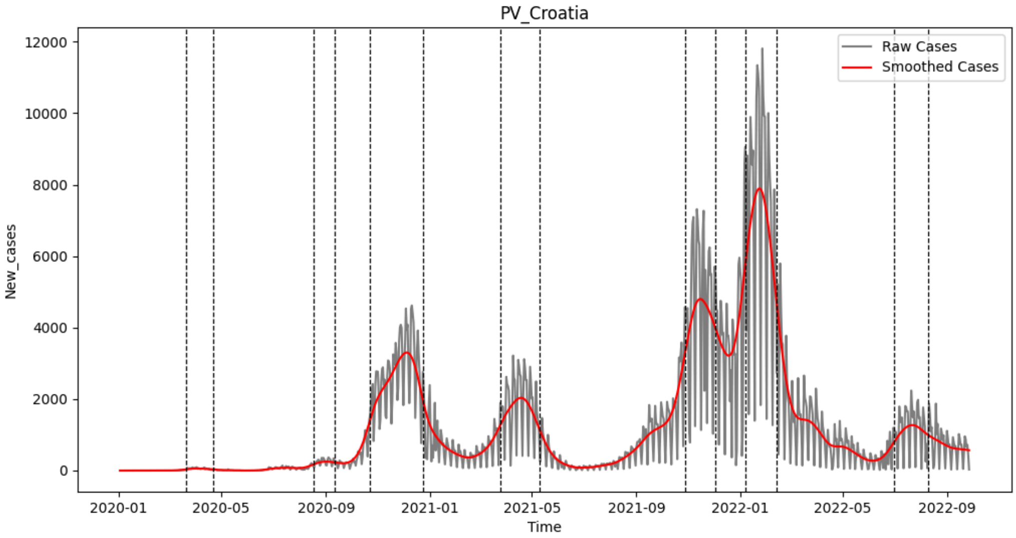

**SI Fig 18. Peaks and Valleys of Croatia COVID-19 epidemic curve using EpidemicKabu.** The raw epidemic curve in gray shows the daily incident cases of COVID-19 between 2020 and 2022, the smoothed epidemic curve is in red, and the dashed vertical lines are the peaks and valleys delimitations. *PV: Peaks and Valleys.

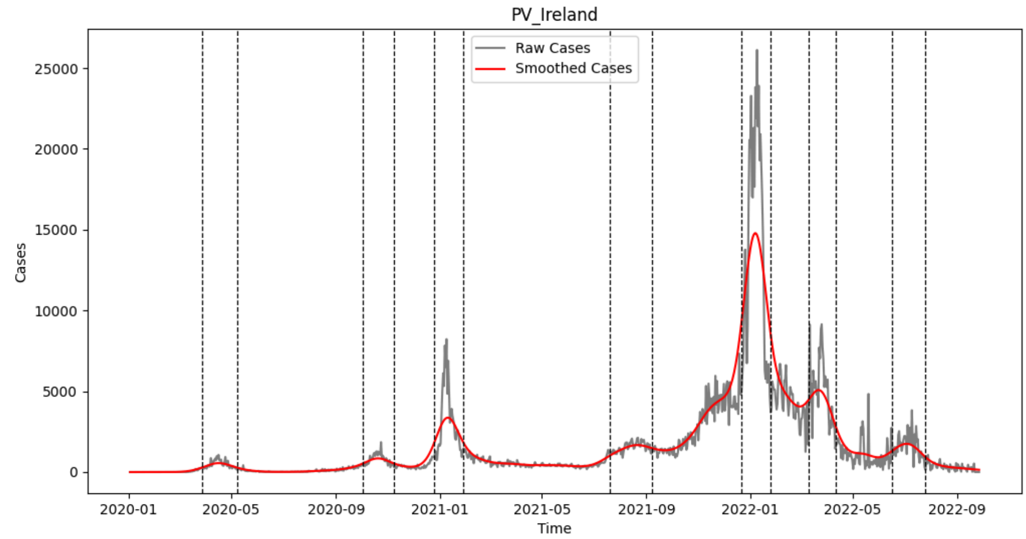

**SI Fig 19. Peaks and Valleys of Ireland COVID-19 epidemic curve using EpidemicKabu.** The raw epidemic curve in gray shows the daily incident cases of COVID-19 between 2020 and 2022, the smoothed epidemic curve is in red, and the dashed vertical lines are the peaks and valleys delimitations. *PV: Peaks and Valleys.

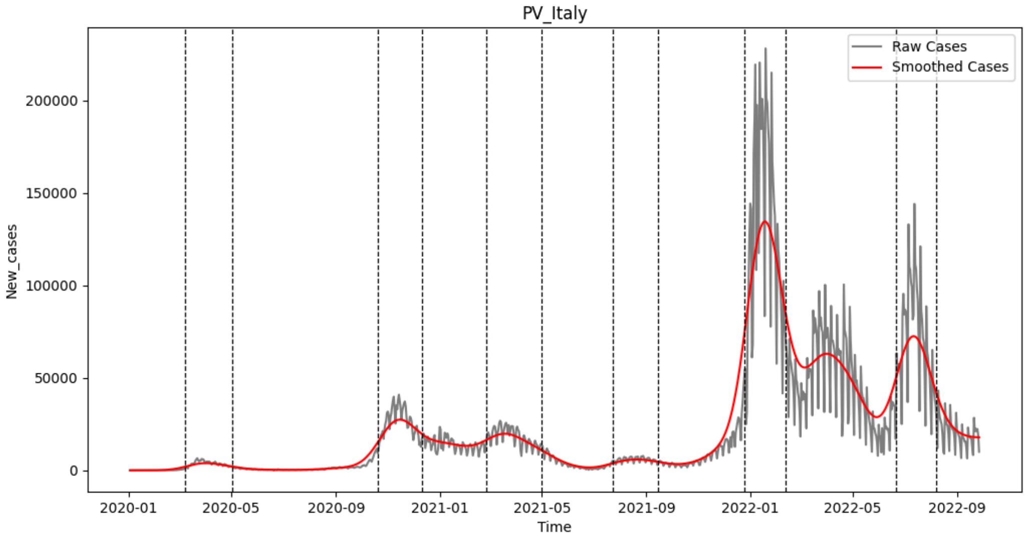

**SI Fig 20. Peaks and Valleys of Italy COVID-19 epidemic curve using EpidemicKabu.** The raw epidemic curve in gray shows the daily incident cases of COVID-19 between 2020 and 2022, the smoothed epidemic curve is in red, and the dashed vertical lines are the peaks and valleys delimitations. *PV: Peaks and Valleys.

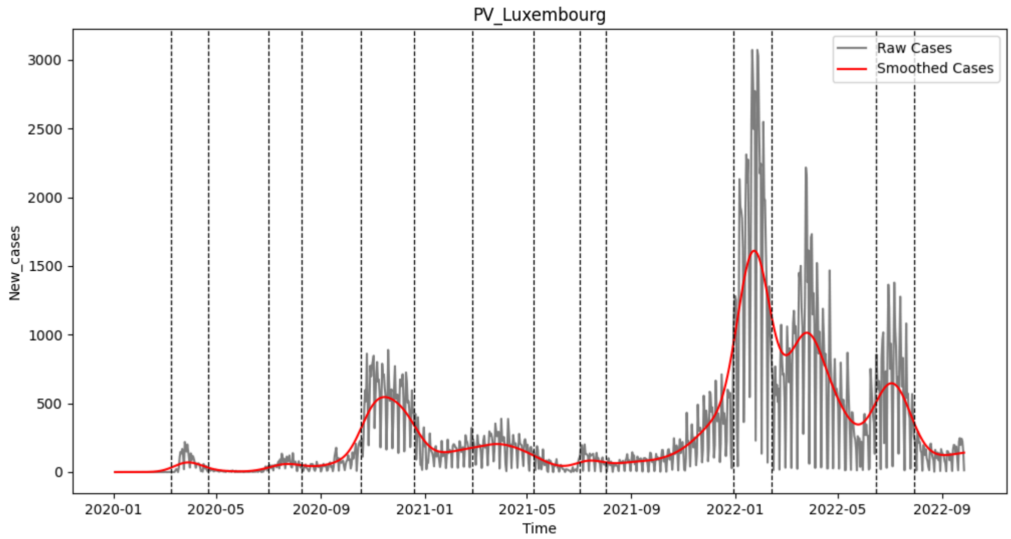

**SI Fig 21. Peaks and Valleys of Luxembourg COVID-19 epidemic curve using EpidemicKabu.** The raw epidemic curve in gray shows the daily incident cases of COVID-19 between 2020 and 2022, the smoothed epidemic curve is in red, and the dashed vertical lines are the peaks and valleys delimitations. *PV: Peaks and Valleys.

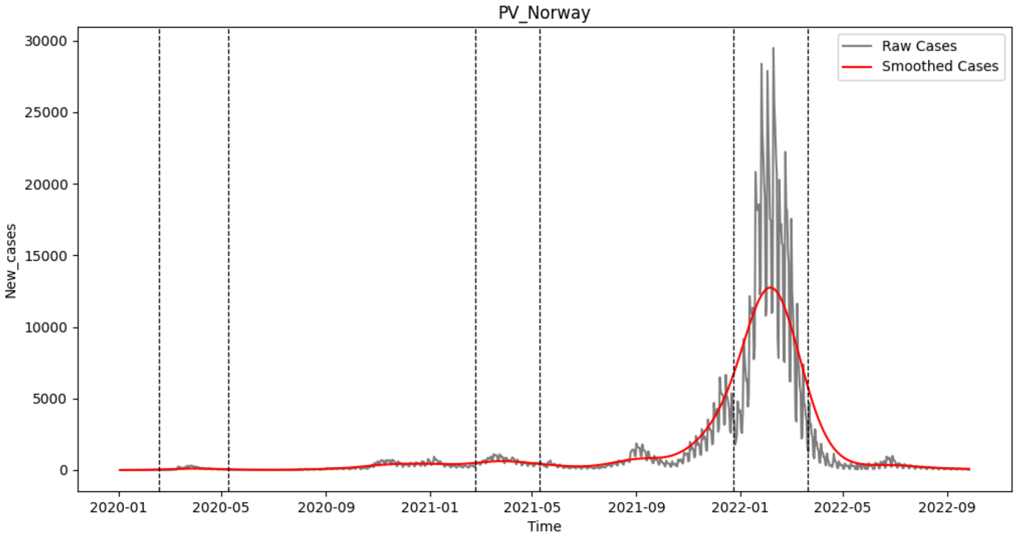

**SI Fig 22. Peaks and Valleys of Norway COVID-19 epidemic curve using EpidemicKabu.** The raw epidemic curve in gray shows the daily incident cases of COVID-19 between 2020 and 2022, the smoothed epidemic curve is in red, and the dashed vertical lines are the peaks and valleys delimitations. *PV: Peaks and Valleys.

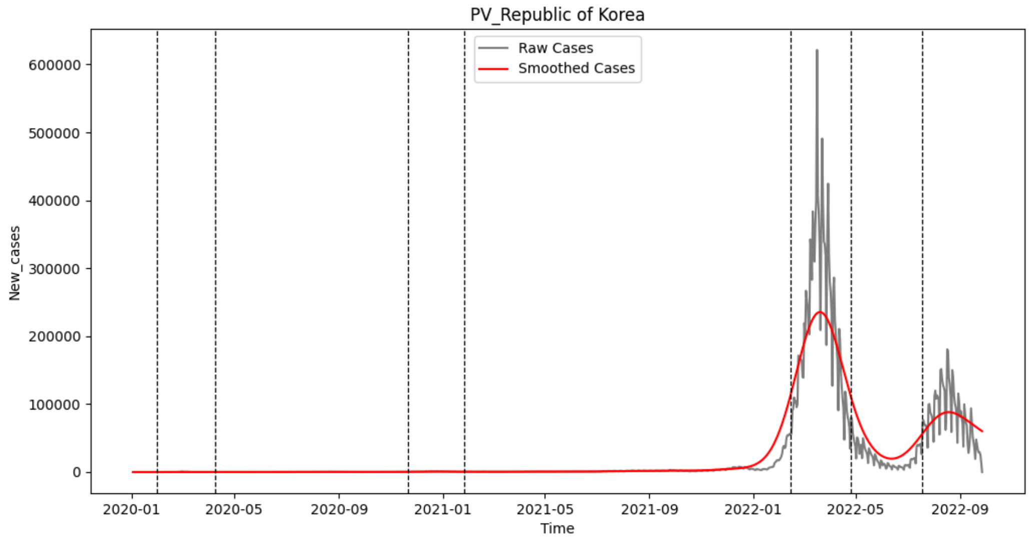

**SI Fig 23. Peaks and Valleys of Republic of Korea COVID-19 epidemic curve using EpidemicKabu.** The raw epidemic curve in gray shows the daily incident cases of COVID-19 between 2020 and 2022, the smoothed epidemic curve is in red, and the dashed vertical lines are the peaks and valleys delimitations. *PV: Peaks and Valleys.

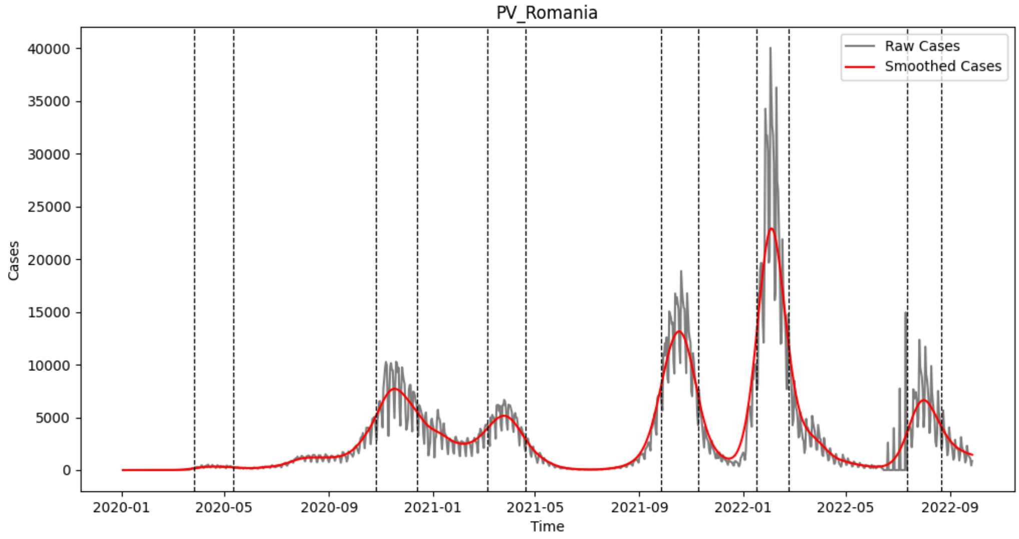

**SI Fig 24. Peaks and Valleys of Romania COVID-19 epidemic curve using EpidemicKabu.** The raw epidemic curve in gray shows the daily incident cases of COVID-19 between 2020 and 2022, the smoothed epidemic curve is in red, and the dashed vertical lines are the peaks and valleys delimitations. *PV: Peaks and Valleys.

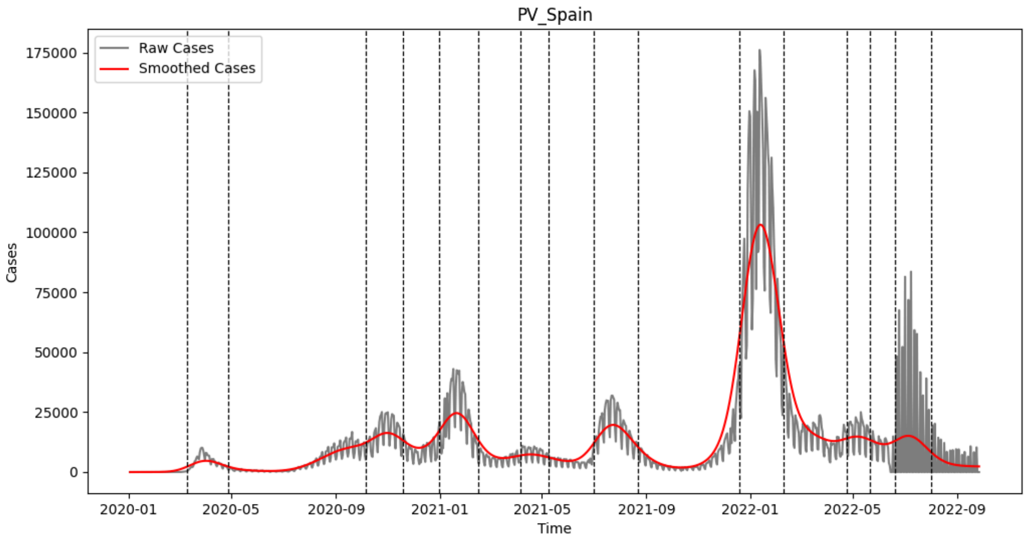

**SI Fig 25. Peaks and Valleys of Spain COVID-19 epidemic curve using EpidemicKabu.** The raw epidemic curve in gray shows the daily incident cases of COVID-19 between 2020 and 2022, the smoothed epidemic curve is in red, and the dashed vertical lines are the peaks and valleys delimitations. *PV: Peaks and Valleys.

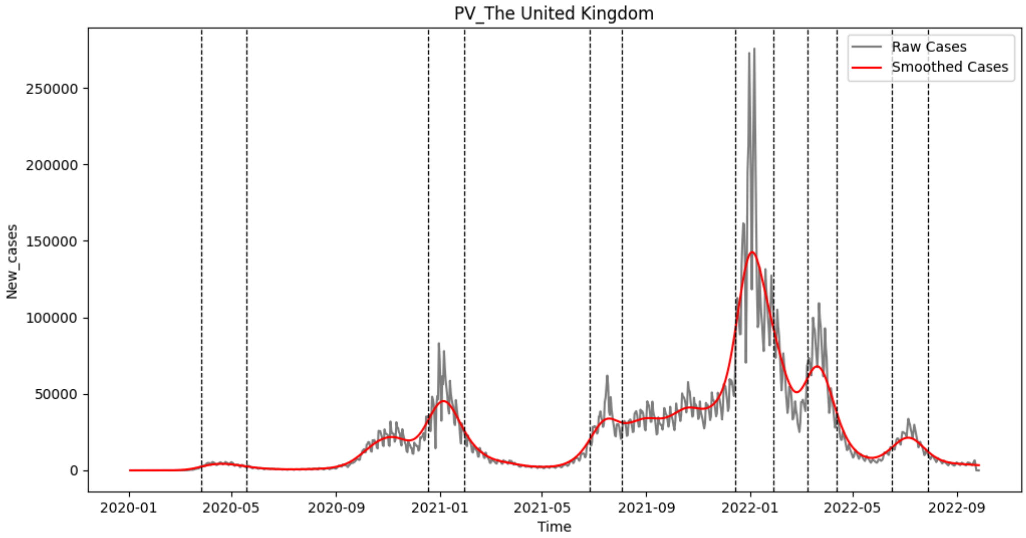

**SI Fig 26. Peaks and Valleys of United Kingdom COVID-19 epidemic curve using EpidemicKabu.** The raw epidemic curve in gray shows the daily incident cases of COVID-19 between 2020 and 2022, the smoothed epidemic curve is in red, and the dashed vertical lines are the peaks and valleys delimitations. *PV: Peaks and Valleys.

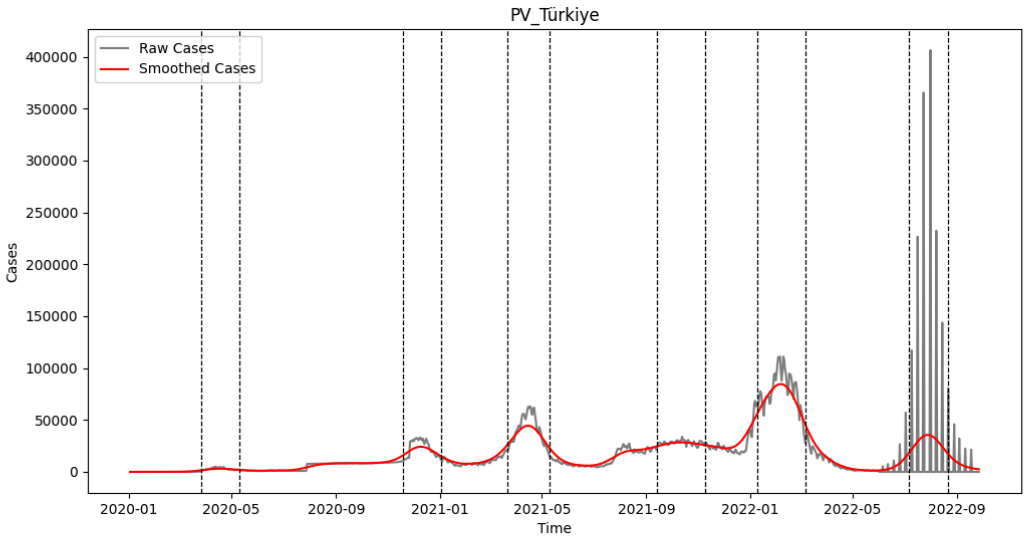

**SI Fig 27. Peaks and Valleys of Türkiye COVID-19 epidemic curve using EpidemicKabu.** The raw epidemic curve in gray shows the daily incident cases of COVID-19 between 2020 and 2022, the smoothed epidemic curve is in red, and the dashed vertical lines are the peaks and valleys delimitations. *PV: Peaks and Valleys.

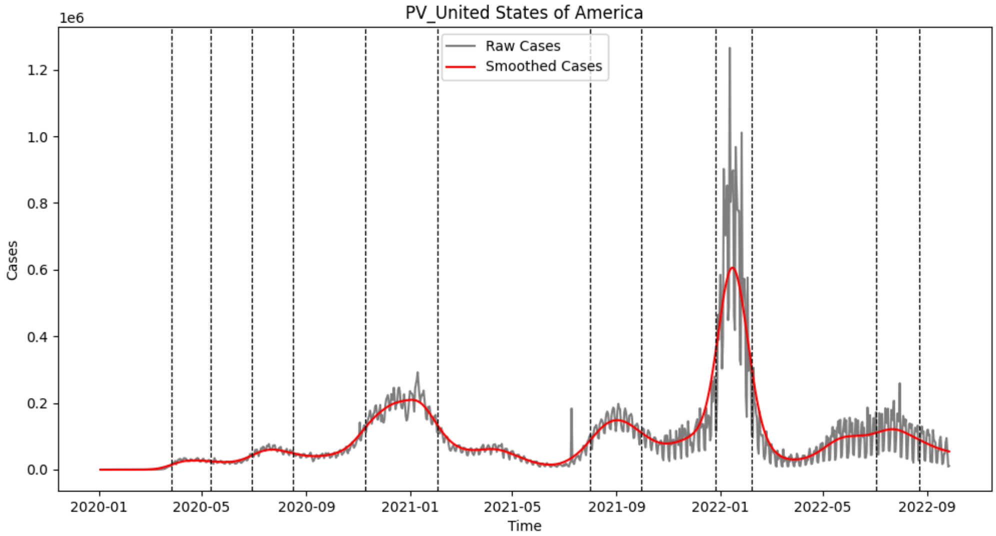

**SI Fig 28. Peaks and Valleys of United States COVID-19 epidemic curve using EpidemicKabu.** The raw epidemic curve in gray shows the daily incident cases of COVID-19 between 2020 and 2022, the smoothed epidemic curve is in red, and the dashed vertical lines are the peaks and valleys delimitations. *PV: Peaks and Valleys.

### Configuration File

**S1 Table 1. Configuration table.** It shows for each country the kernel1 to smooth epidemic curve and kernel2 to smooth its first derivative. λ refers if the used kernel was the recommended or the strong or weak constraint. A complete file with kernels for other countries is in https://github.com/LinaMRuizG/EpidemicKabuLibrary/blob/main/examples/data/configurationFile.csv

| Country | kernel1 | kernel2 | λ |
|---|---|---|---|
| Belgium | 20.8 | 20.8 | recommend |
| Bosnia and Herzegovina | 31 | 31 | recommend |
| Brazil | 47.4 | 47.4 | strong |
| Colombia | 43.6 | 43.6 | recommend |
| Croatia | 16.1 | 16.1 | weak |
| Ireland | 23.8 | 23.8 | recommend |
| Italy | 30.9 | 30.9 | recommend |
| Luxembourg | 29.7 | 29.7 | recommend |
| Norway | 52.8 | 52.8 | strong |
| Republic of Korea | 43.4 | 43.4 | strong |
| Romania | 25.2 | 25.2 | recommend |
| Spain | 27.6 | 27.6 | recommend |
| The United Kingdom | 24 | 24 | recommend |
| United States | 20.3 | 20.3 | recommend |
| Turkiye | 28 | 28 | strong |

### Waves of the epidemic curve base on the indicator

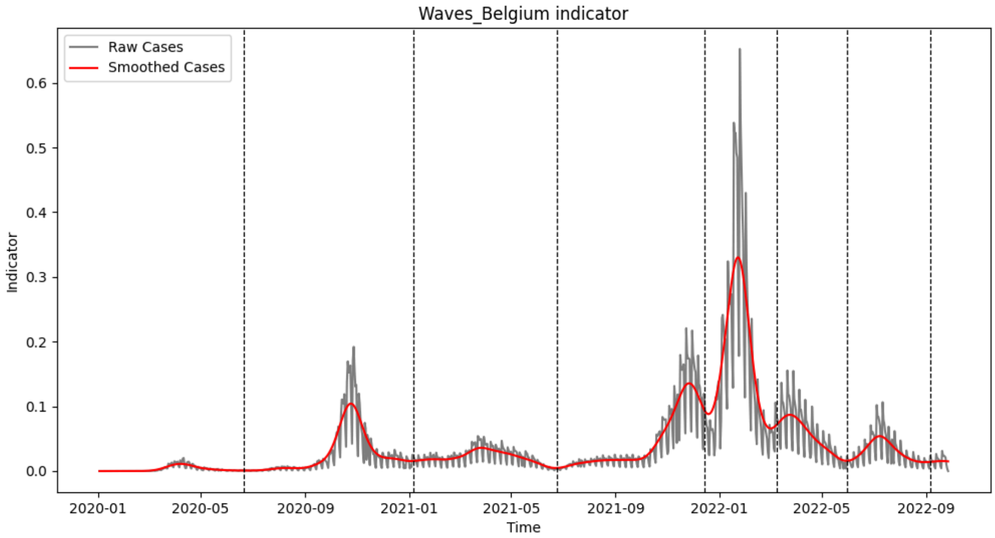

**SI Fig 29. Waves of Belgium COVID-19 epidemic curve with an indicator.** The raw epidemic curve in gray shows the daily indicator, the smoothed epidemic curve is in red, and the dashed vertical lines are the waves delimitations.

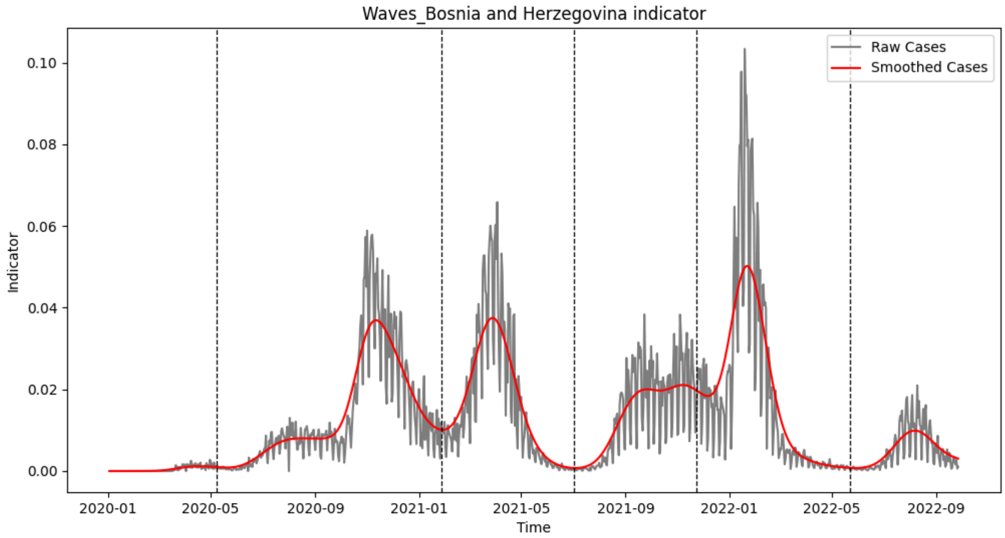

**SI Fig 30. Waves of Bosnia and Herzegovina COVID-19 epidemic curve with an indicator.** The raw epidemic curve in gray shows the daily indicator, the smoothed epidemic curve is in red, and the dashed vertical lines are the waves delimitations.

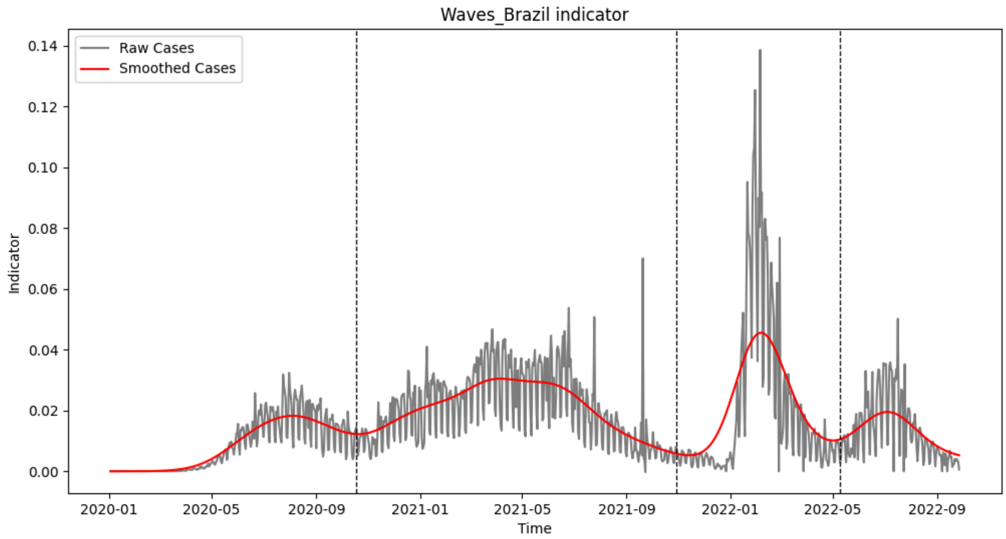

**SI Fig 31. Waves of Brazil COVID-19 epidemic curve with an indicator.** The raw epidemic curve in gray shows the daily indicator, the smoothed epidemic curve is in red, and the dashed vertical lines are the waves delimitations.

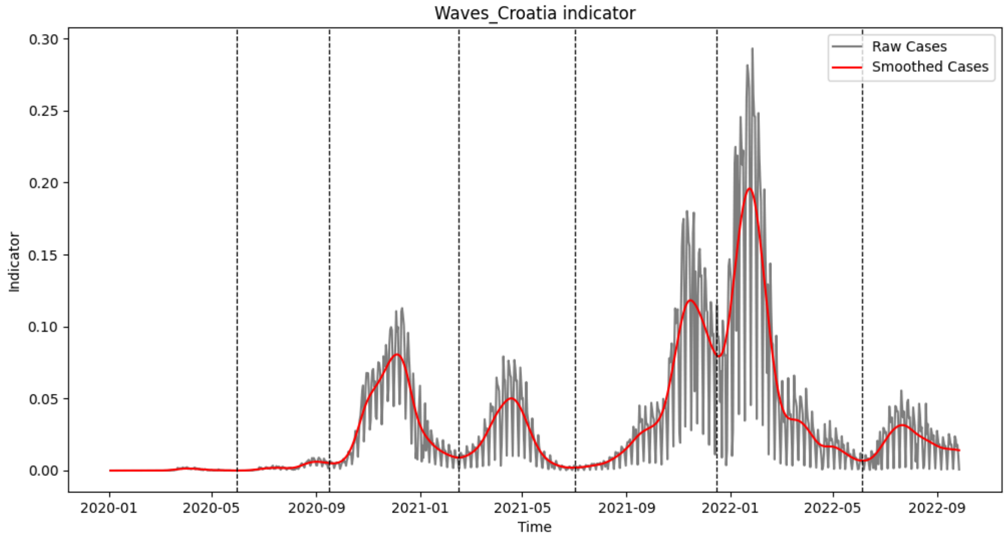

**SI Fig 32. Waves of Croatia COVID-19 epidemic curve with an indicator.** The raw epidemic curve in gray shows the daily indicator, the smoothed epidemic curve is in red, and the dashed vertical lines are the waves delimitations.

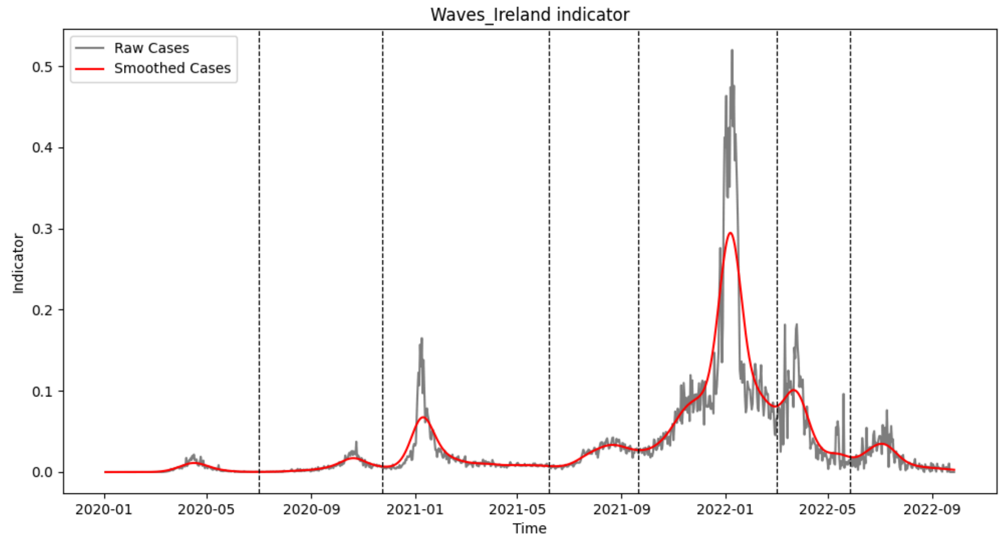

**SI Fig 33. Waves of Ireland COVID-19 epidemic curve with an indicator.** The raw epidemic curve in gray shows the daily indicator, the smoothed epidemic curve is in red, and the dashed vertical lines are the waves delimitations.

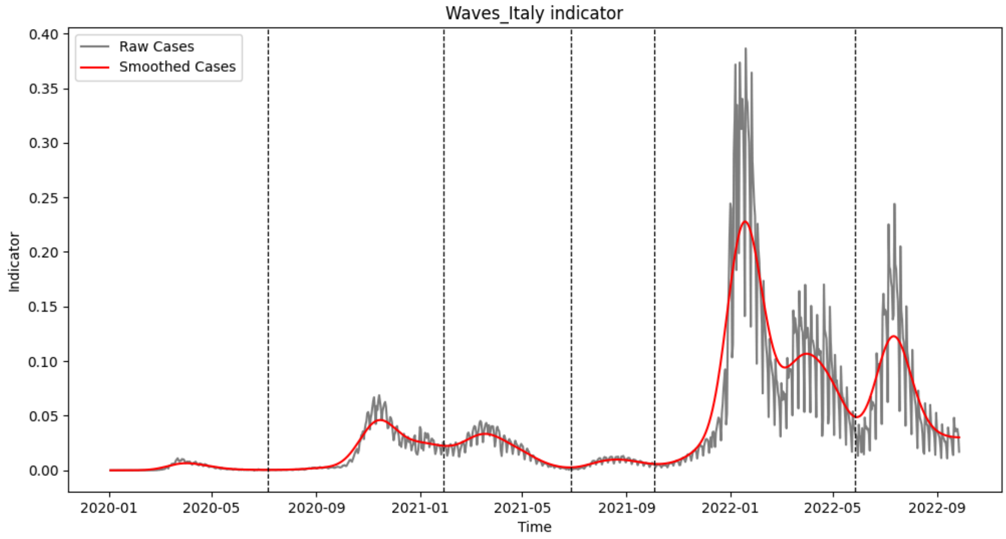

**SI Fig 34. Waves of Italy COVID-19 epidemic curve with an indicator.** The raw epidemic curve in gray shows the daily indicator, the smoothed epidemic curve is in red, and the dashed vertical lines are the waves delimitations.

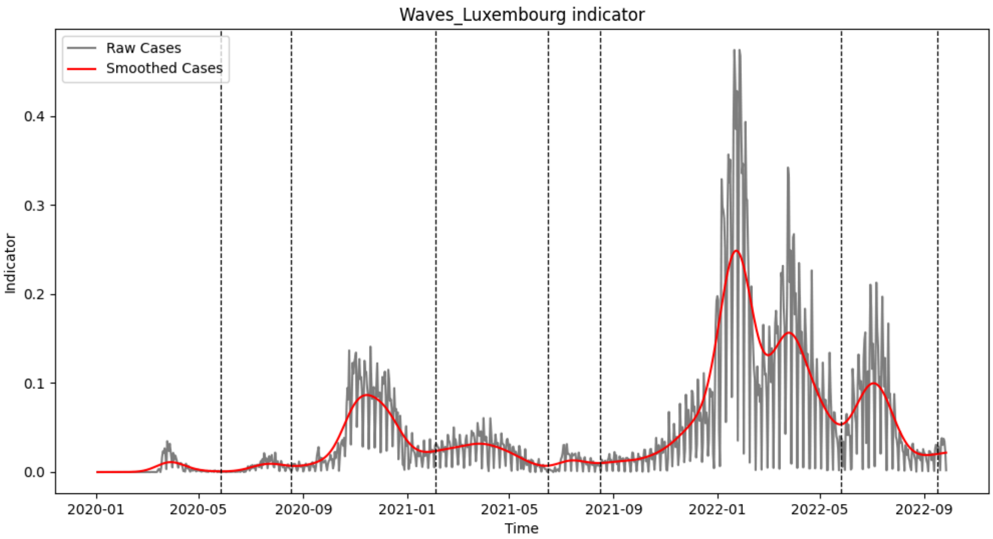

**SI Fig 35. Waves of Luxembourg COVID-19 epidemic curve with an indicator.** The raw epidemic curve in gray shows the daily indicator, the smoothed epidemic curve is in red, and the dashed vertical lines are the waves delimitations.

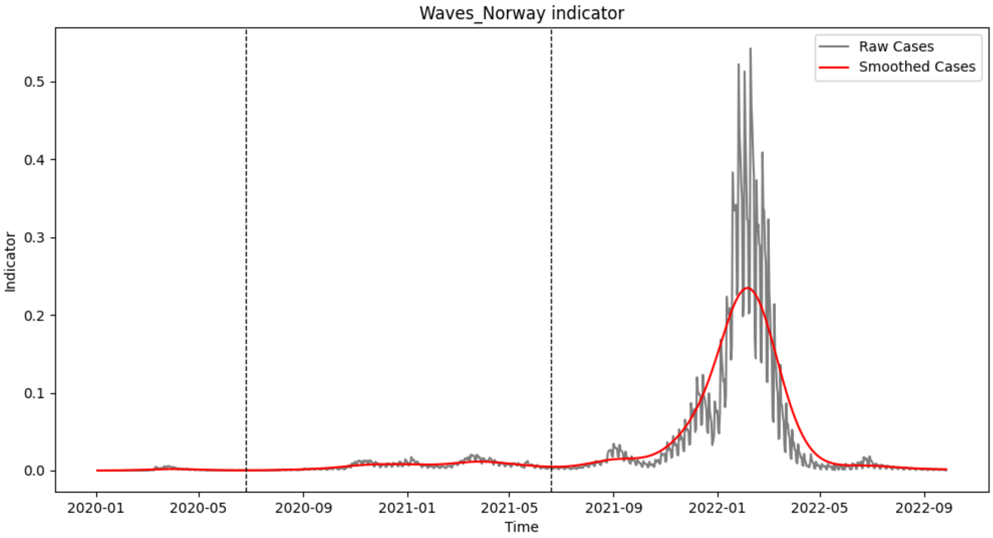

**SI Fig 36. Waves of Norway COVID-19 epidemic curve with an indicator.** The raw epidemic curve in gray shows the daily indicator, the smoothed epidemic curve is in red, and the dashed vertical lines are the waves delimitations.

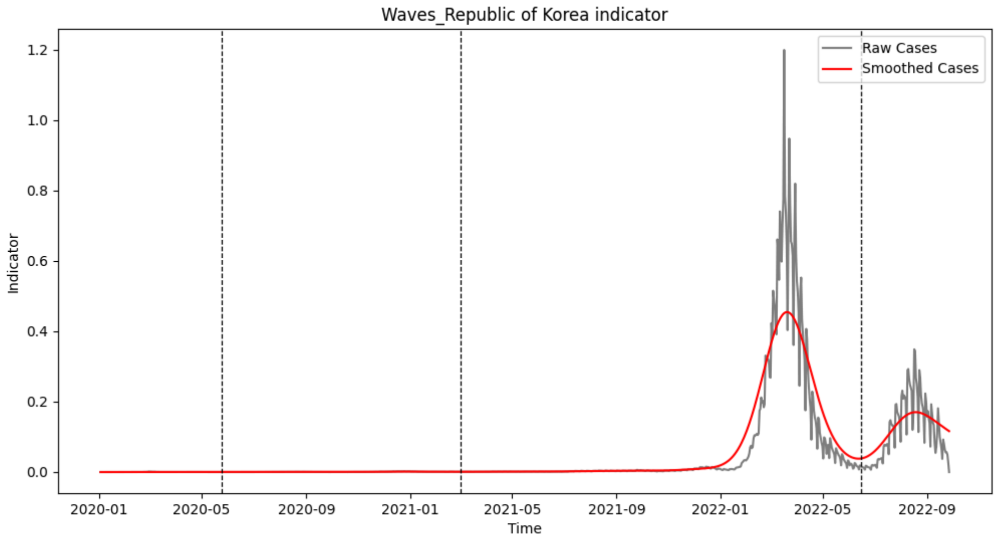

**SI Fig 37. Waves of Republic of Korea COVID-19 epidemic curve with an indicator.** The raw epidemic curve in gray shows the daily indicator, the smoothed epidemic curve is in red, and the dashed vertical lines are the waves delimitations.

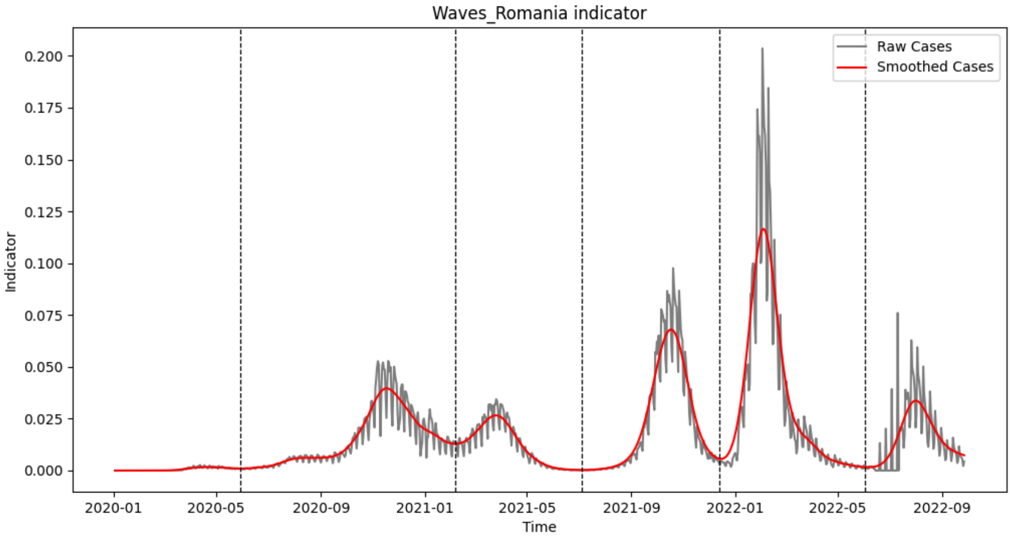

**SI Fig 38. Waves of Romania COVID-19 epidemic curve with an indicator.** The raw epidemic curve in gray shows the daily indicator, the smoothed epidemic curve is in red, and the dashed vertical lines are the waves delimitations.

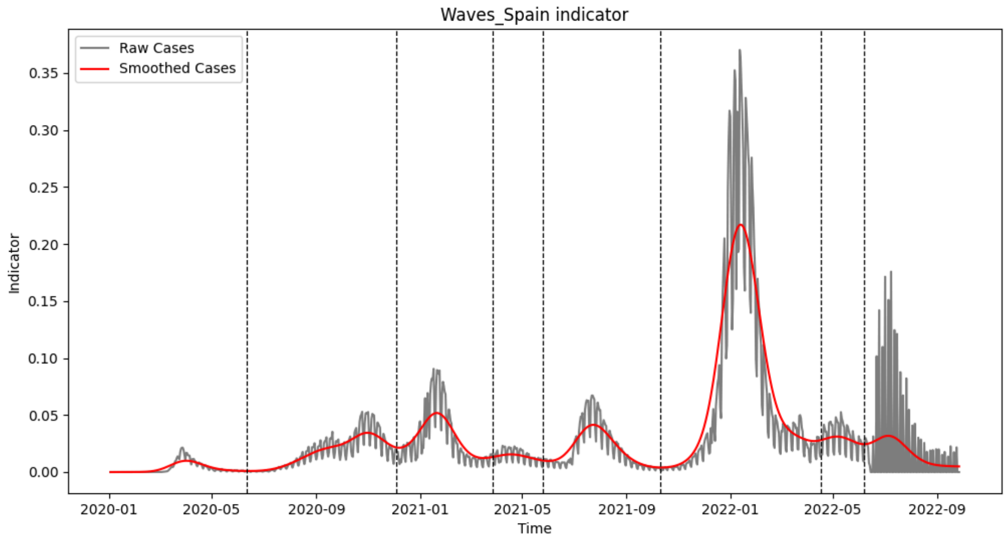

**SI Fig 39. Waves of Spain COVID-19 epidemic curve with an indicator.** The raw epidemic curve in gray shows the daily indicator, the smoothed epidemic curve is in red, and the dashed vertical lines are the waves delimitations.

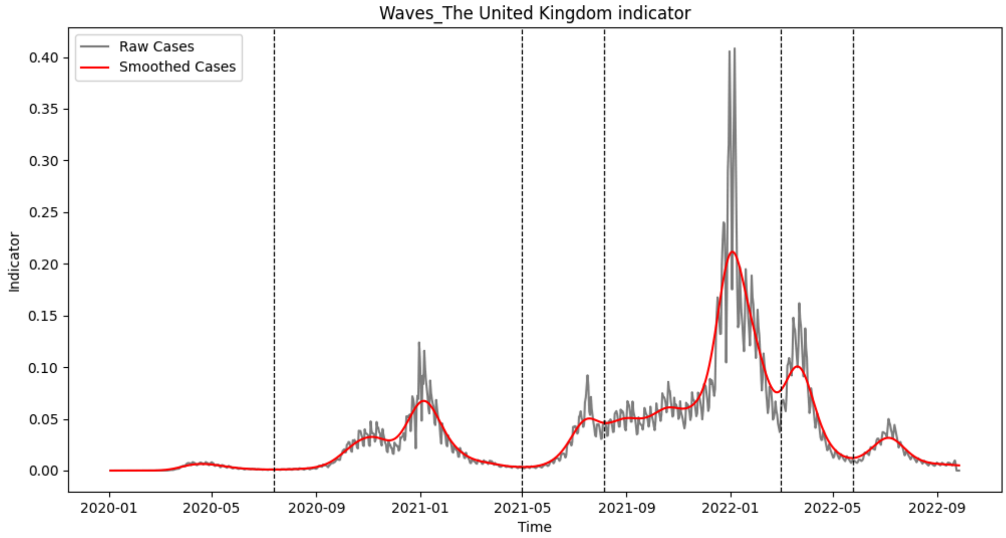

**SI Fig 40. Waves of United Kingdom COVID-19 epidemic curve with an indicator.** The raw epidemic curve in gray shows the daily indicator, the smoothed epidemic curve is in red, and the dashed vertical lines are the waves delimitations.

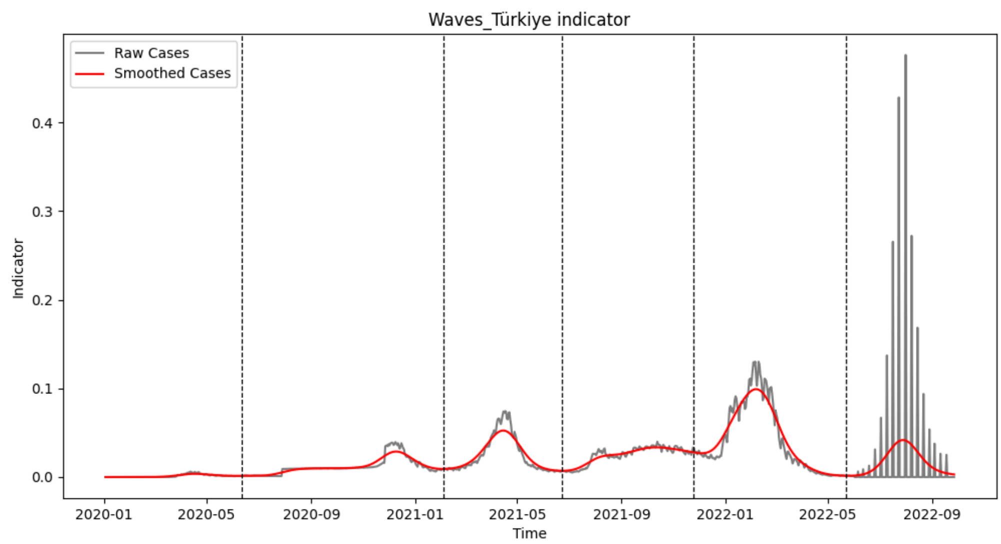

**SI Fig 41. Waves of Colombia COVID-19 epidemic curve with an indicator.** The raw epidemic curve in gray shows the daily indicator, the smoothed epidemic curve is in red, and the dashed vertical lines are the waves delimitations.

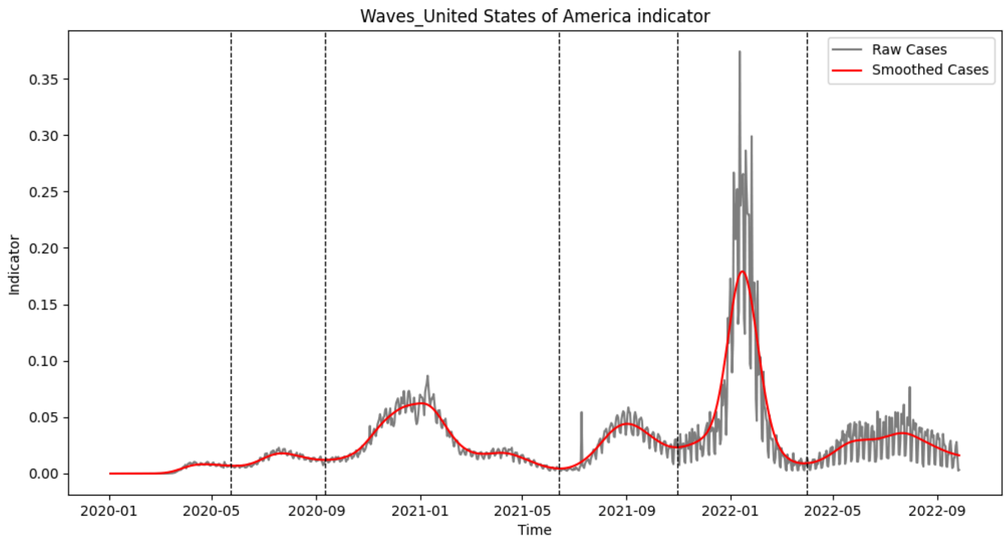

**SI Fig 42. Waves of United States COVID-19 epidemic curve with an indicator.** The raw epidemic curve in gray shows the daily indicator, the smoothed epidemic curve is in red, and the dashed vertical lines are the waves delimitations.

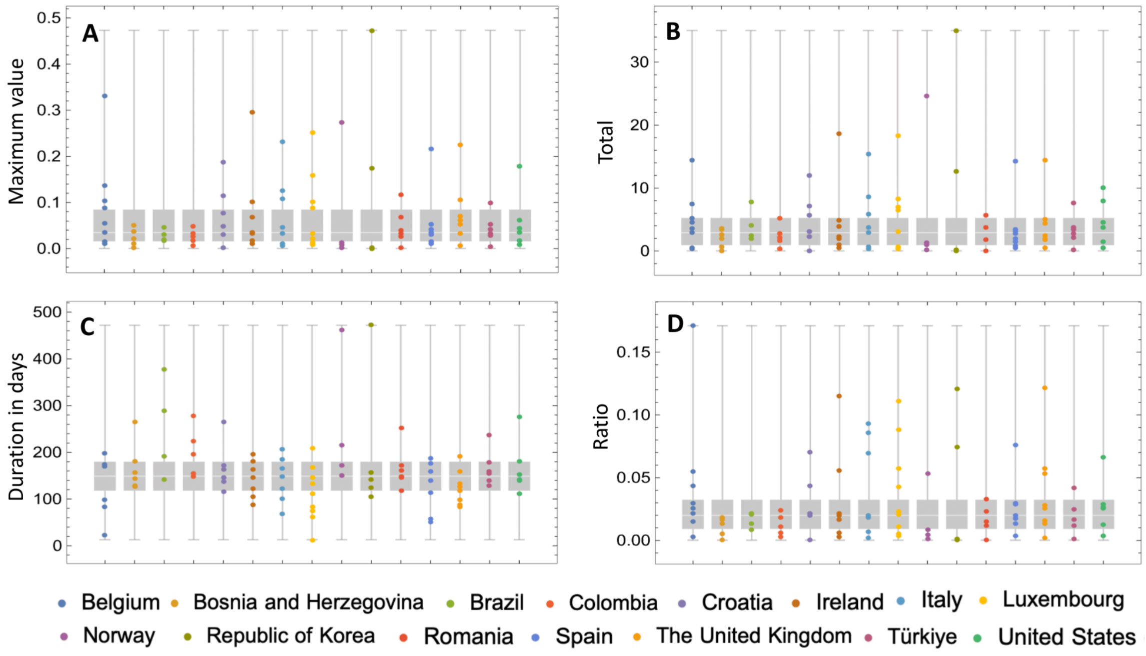

**SI Fig 43. Comparison of the measures for the COVID-19 waves of each country with all the countries.** In gray is shown the distribution of each measure for all the countries and in colors is shown the distribution for each country inside the global distribution, **(A).** Boxplots of the maximum value of the indicator for each wave of each country, **(B).** Boxplots of the sum of the indicator since the start and until the end of the wave for each wave of each country, **(C).** Boxplots of the number of days since the start and until the end of the wave for each wave of each country, **(D).** Boxplots of the ratio between the Total and the Duration in days for each wave of each country. *The bottom shows the countries names from left to right and top to bottom order as they appear in the plots.
