## Supplementary figures and images for "EpidemicKabu a new method to identify epidemic waves and their peaks and valleys"

### SIFig1.png

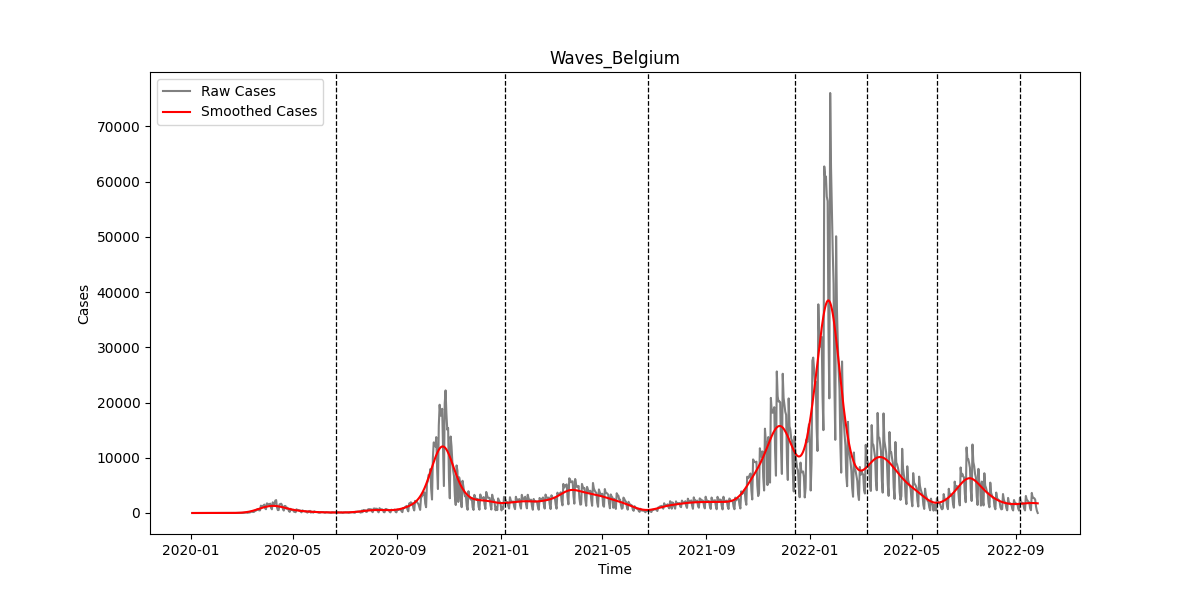

### SIFig2.png

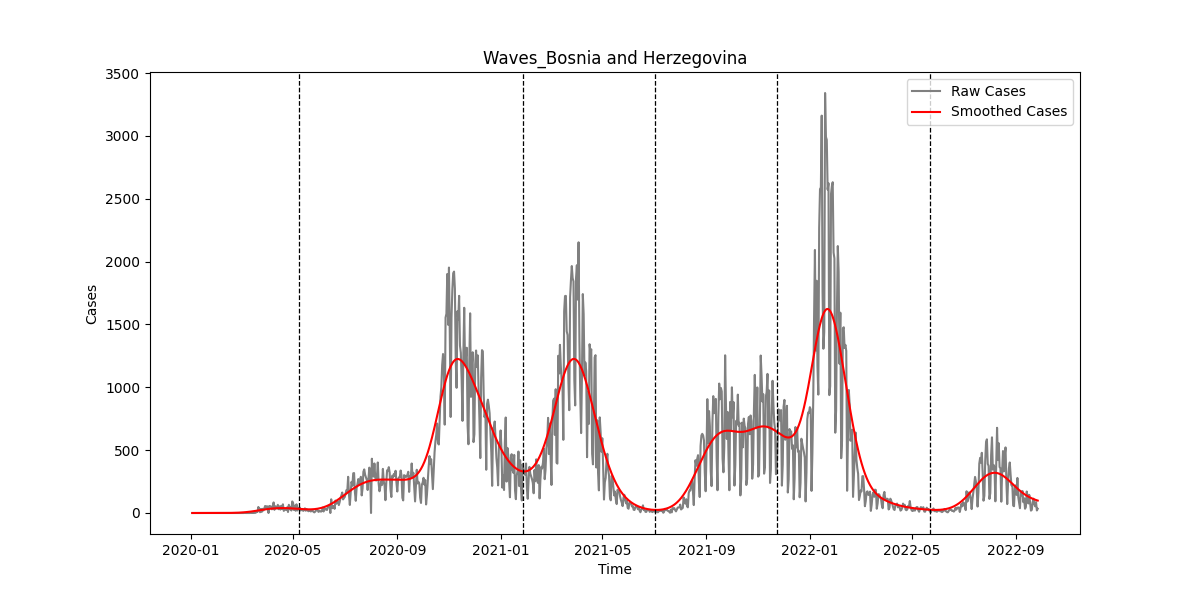

### SIFig3.png

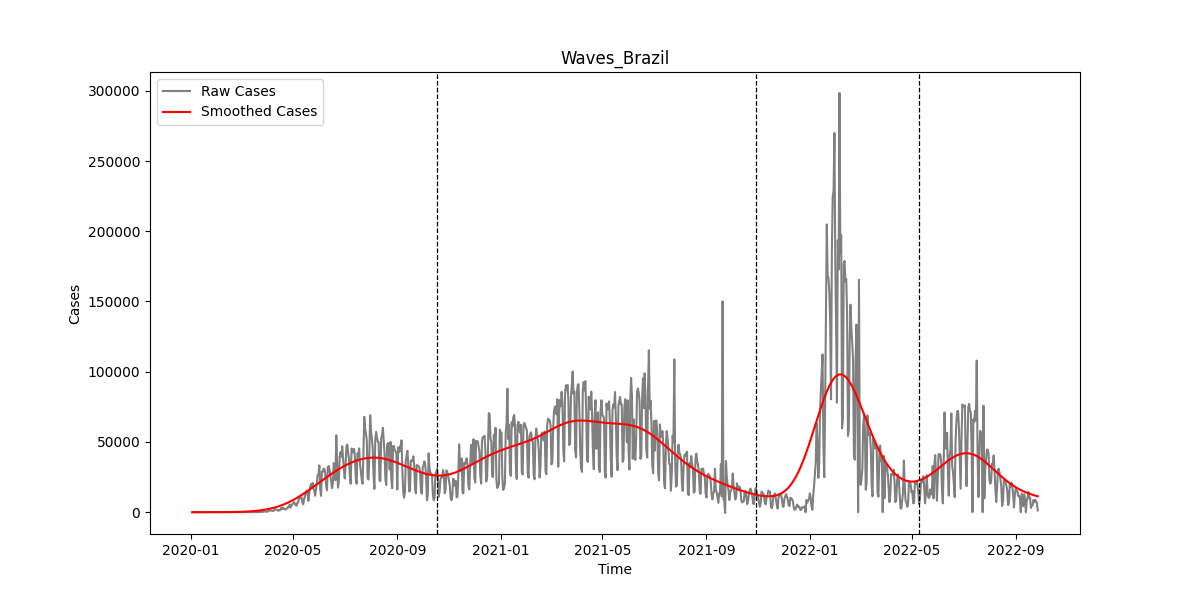

### SIFig4.png

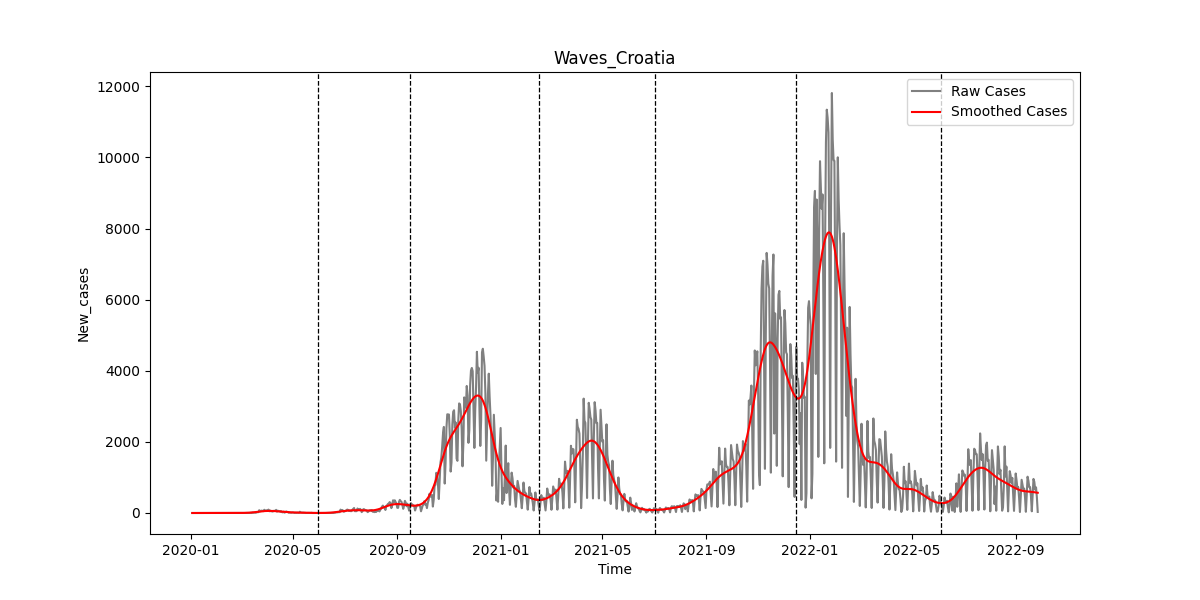

### SIFig5.png

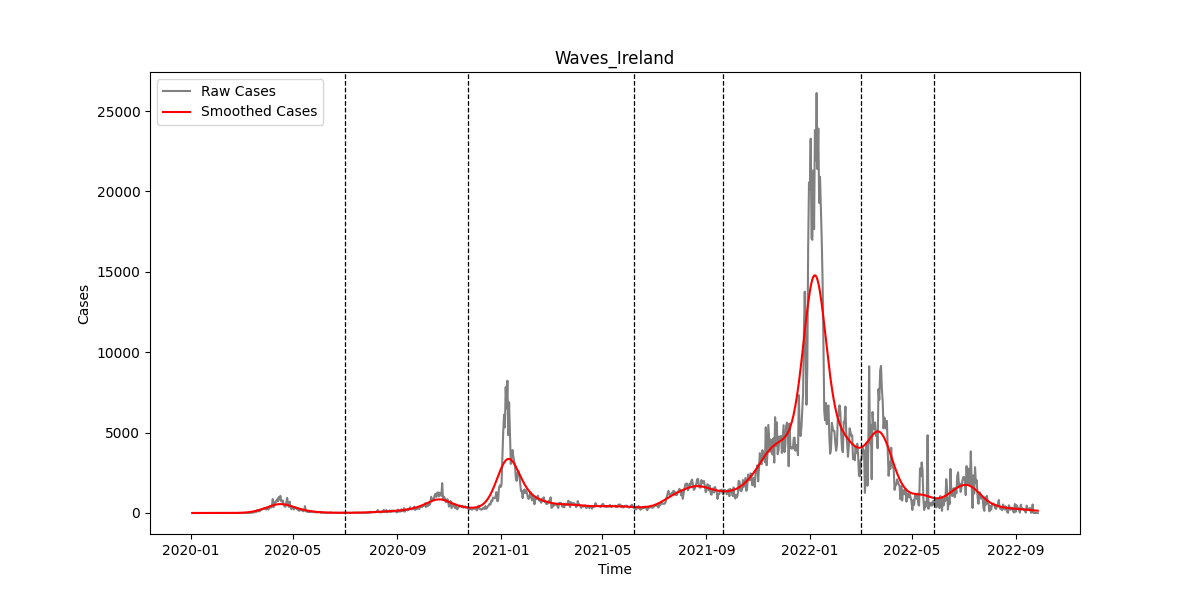

### SIFig6.png

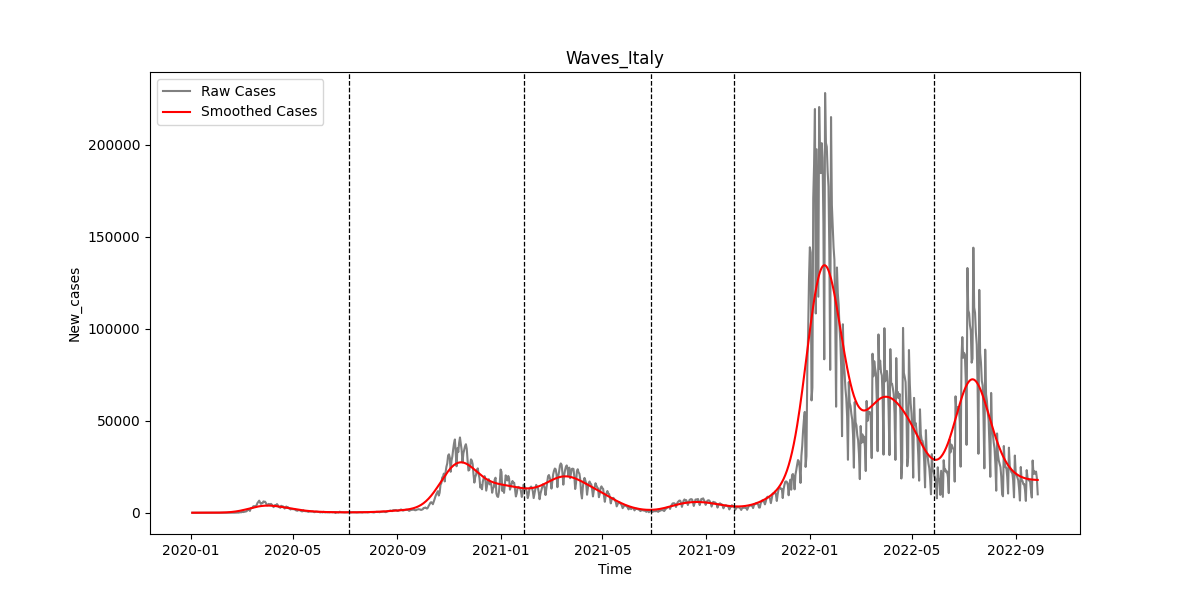

### SIFig7.png

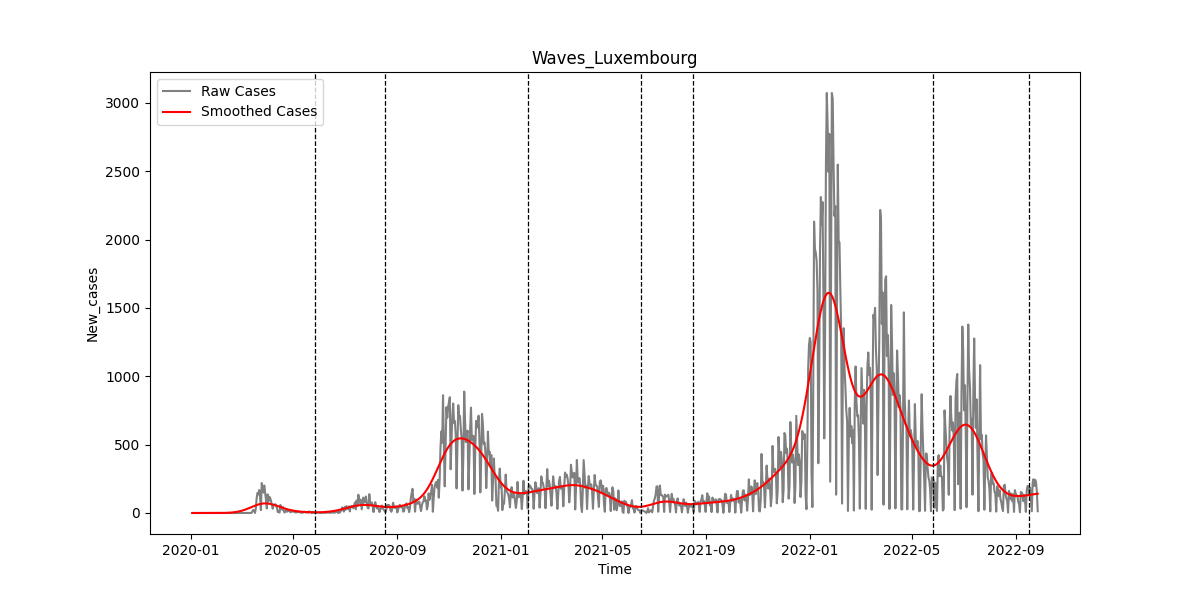

### SIFig8.png

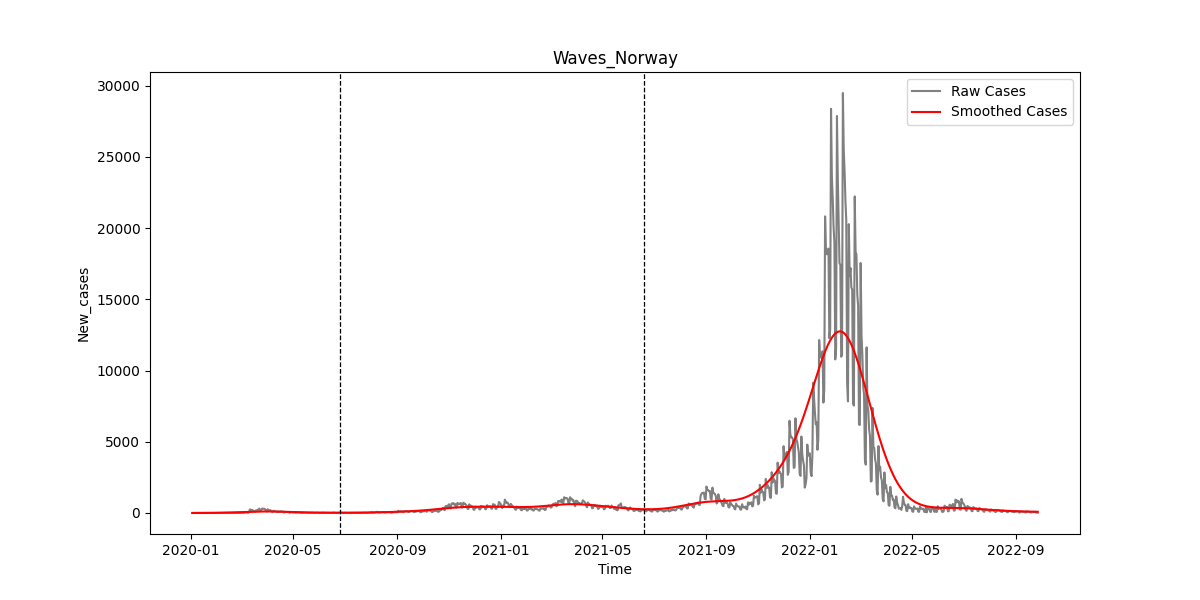

### SIFig9.png

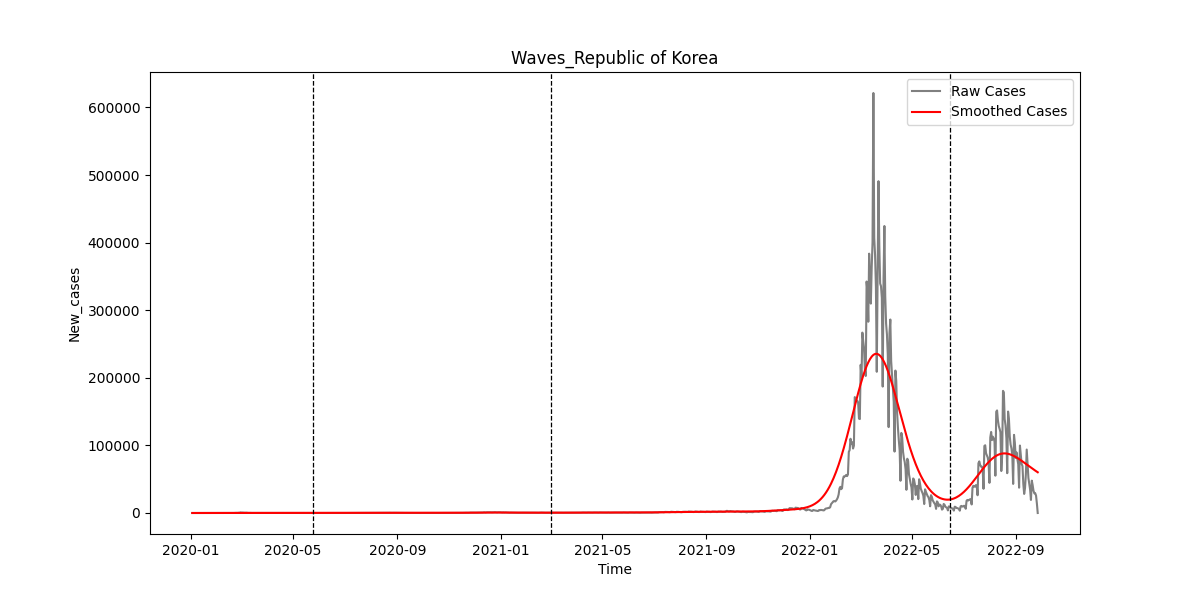

### SIFig10.png

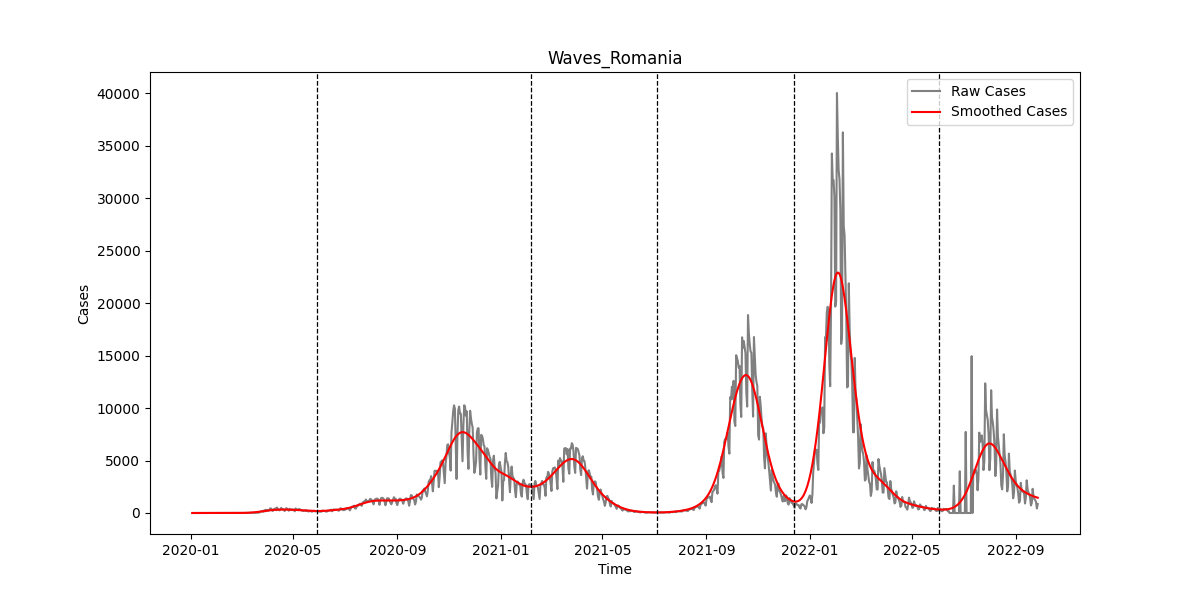

### SIFig11.png

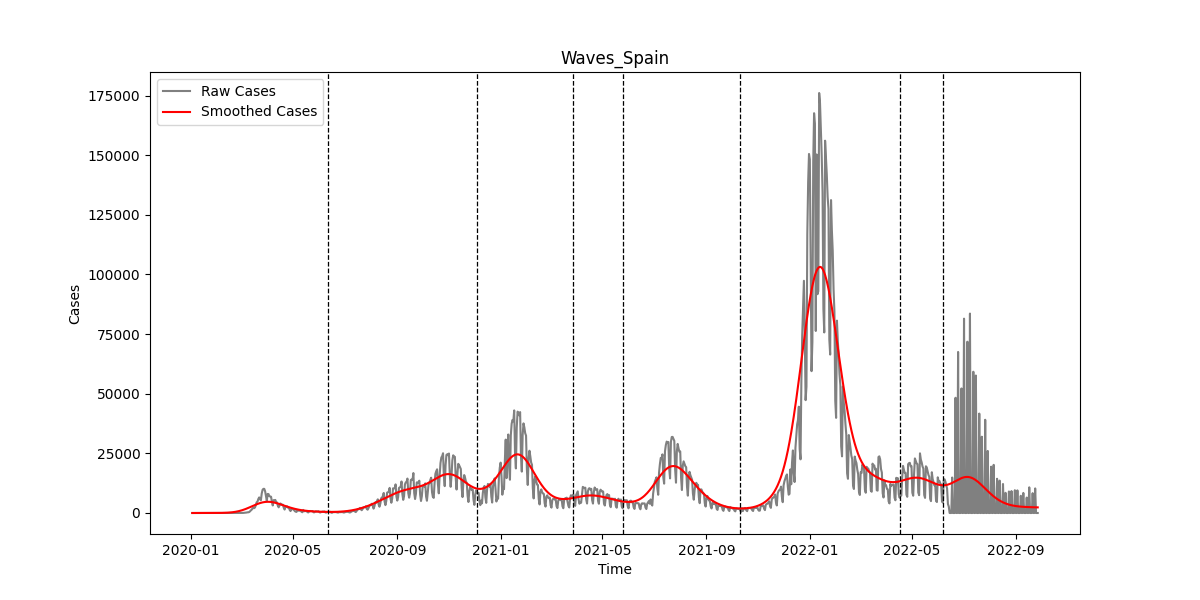

### SIFig12.png

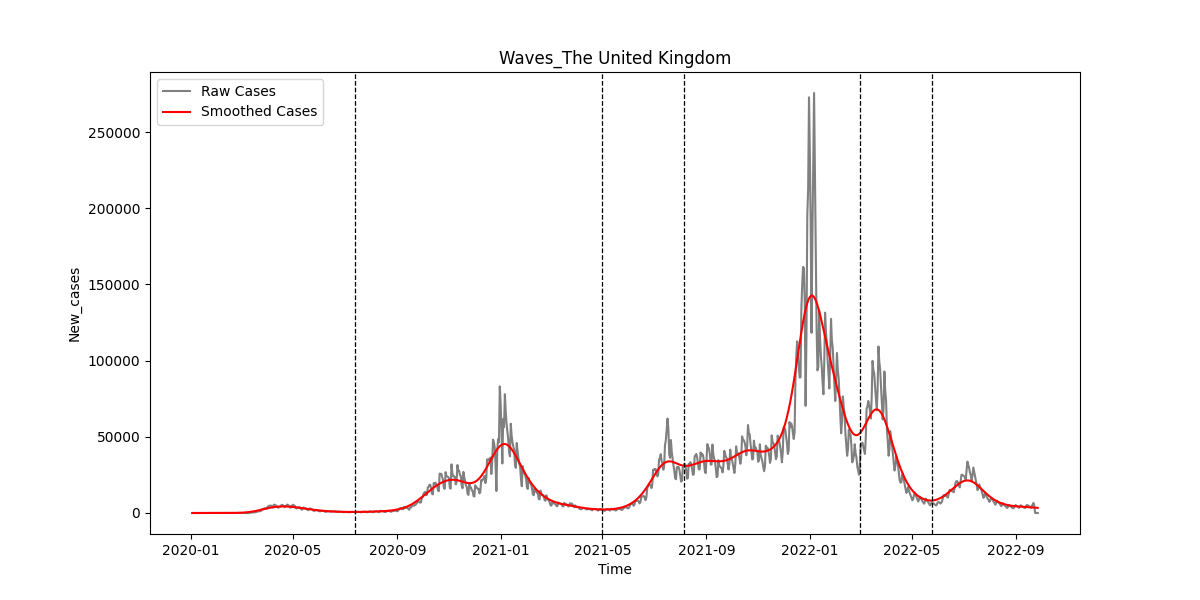

### SIFig13.png

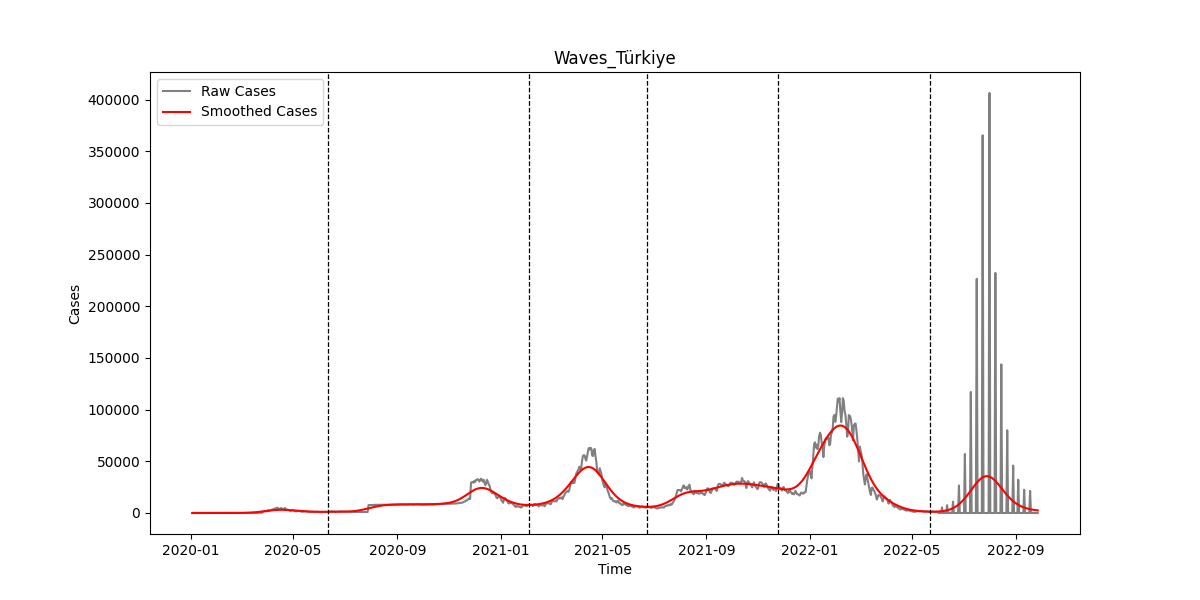

### SIFig14.png

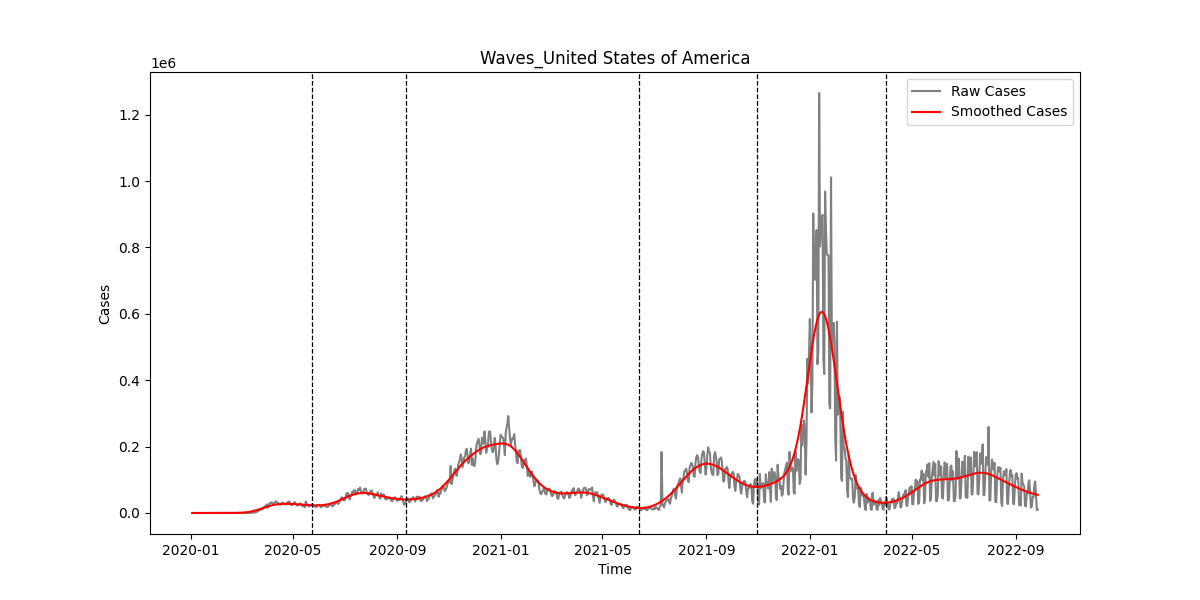

### SIFig15.png

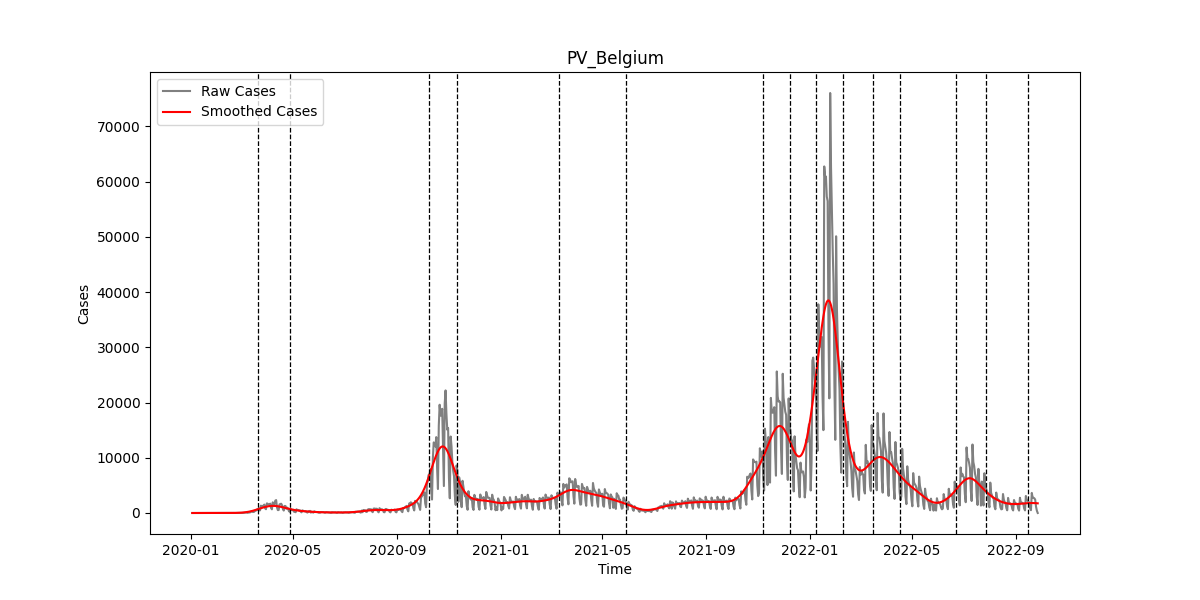

### SIFig16.png

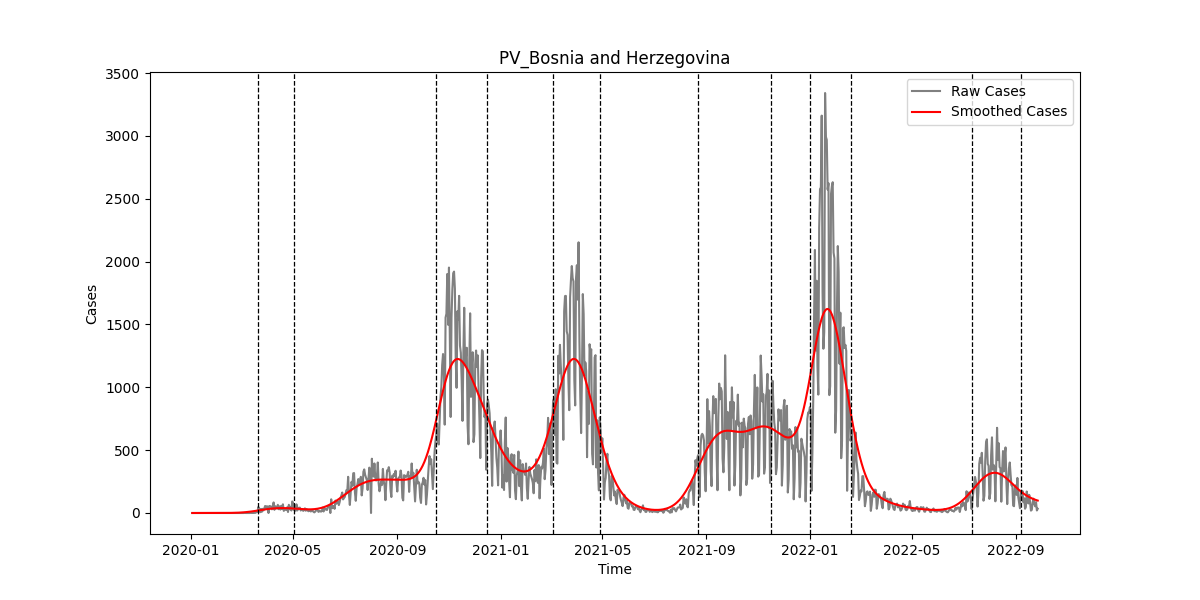

### SIFig17.png

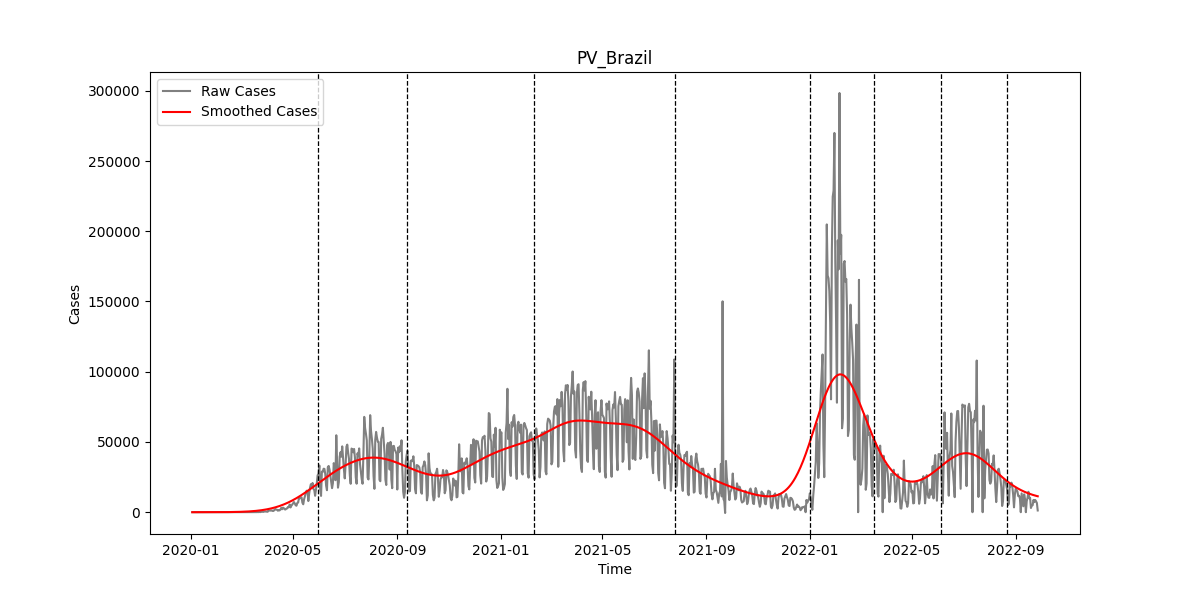

### SIFig18.png

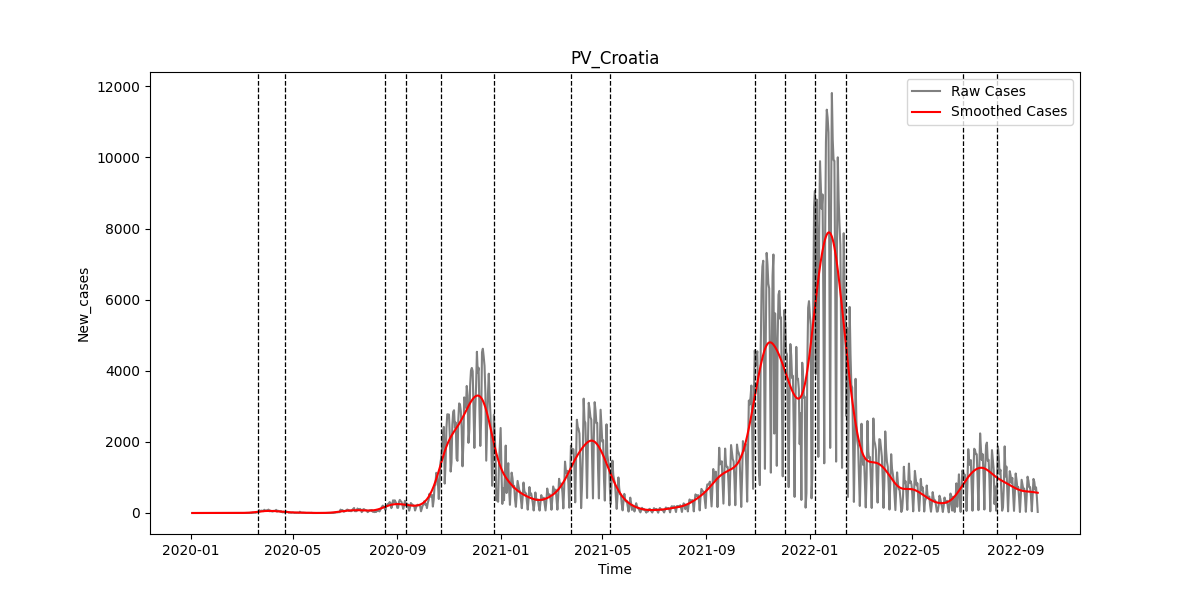

### SIFig19.png

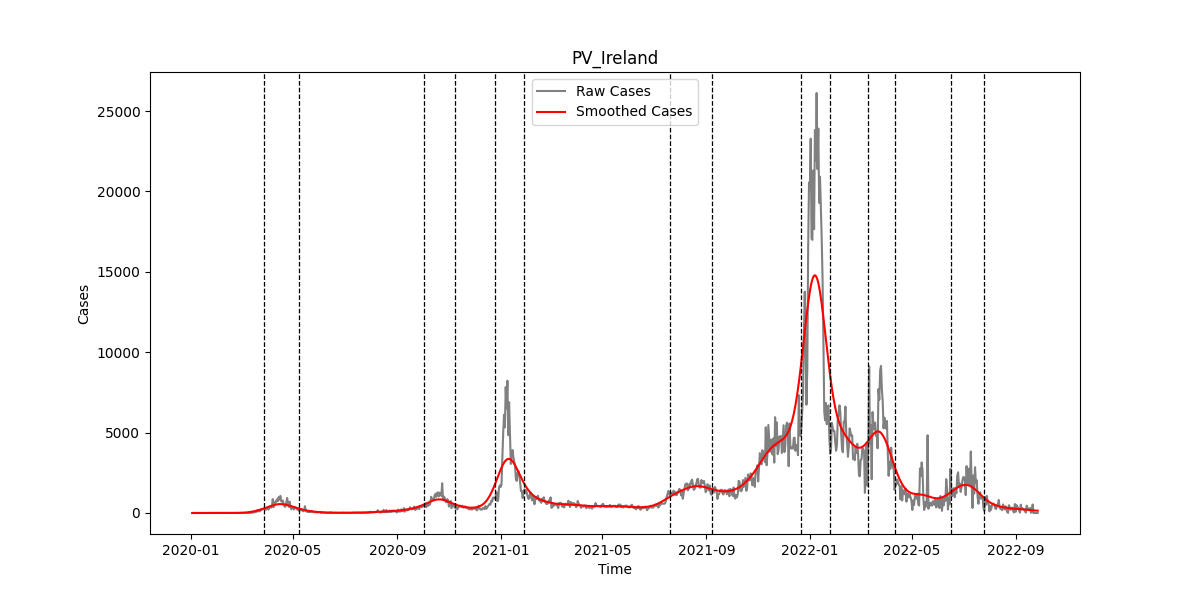

### SIFig20.png

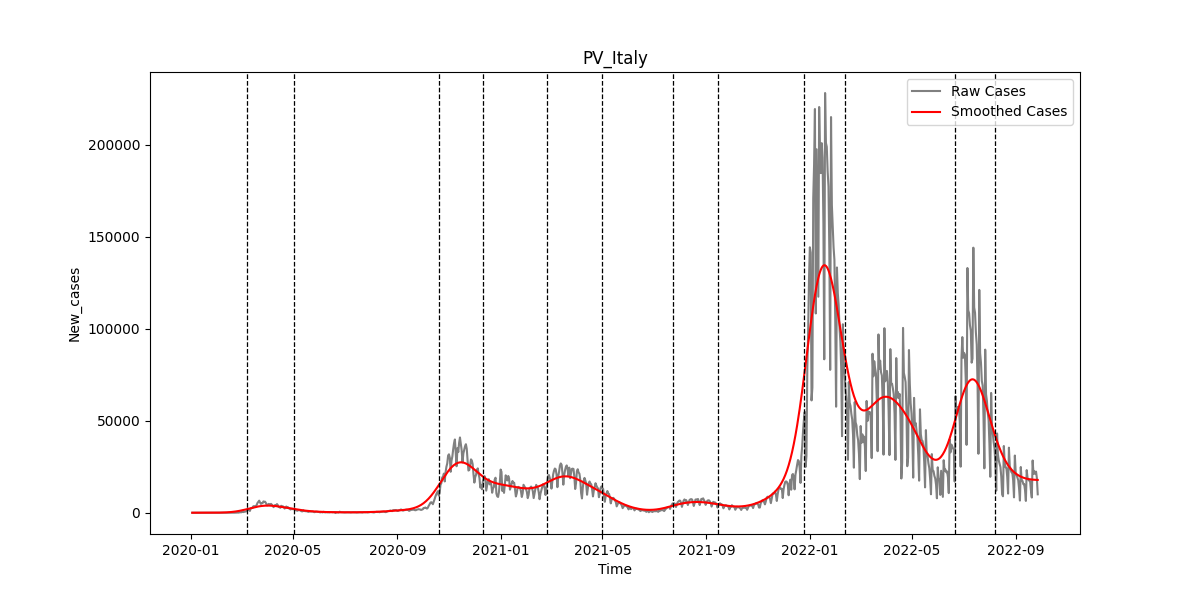

### SIFig21.png

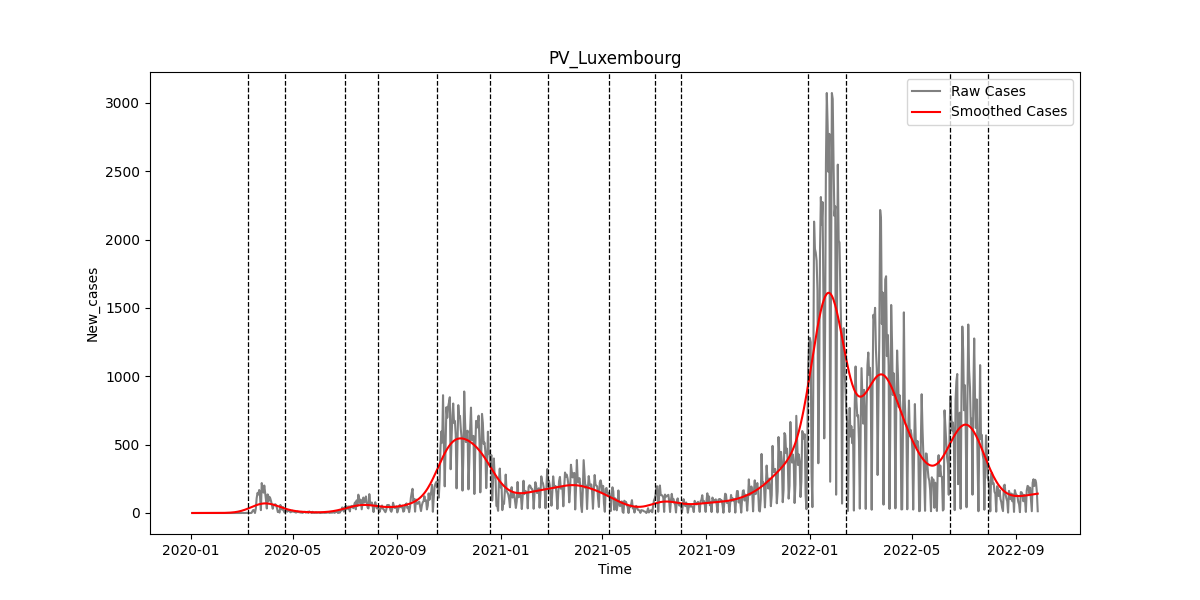

### SIFig22.png

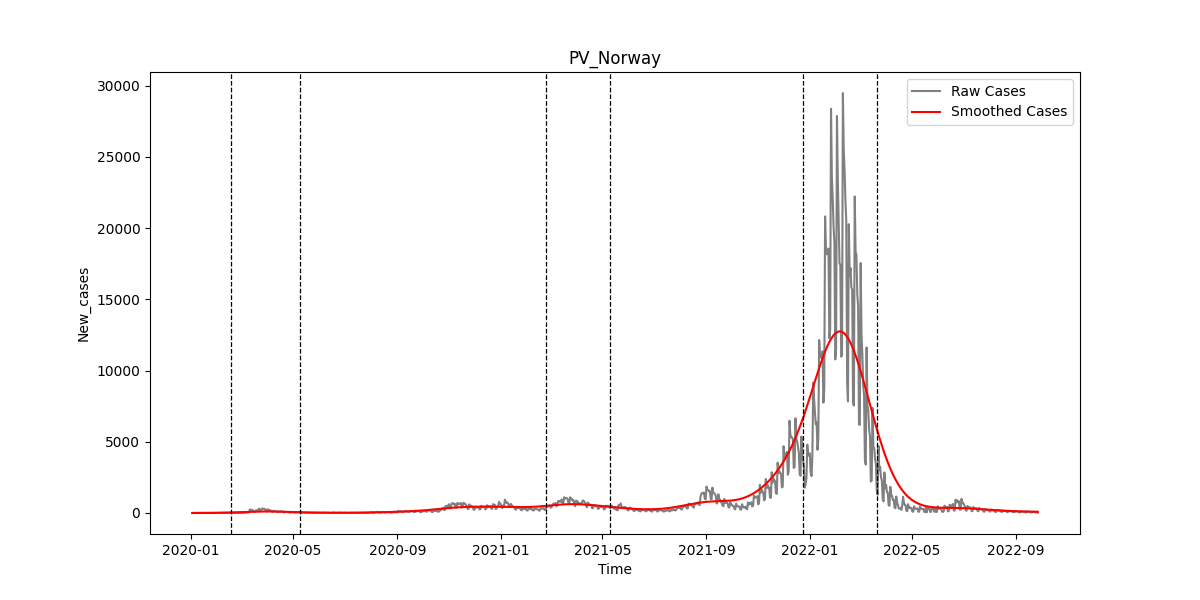

### SIFig23.png

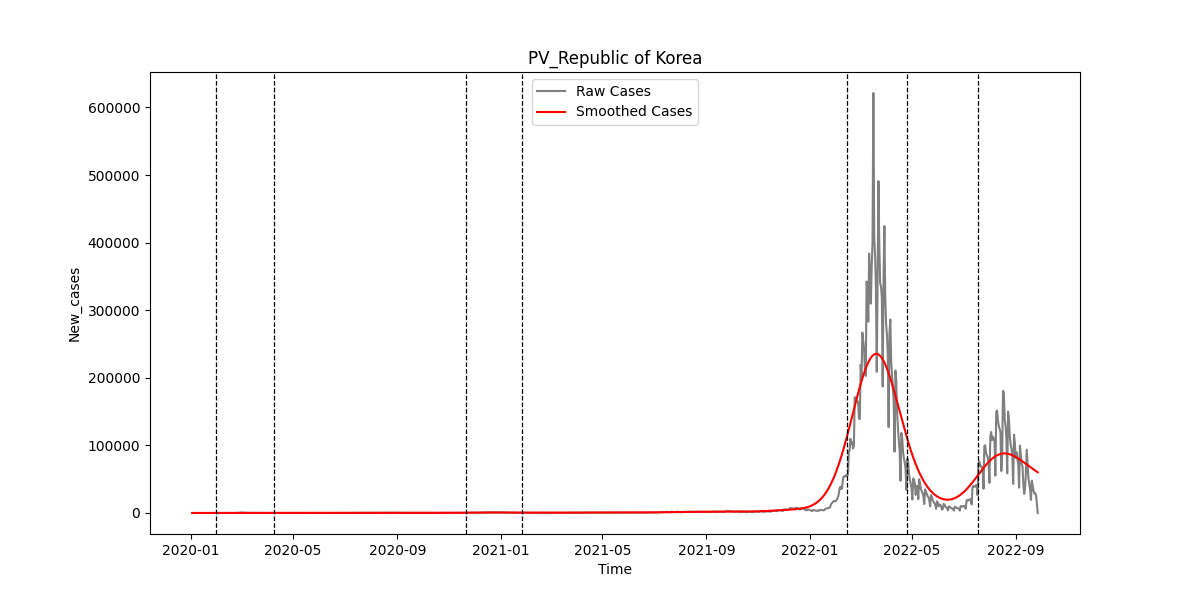

### SIFig24.png

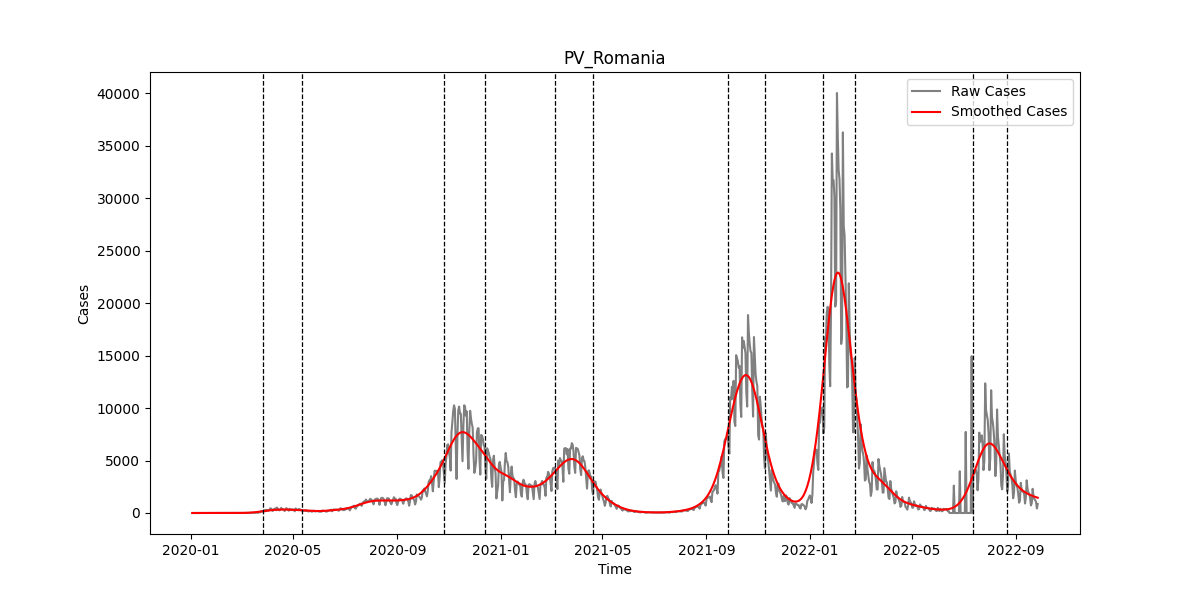

### SIFig25.png

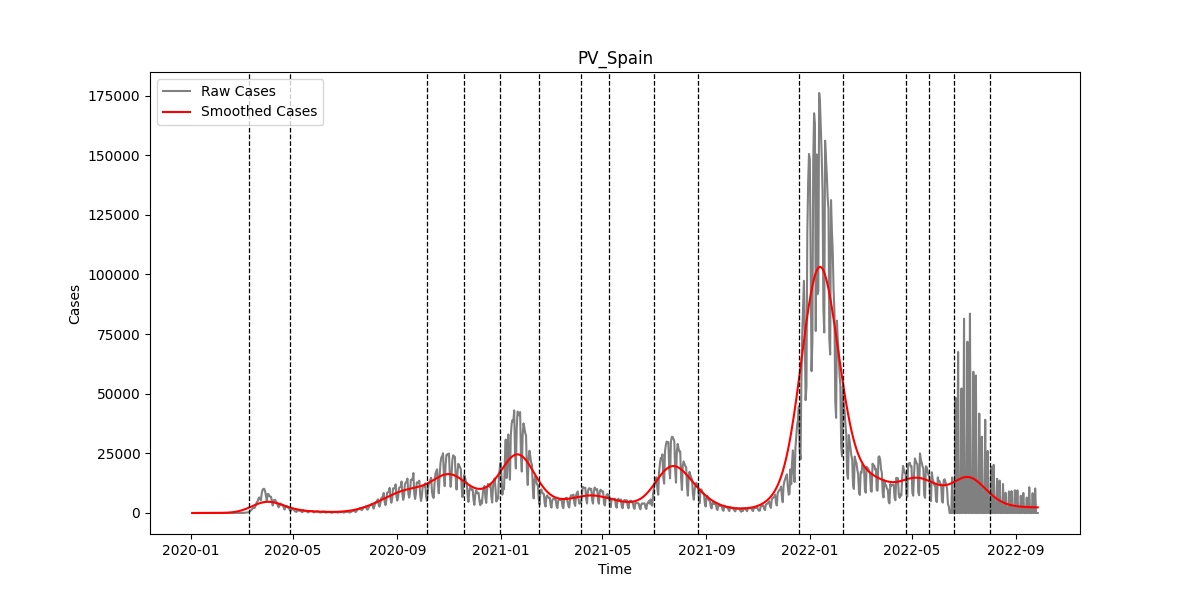

### SIFig26.png

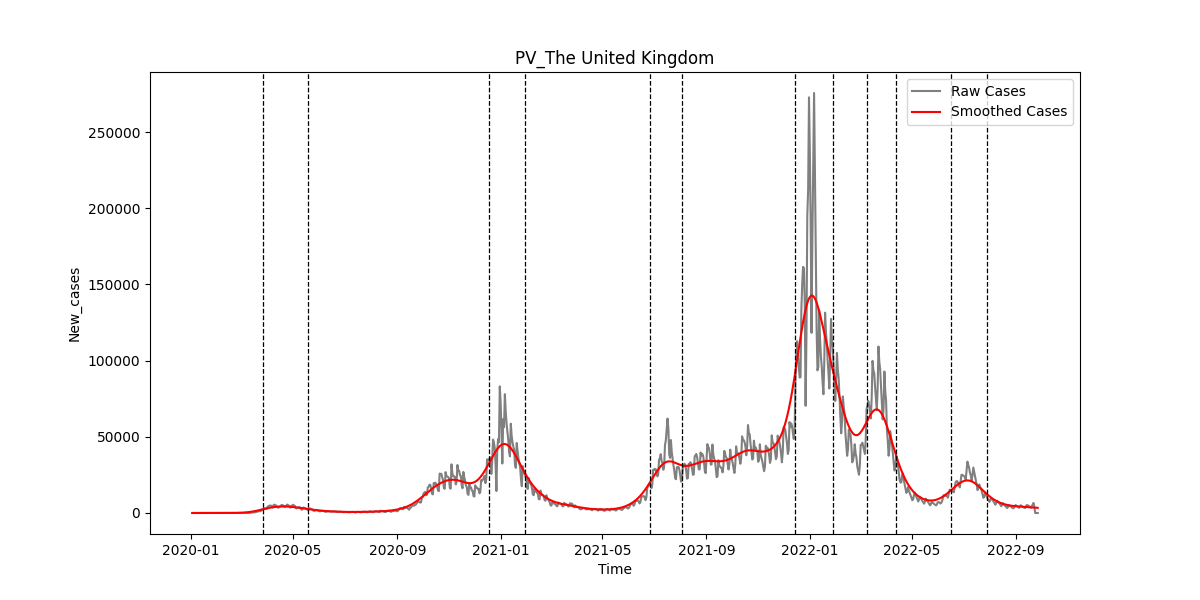

### SIFig27.png

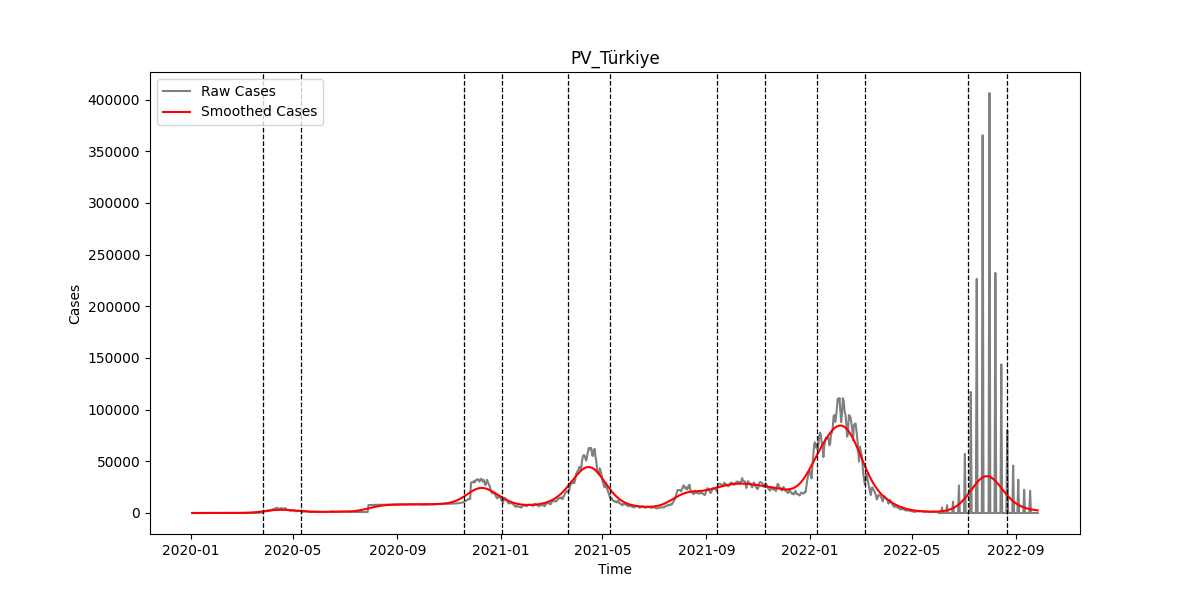

### SIFig28.png

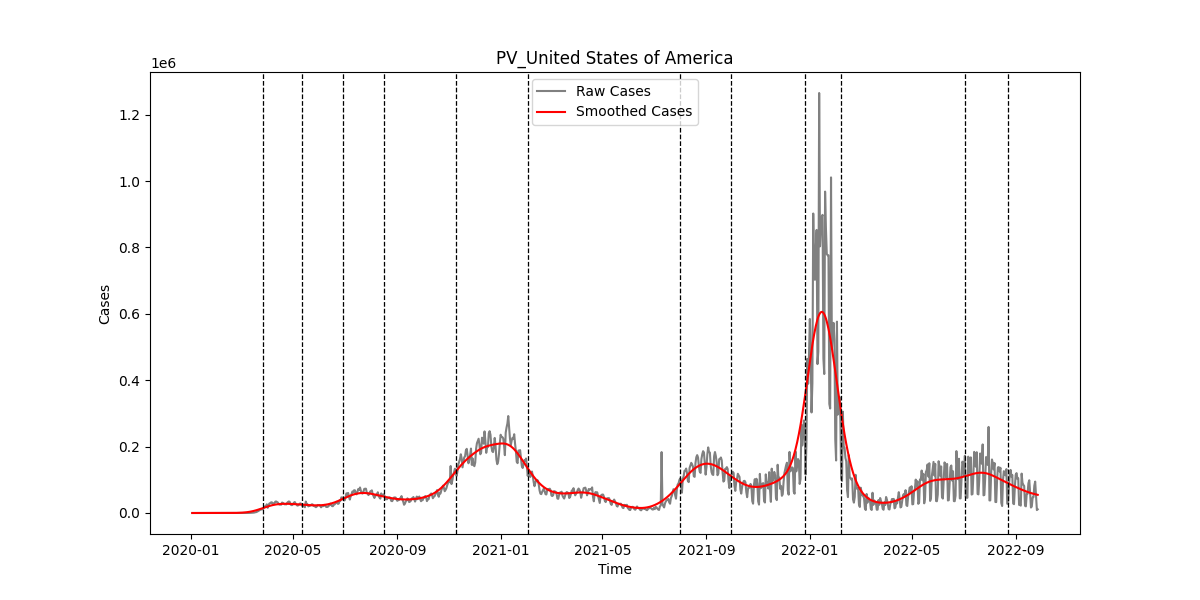

### SIFig29.png

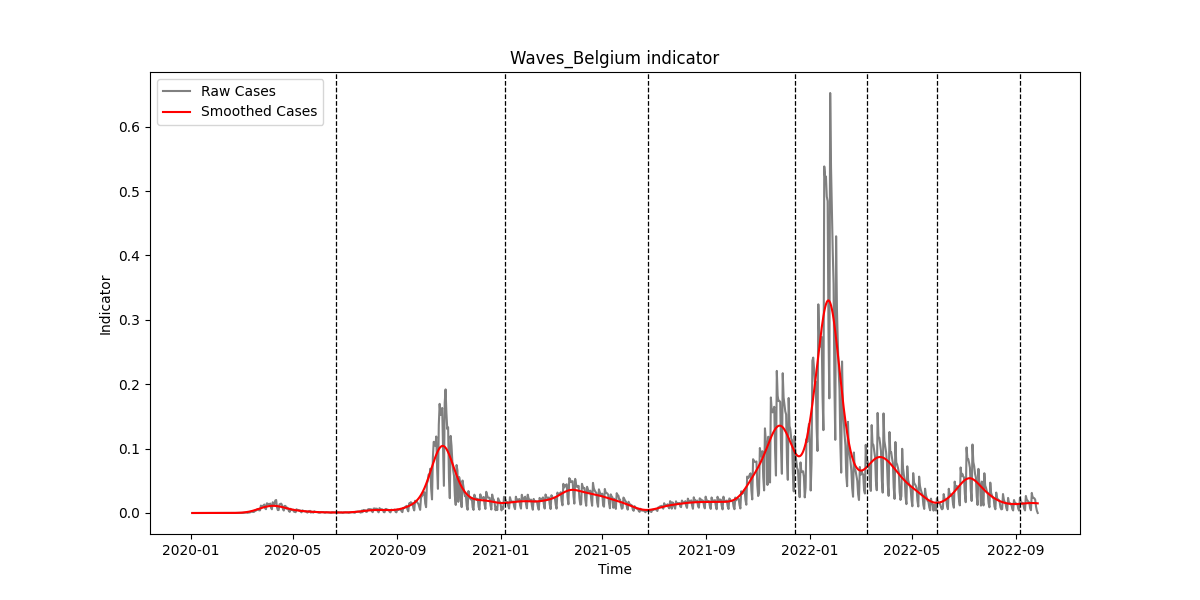

### SIFig30.png

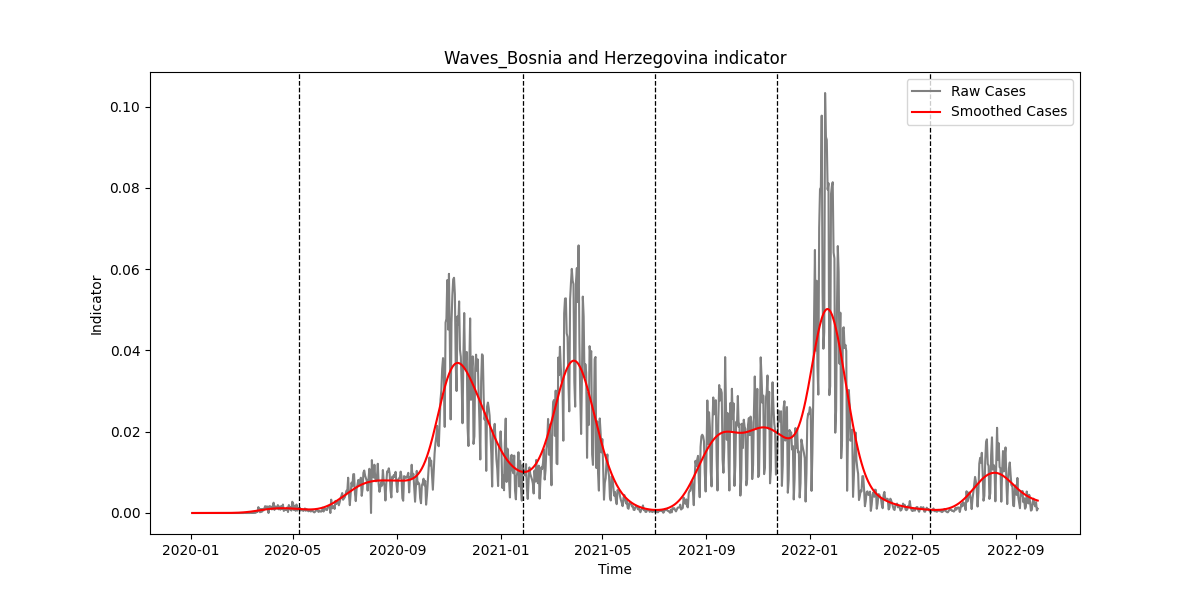

### SIFig31.png

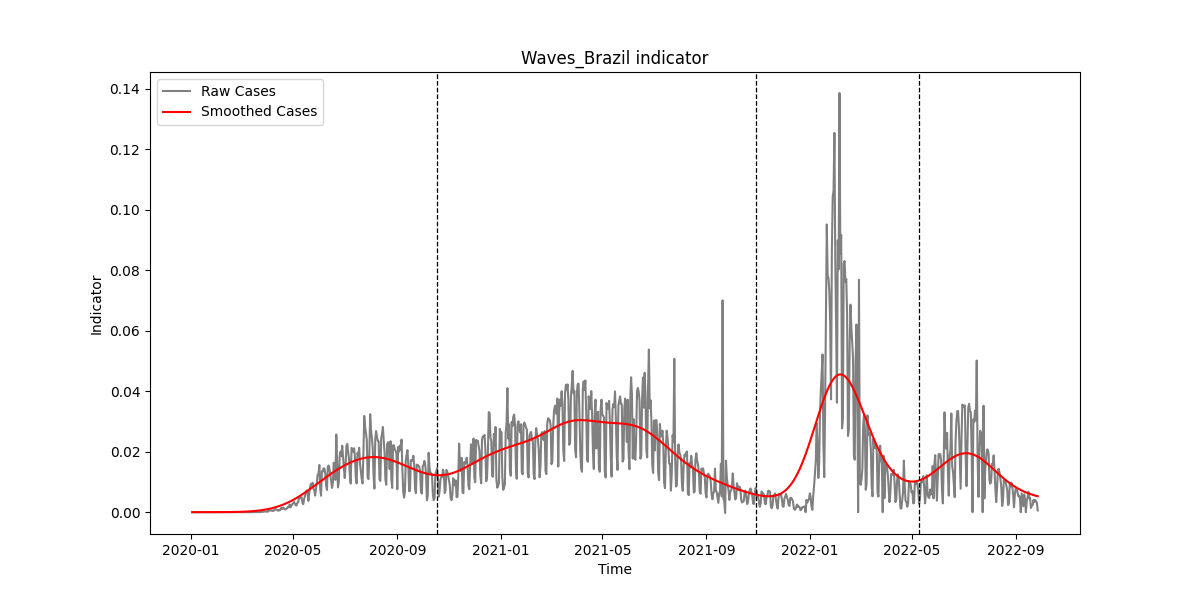

### SIFig32.png

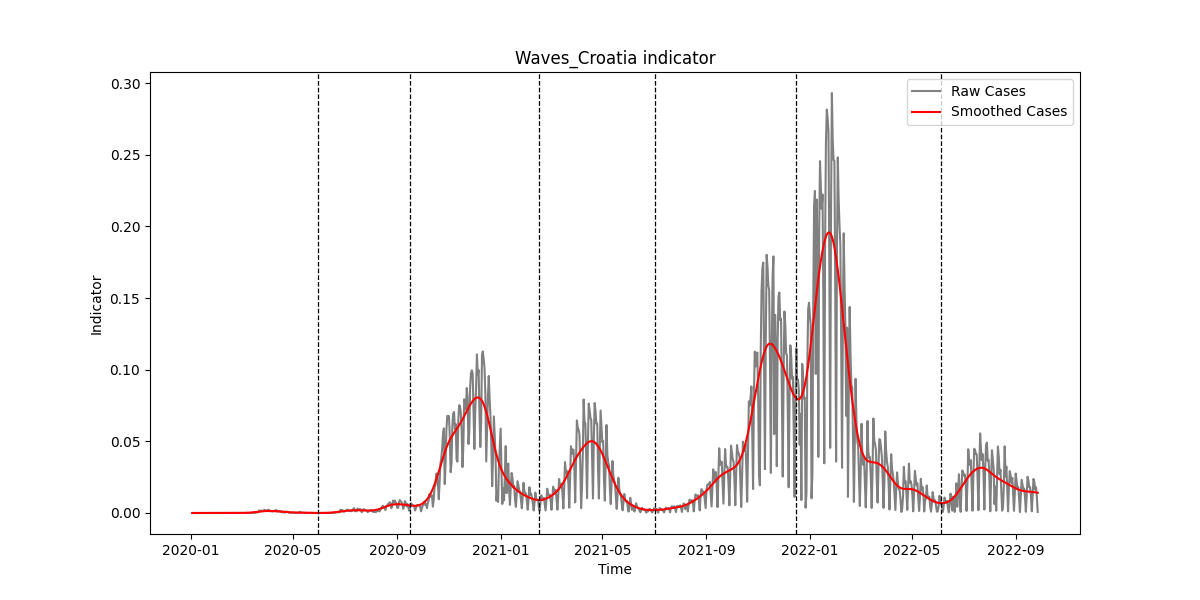

### SIFig33.png

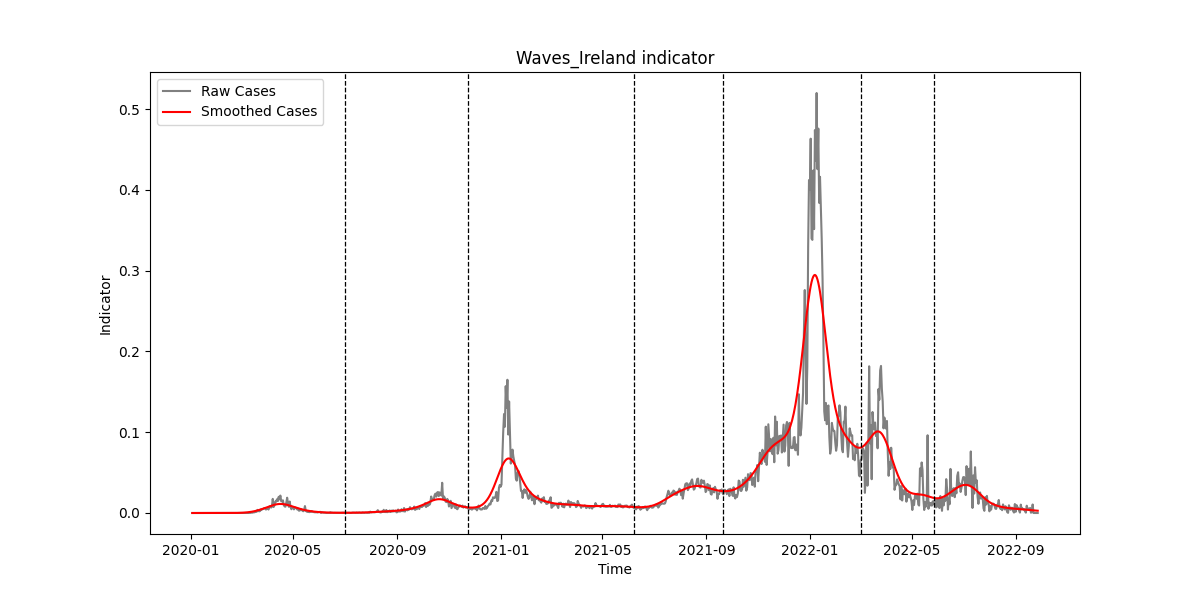

### SIFig34.png

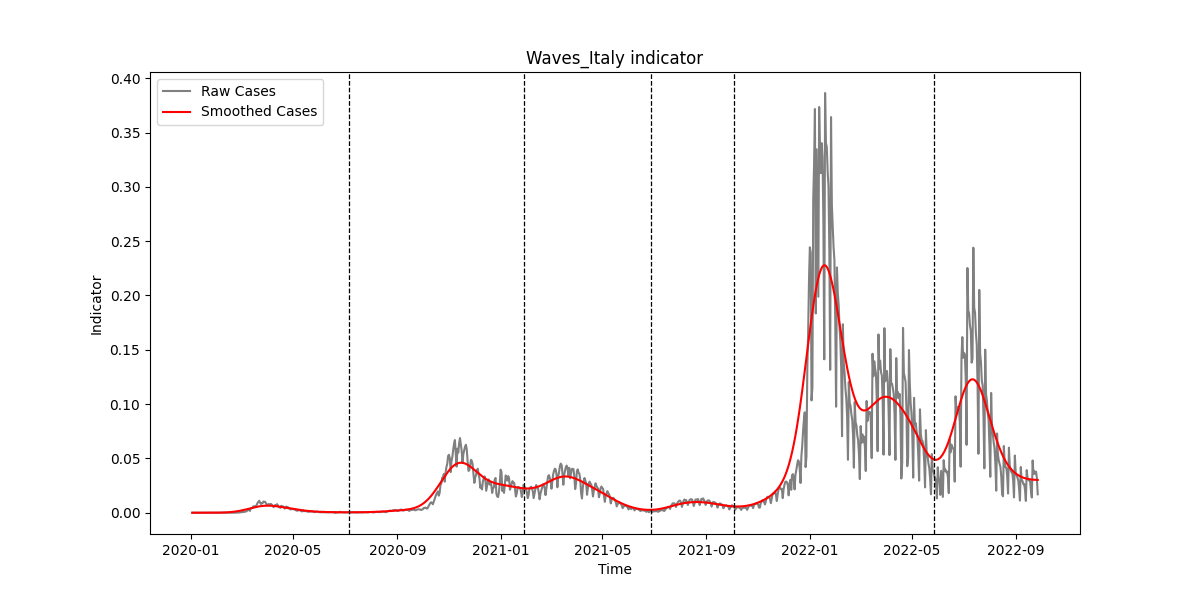

### SIFig35.png

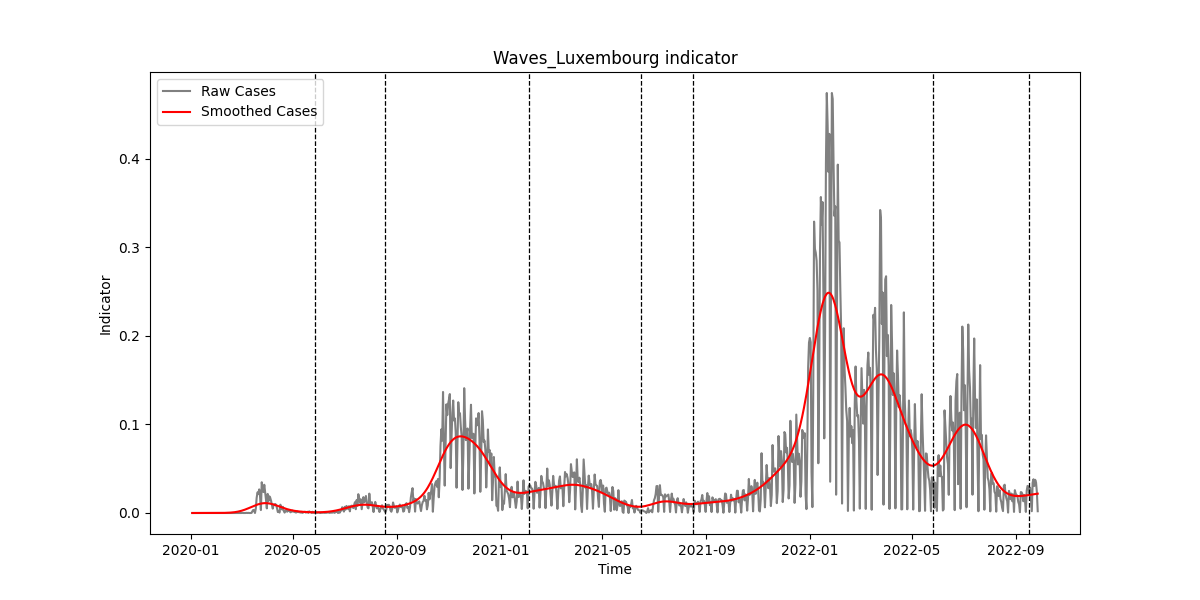

### SIFig36.png

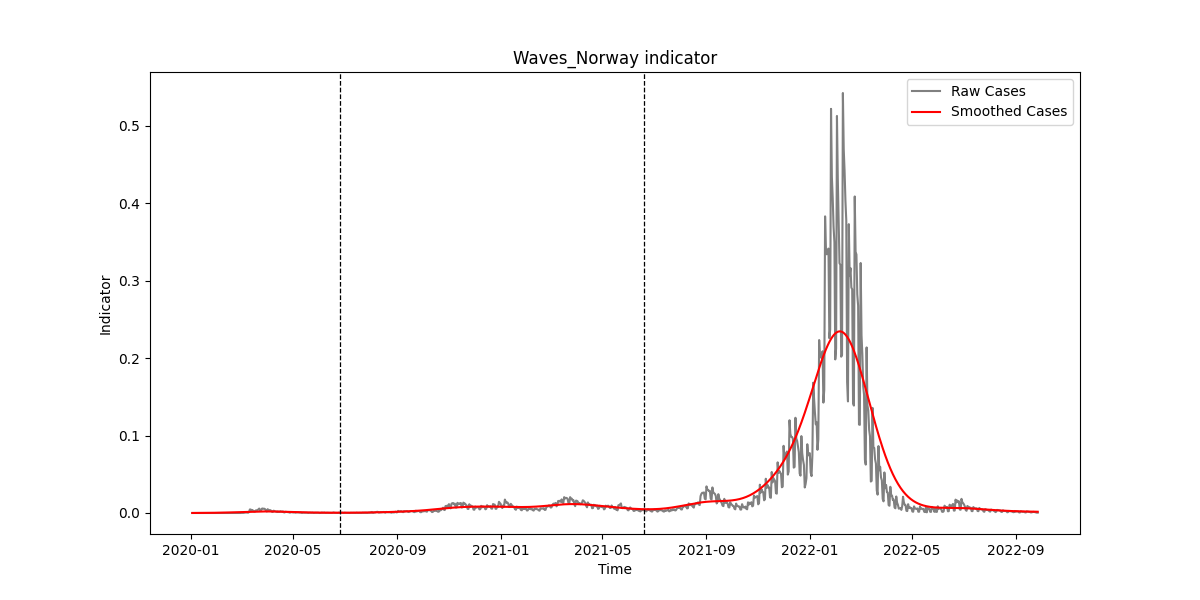

### SIFig37.png

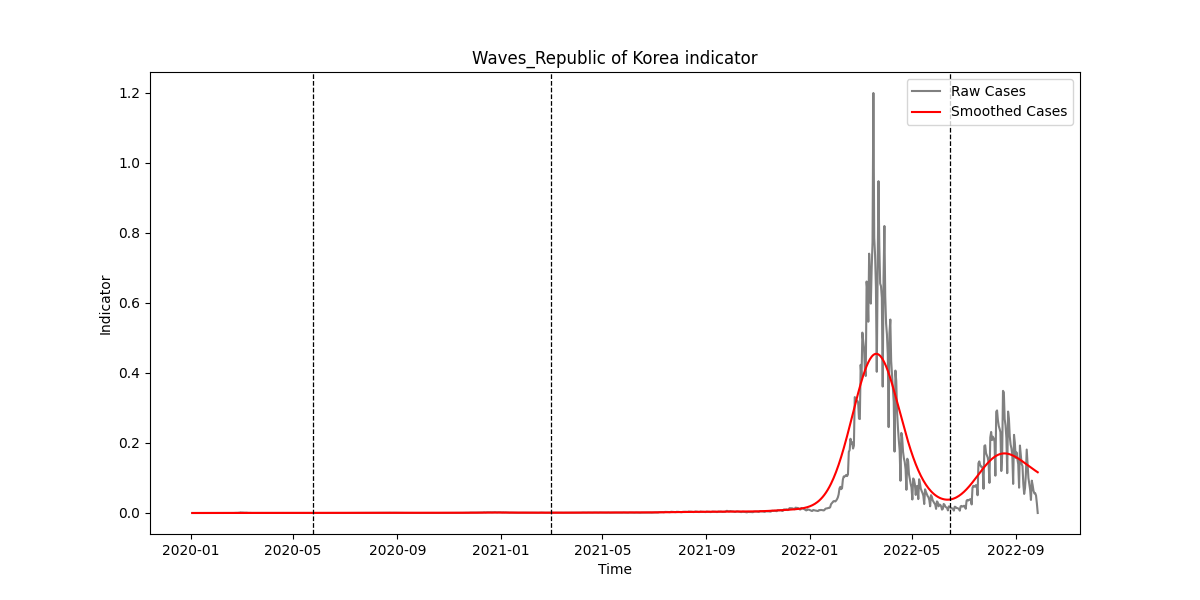

### SIFig38.png

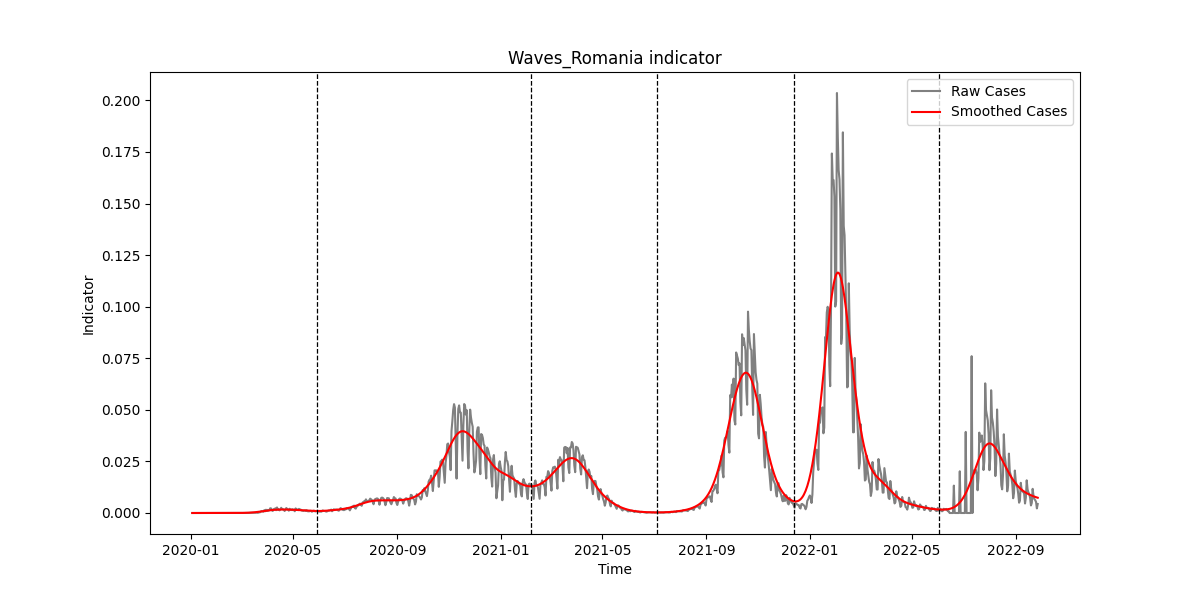

### SIFig39.png

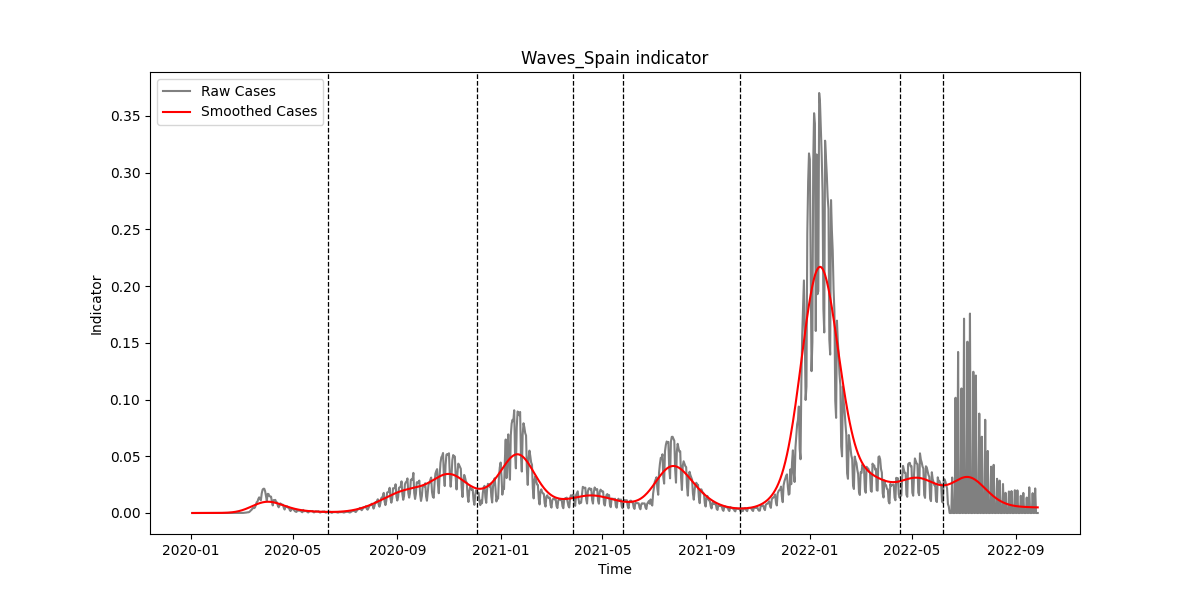

### SIFig40.png

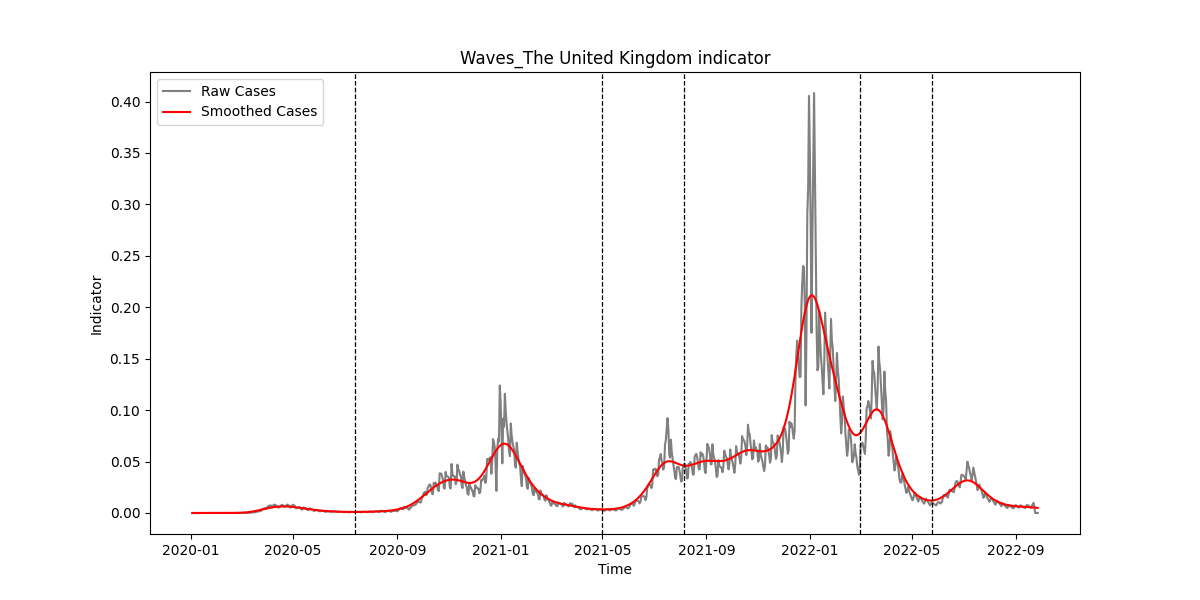

### SIFig41.png

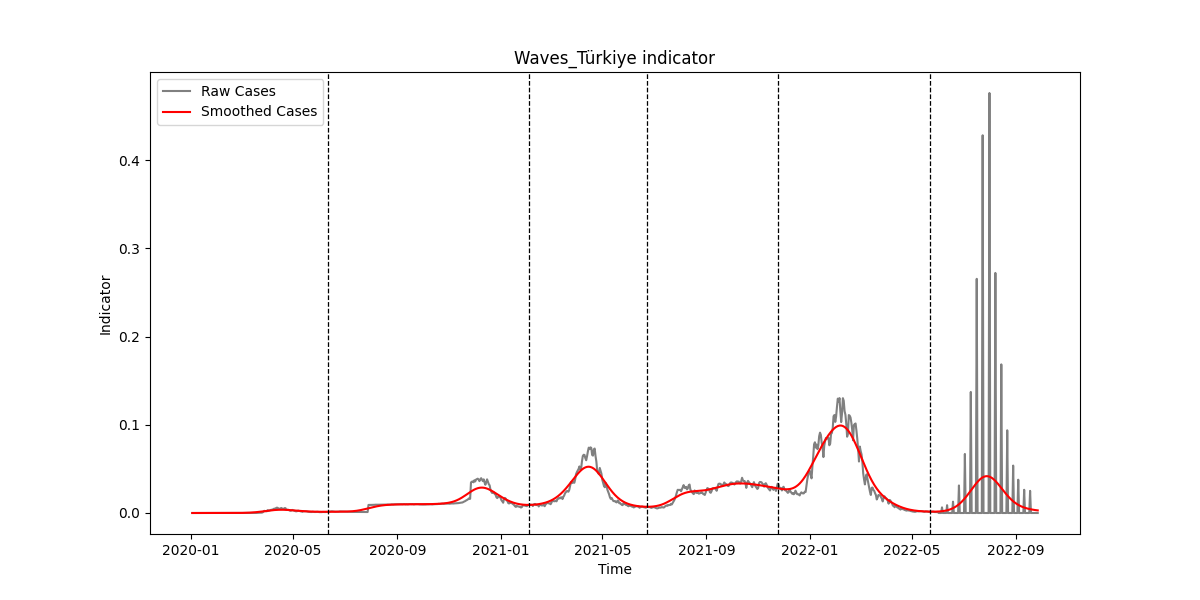

### SIFig42.png

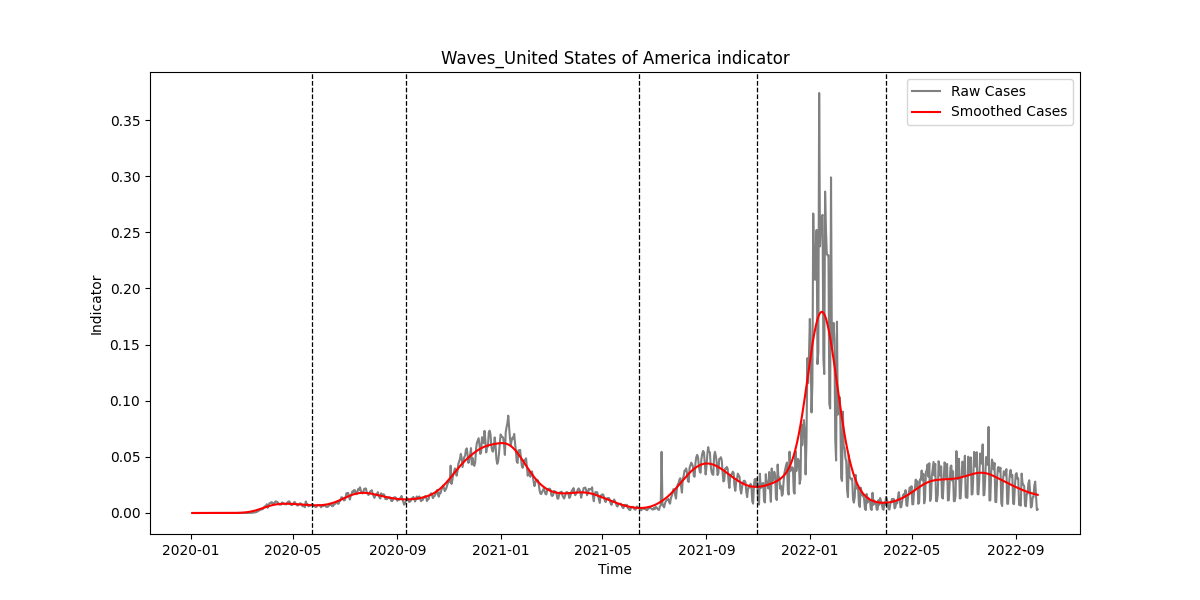

### SIFig43.png

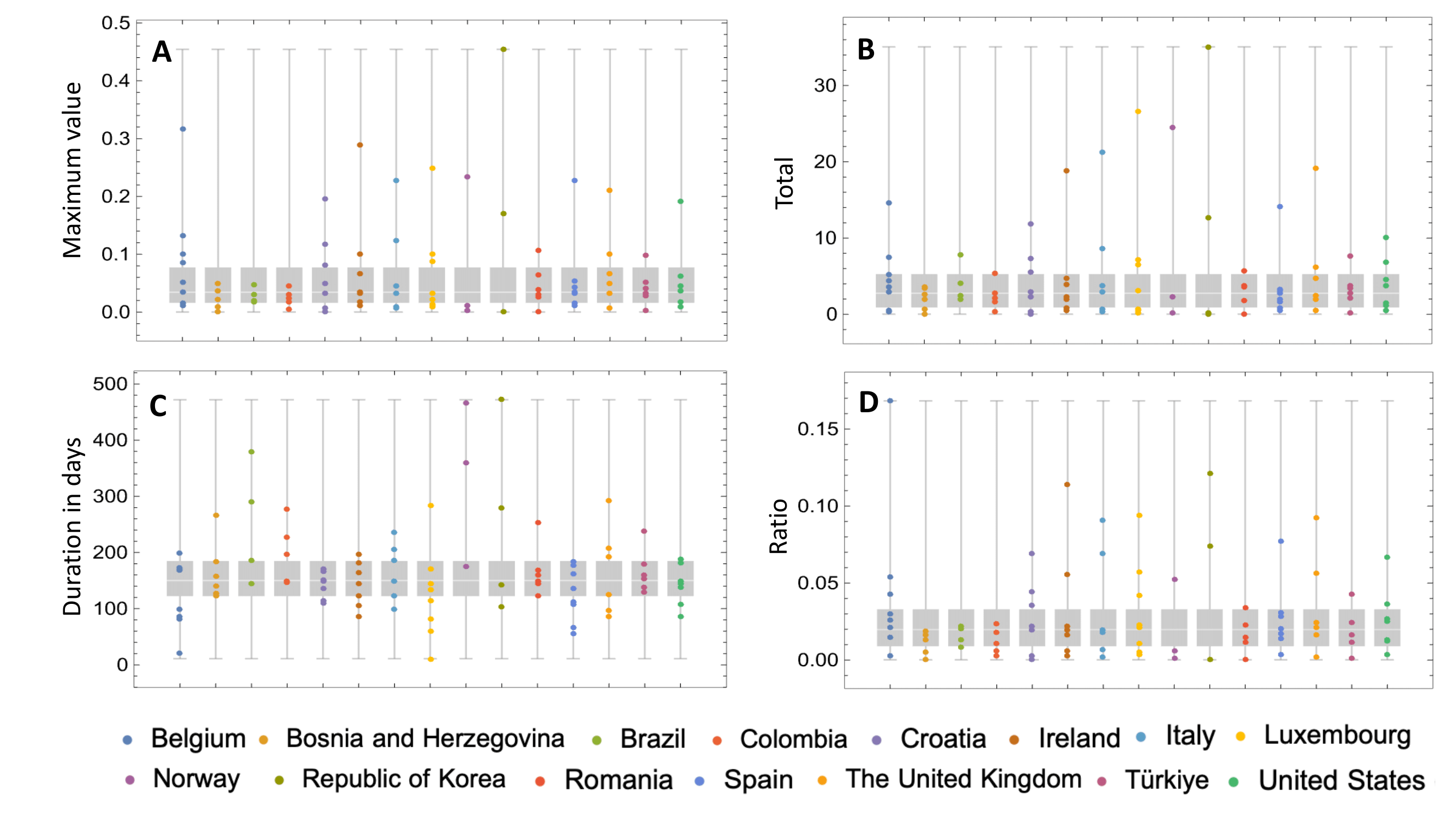
